# Integrating Multidimensional Data Analytics for Precision Diagnosis of Chronic Low Back Pain

**DOI:** 10.1101/2024.10.29.24316352

**Authors:** Sam Vickery, Frederick Junker, Rebekka Döding, Daniel L Belavy, Maia Angelova, Chandan Karmakar, Luis Becker, Nima Taheri, Matthias Pumberger, Sandra Reitmaier, Hendrik Schmidt

## Abstract

Low back pain (LBP) is a leading cause of disability worldwide, with up to 25% of cases become chronic (cLBP). Optimal diagnostic tools for cLBP remains unclear. Here we leveraged a comprehensive multi-dimensional data-set and machine learning-based variable importance selection to identify the most effective diagnostic tools for cLBP patient stratification. The dataset included questionnaire data, clinical and functional assessments, and spino-pelvic magnetic resonance imaging (MRI), encompassing a total of 144 parameters from 1,161 adults with (n=512) and without cLBP (n=649). Boruta and random forest were utilised for variable importance selection and cLBP classification respectively. Boruta variable selection led to pronounced variable reduction (median of all 15 datasets: 63.3%), while performing comparable to using all variables across all modality datasets. Boruta selected key variables from questionnaire, clinical, and MRI data were the most effective in distinguishing cLBP patients from controls. The most robust variables (n=9) across the whole dataset identified were psychosocial factors, neck and hip mobility, as well as lower lumbar disc herniation and degeneration. These critical variables outperformed all parameters in an unseen holdout dataset, demonstrating superior patient delineation. Paving the way for targeted diagnosis and personalized treatment strategies, ultimately enhancing clinical outcomes for cLBP patients.

## Introduction

In recent years, chronic low back pain (cLBP) has become one of the most prevalent and challenging conditions in clinical practice, affecting a significant portion of the global population ^1,2^. Despite its widespread occurrence, diagnosing and assessing cLBP remains difficult due to the complex and multifactorial nature of the disease ^3,4^, which encompasses physical, psychological, and social dimensions ^5^. Traditional diagnostic approaches often rely on self-reported symptoms, which can be subjective and prone to variability. Furthermore, surgical and non-surgical treatment outcomes are still inconsistent, refelected in a high rate of treatment-refactory in cLBP patients ^6^. As such, there is an increasing interest in exploring more objective and comprehensive methods that incorporate multimodal data, including medical imaging, physical assessments, clinical evaluations, and patient-reported outcomes.

Machine learning algorithms provide models for identifying distinct subgroups that can help explain the occurrence and characteristics of a disease ^7,8^. A previous systematic review by our team using machine learning applications in LBP ^9^ highlighted that a narrow range of mechanistic domains have been assessed, and sample sizes in these studies were consistently small, ranging up to only 171 participants. Consequently, using limited data and modalities limits the robustness and applicability of such models. Through gathering many data points across multiple modalities one can ascertain which variables and modalities are the most informative at distinguishing cLBP patients from asymptomatic controls. Reducing the number of variables to those that are most informative has been previously employed in predictive and classification modelling to improve accuracy ^10,11^. This approach can be applied as the main outcome and not only in model preprocessing, in order to obtain a data-driven decision on the most important variables in multi-dimensional clinical data. Such a systematic data-informed investigation of back pain diagnosis in a large multi-modality sample is lacking to help inform future studies in selecting which data to acquire and for clinicians in which tests to conduct. Therefore, the aim of this study was to identify and compare the most informative domains and variables in delineating patients with and without cLBP utilising a large multi-modality dataset.

## Results

### Study sample

The prospective cross-sectional study draws its data from the ongoing “Berliner Rückenstudie” (“Berlin Back Study”; https://spine.charite.de/en/spine_study/; running time: 01/01/2022 to 31/12/2025), which was registered at the German Clinical Trial Register (DRKS-ID: DRKS00027907). Recruitment procedures vary from local promotion (i.e., postal flyers, notice boards, internet approaches, and social media) at the Charité-Universitaetsmedizin Berlin, in the general public (i.e., newspapers, magazines, podcasts) to cooperation with local companies, administrative authorities, and word-of-mouth. The protocol is in accordance with the Helsinki Declaration of ethical principles ^12^ and has been approved by the Ethics Committee of the Charité – Universitätsmedizin Berlin (registry numbers: EA4/011/10, EA1/162/13). Written informed consent was obtained from all participants. The STROBE guideline ^13^ (Supplementary Table S1) and TRIPOD statement ^14^ (Supplementary Table S2) for prediction model development were used to report this study. Data collection started on 1st January 2022 and cut-off for inclusion in the current analysis was 5th April 2024. Data collection occurred in a research centre within a university-hospital.

Study participants were recruited through a telephone interview and excluded if they met any exclusion criteria, as well as some excluded at the testing site (Supplementary Table S3). A total of 1273 participants were included in the study at cut-off point. These participants were initially guided through self-administered questionnaires by a study coordinator. Then they continued to a clinical examination by a trained medical doctor, which included physical examinations, questions, as well as a back shape and function test. The examinations and questionnaires took a total of 90 minutes to complete. Additionally, participants were offered a magnetic resonance imaging (MRI) within 14 days of the spino-pelvic region. During the clinical assessment the participants were classified by the clinician as asymptomatic (no back pain), symptomatic (cLBP), or previously suffering from cLBP. To ensure a more robust cLBP patient classification previous symptomatic subjects were removed from the sample. The criteria for designation as cLBP was daily back pain for more than three months. Furthermore, participants who revoked their inclusion in the study and those who were missing demographic data; age, sex, body mass index (BMI), and patient status were removed. This resulted in a study sample of 1161 subjects that included 649 asymptomatic (19 – 72 years old, mean age = 40.7 ± 12.6, females = 353) and 512 cLBP (19 – 65 years old, mean age = 43.5 ± 11.7, females = 306) participants (Table 1). This sample was sub-divided into four modalities; questionnaires (Q), clinical physical assessment (C), back shape and function (S), and MRI (M). Each modality was combined with demographic data (age, sex, and BMI) and then joined with all combinations of the four modalities, resulting in 15 datasets (Figure 1A, and Supplementary Table S4 - S18). Both complete datasets (Fig. 2) and imputed datasets were used for cLBP classification (Fig. 1B). Imputation can lead to bias results, in particular when it comes to variable importance selection ^15^. Therefore, presentation of variable importance was only considered for non-imputed datasets. Highly correlated (r > 0.9) variables were removed from the daatsets to reduce colinearlity between dataset variables (Supplementary Figure S1 – S6). An overview of all Berlin Back Study variables (Supplementary Table S19), those removed during preprocessing (Supplementary Table S20), and a list of all variables (144) used for modelling is presented in Supplementary (Supplementary Table S21).

**Figure 1.**
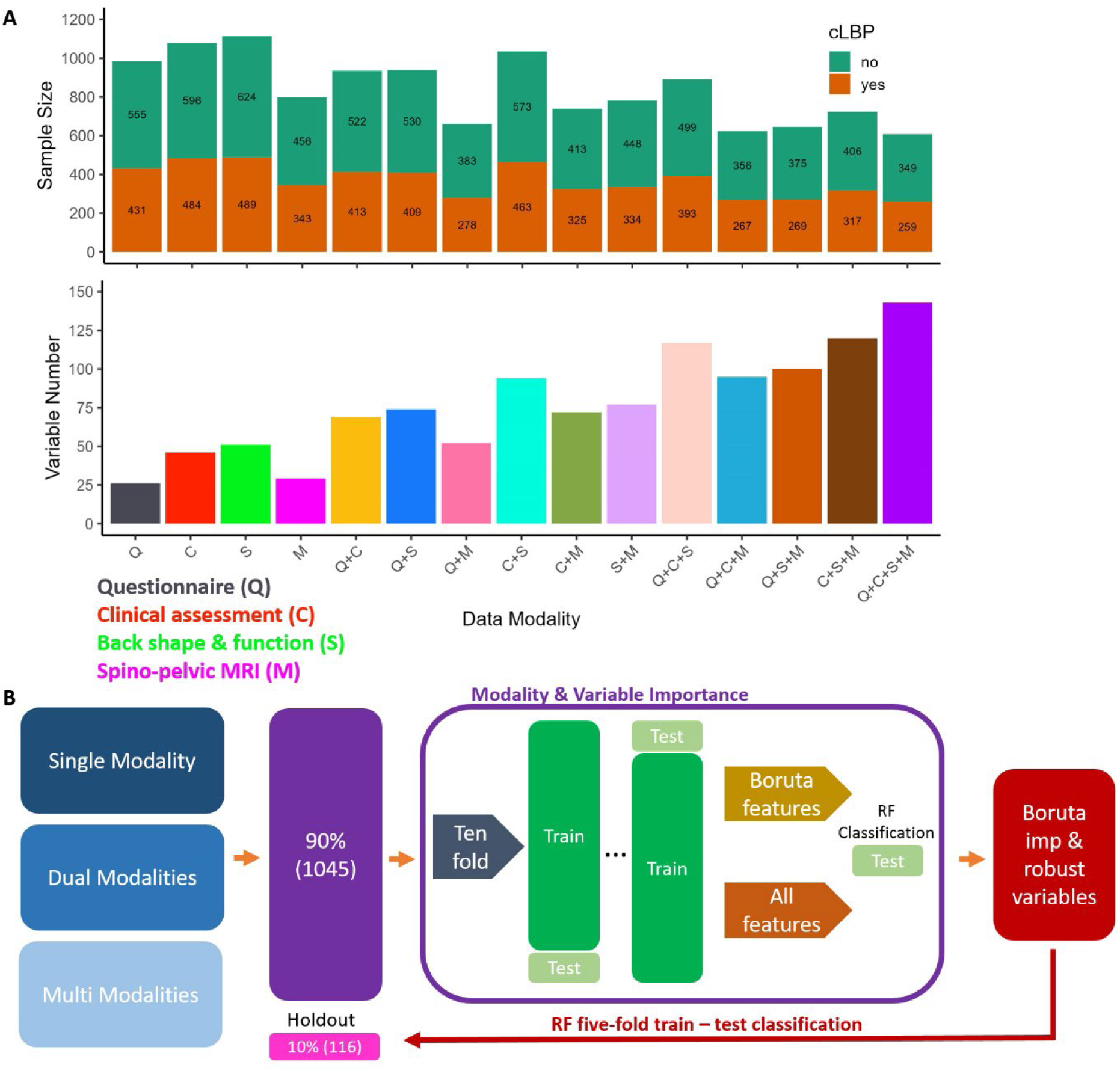
Modality dataset distributions and machine leaning workflow. A – Top shows the chronic low back pain (cLBP) sample size distribution across all 15 dataset modalities. Bottom presents the number of variables used for cLBP classification and variable importance selection across the 15 dataset modalities. B – Represents the machine learning workflow implemented to compare the different modalities and determine the most important variables for cLBP patient delineation using a random forest binary classification algorithm for training and testing.

**Figure 2.**
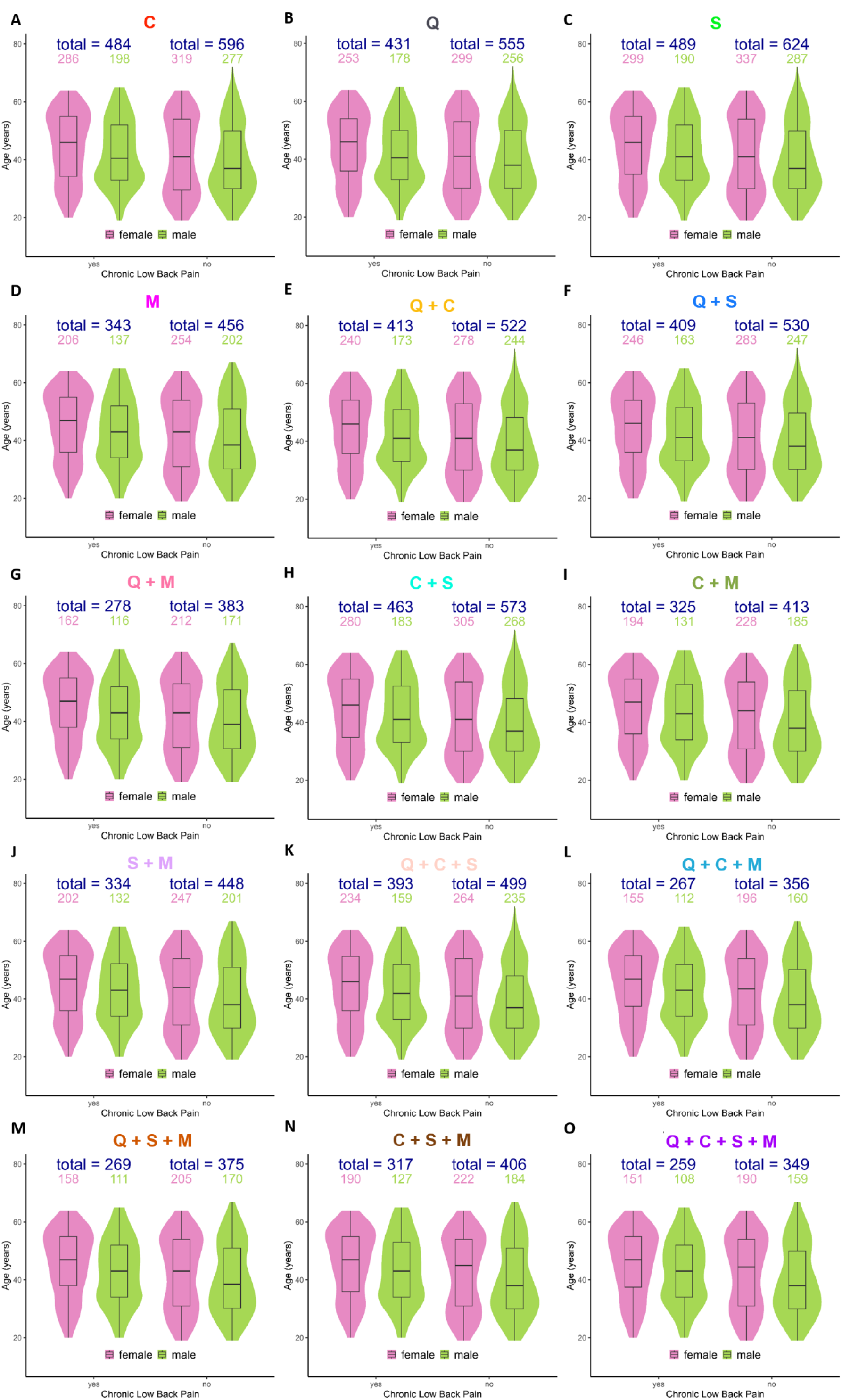
Modality datasets age, sex, and cLBP distributions. Violin plots for non-imputed dataset A - questionnaire; B – clinical assessment: C – back shape and function; D – MRI; E – questionnaire + clinic; F – questionnaire + back shape and function; G – questionnaire + MRI; H – clinic + back shape and function; I – clinic + MRI; J – back shape and function + MRI; K – questionnaire + clinic + back shape and function; L – questionnaire + clinic + MRI; M – questionnaire + back shape and function + MRI; N – clinic + back shape and function + MRI; O – all datasets.

**Table 1.**
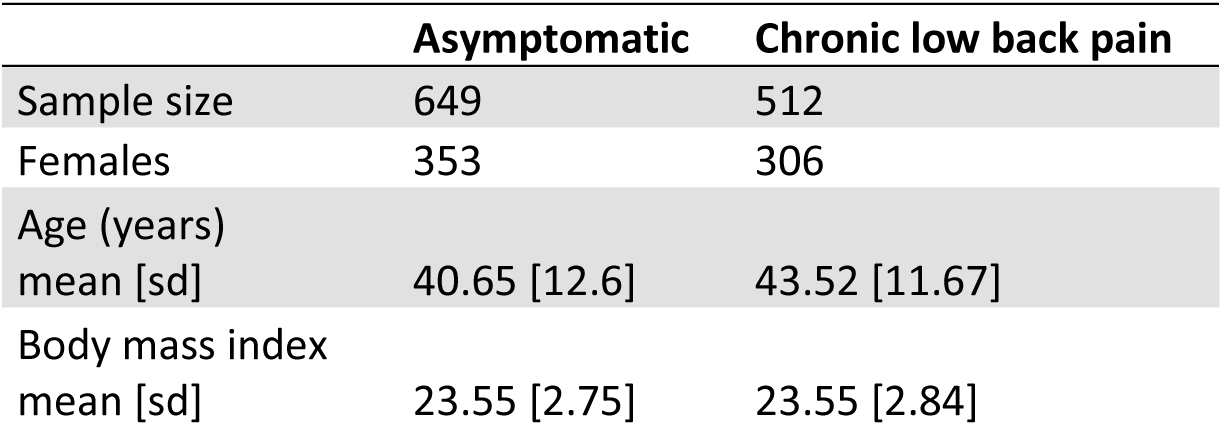
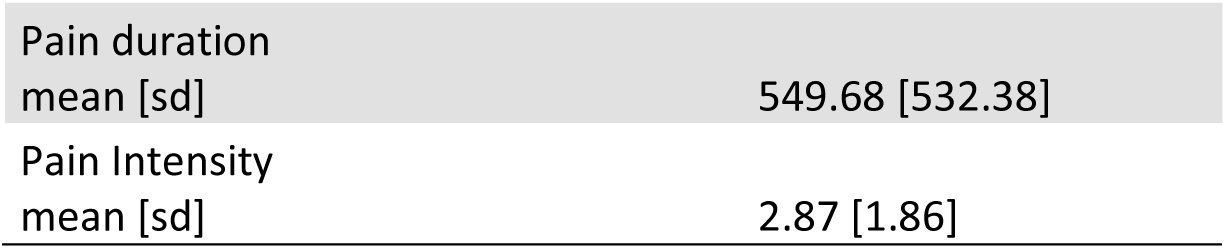
Demographic data of entire Berlin Back dataset.

### Chronic low back pain classification

Determining modality and variable importance was conducted on 90% (n = 1045, 19 – 72 y/o, mean age = 41.71 ± 12.28, cLBP = 469) of the whole dataset, with 10% (n = 116, 20 – 64 y/o, mean age = 43.81 ± 12.12, cLBP = 43) used as a hold-out sample to test cLBP patient classification performance of the most robust and important variables (Fig. 1B). Variable selection and importance was determined using the Boruta algorithm ^16^ on the training data within an iterative ten-fold train-test loop across all 15 modality datasets. Boruta is a wrapper function selects important variables and removes unimportant variables using random random forest (RF) ^17^. A binary RF classification model was trained using the Boruta ^16^ selected variables as well as all variables to classify cLBP status in the test sample across the ten-fold train-test loop. Furthermore, imputation using missForest ^18^ was independently conducted on the training and test datasets within the ten-fold loop (Supplementary Table S23). Imputed datasets performed generally slightly worse than the full datasets. We used AUC (area under the receiver operating characteristic (ROC) curve) as the main model performance metric. AUC provides a good combination of sensitivity and specificity for comparing Boruta selected variables to all variables and the different modality datasets in their performance of cLBP patient delineation.

The best single modality for cLBP classification was Boruta selected variables from MRI (Fig. 3A) with a mean AUC of 0.645 with a 95% confidence interval (CI) of 0.618 – 0.672 and accuracy of 0.657 (95% CI, 0.636 – 0.678). The Boruta selected questionnaire dataset modality produced only minimally worse classification performance (mean AUC = 0.631, 95% CI = 0.610 – 0.652) than Boruta reduced and all MRI variables (mean AUC = 0.637, 95% CI = 0.599 – 0.675) using the least amount of variables (mean = 8) across all modality datasets (Fig. 3A-C). Dual and multi modalities generally performed better than single modality models in cLBP classification. Questionnaire, clinical physical assessment, and MRI (Q + C + M, Fig. 3C) modality with Boruta selected variables represents the best performing modality model with a mean AUC of 0.699 (95% CI, 0.669 – 0.729) and a mean accuracy of 0.709 (95% CI, 0.679 – 0.739). Moreover, this model showed the highest sensitivity (mean = 0.622, 95% CI – 0.568 – 0.676). The modality model showing the highest specificity was using all variables and all modalities (Q + C + S + M, mean = 0.840, 95% CI = 0.799 – 0.881). The three best dual modalities models, C + M (mean AUC = 0.679, 95% CI = 0.633 – 0.725), Q + M (mean AUC = 0.674, 95% CI = 0.623 – 0.725), and Q + C (mean AUC = 0.674, 95% CI = 0.639 – 0.709) all with Boruta selected variables (Fig. 3B), performed only slightly worse than the best model (Boruta – Q + C + M). Overall back shape and function dataset using Boruta selected variables produces the worst classification performance (Fig. 3A, AUC = 0.569, 95% CI = 0.538 – 0.60). Additionally, the dual modalities continuing back shape and function data always performed worse than those without (Fig. 3B), when using both all and Boruta selected variables. Classification performance metrics across all models is provided in Supplementary Table S7.

**Figure 3.**
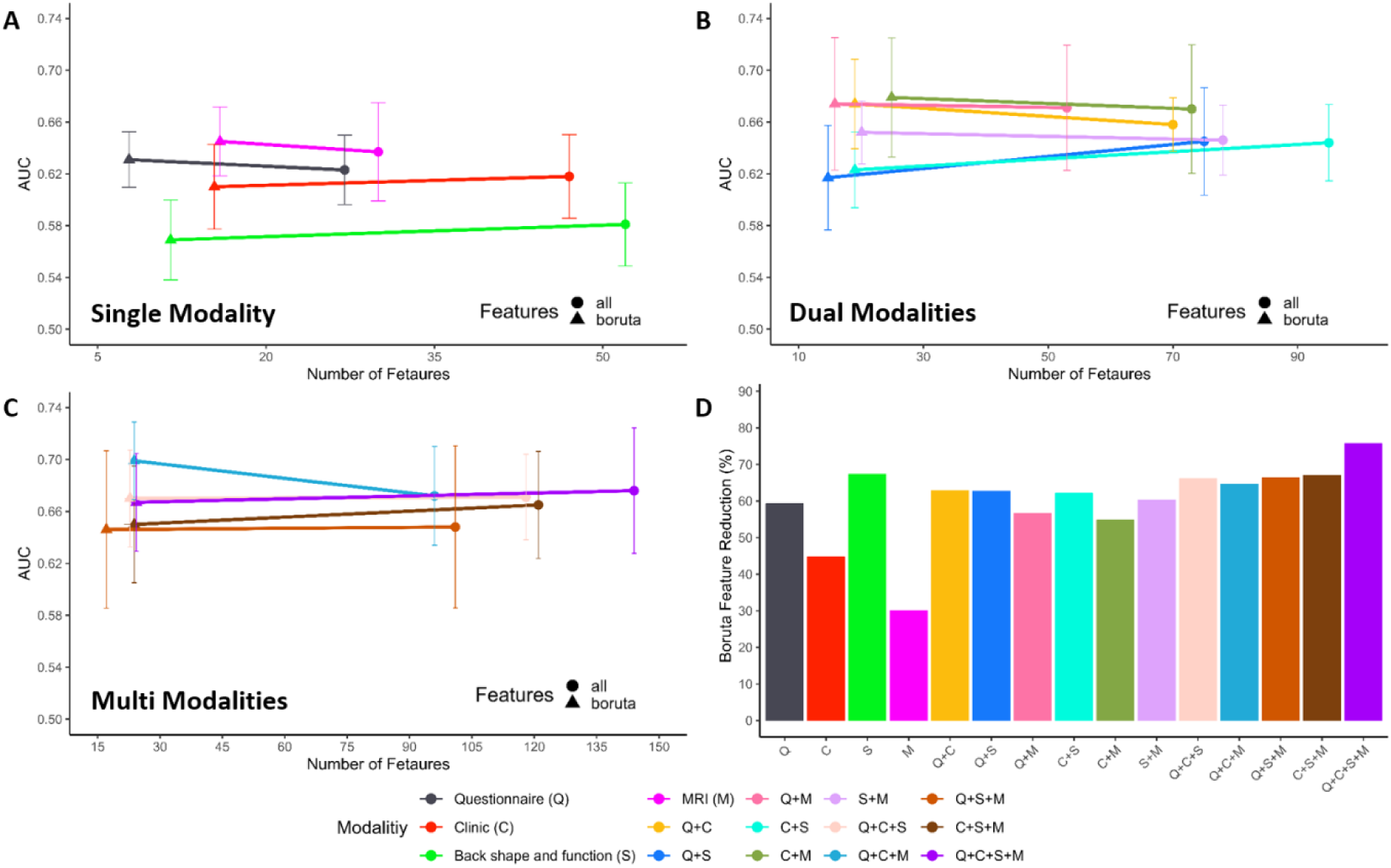
Boruta variable reduction performance. A – C shows RF classification model performance (AUC) following the reduction of variables using Boruta and all variables in the single, dual, and multi data modalities respectively. This shows the change in performance follow variable reduction. Error bars represent 95% CI in AUC over 10-fold train-test splits. D – Shows the amount of variable reduction by using Boruta as a percentage of the total number of variables within each modality dataset.

### Boruta variable importance

Boruta variable importance selection resulted in a median of 62.7% reduction in the number of variables across all 15 datasets (Fig. 3D). This large reduction in variables from Boruta performed comparable (eight slightly worse and seven better) compared to using all variables, with high overlap in confidence intervals. Furthermore, three of the top five performing modality models where those employing Boruta selected variables (Supplementary Table S22). The greatest performance improvement following Boruta variable selection was found in the Q + C + M datasets with an AUC increase of 0.270 and an average reduction of 72.2 variables (Fig. 3C). The smallest variable reduction was shown in MRI (30%) with an AUC increase of 0.008, while the largest variable reduction was found in the whole dataset (Q + C + S + M) with a reduction of 75.7% of the variables and an AUC decrease of 0.009.

Boruta selected variables represented the best patient delineation performance in single, dual, and multi-modality datasets (Fig. 3A-C). MRI was found to be the best single modality model using Boruta selected variables (Fig. 3A). The most important variables (Supplementary Table S27) were intervertebral disc (IVD) herniation L4 – L5, spinal canal width L2, IVD degeneration L3 – L4 and L4 – L5, and spinal canal width L1, showing a mean importance across the ten iterations of 19.49, 9.94, 9.46, 9.14, and 8.13 respectively. The best dual modalities cLBP patient stratification model, Boruta selected C + M (Fig. 3B), showed the second highest AUC (mean = 0.679). The MRI variable IVD herniation L4 – L5 was found to be the most important (mean = 13.72), with clinical mobility assessments of the hip, cervical spine, and the whole body showing high importance (Supplementary Table S31). The most important and robust variables of the best performing model (Boruta – Q + C + M, Fig. 3C) contained assessments from all three modalities (Supplementary Table S34). The Short-form 36 Health Status Questionnaires (SF-36) psychological well-being, SF-36 social function, and hip pain presented a mean importance of 13.36, 13.07, and 9.71 respectively. The clinical assessment, cervical axial rotation (left) and MRI variable IVD herniation L4 – L5 each showed an importance of 9.47 and 9.12. These represent the top five most important variables and the rest are provided in Table 3. The questionnaire Boruta reduced model provides a sparse model with decent performance, meaning the modality performs comparably well with a small amount of data. On average, the questionnaire Boruta model used 8 variables with a mean AUC reduction of 0.068 compared to the best model (Q + C + M). The most robust and important variables (Supplementary Table S39) were SF-36 social function (mean = 21.42), SF-36 psychological well-being (mean = 20.02), hip pain (mean =15.0), smoking in pack years (mean = 11.8), and family history of back pain (mean = 7.27). All Boruta selected important variables across the datasets are provided in Supplementary Table S24 – S38 and their correlation matrices in Supplementary Figure S7 – S21.

### Robust and important variables

To select the most robust and important variables for cLBP patient classification, the percentage of selection and average importance score was calculated across the 15 datasets (Supplementary Table S39). The variables that were selected at every opportunity (100%) across the multiple iterations and datasets were defined as most important and solely implemented in cLBP classification compared to all variables. This resulted in nine robust and important variables (Fig. 4A). The variables importance score is the Z-score of the mean decrease accuracy measure and is computed by dividing the average accuracy loss by its standard deviation. Meaning this value shows how much the models accuracy would decrease without this variable included. These variables represented psychosocial factors, IVD herniation and degeneration of the lower lumbar spine, presence of hip pain, as well as mobility of the neck and general mobility of the whole body. The performance of these nine variables were compared to all variables utilising the hold-out dataset (Fig. 1B) in a five-fold train-test workflow to provide an unbiased comparison. The best nine variables showed better mean accuracy (Fig. 4B, Boruta = 0.724, 95% CI = 0.593 – 0.855, All = 0.680, 95% CI = 0.625 – 0.736), AUC (Boruta = 0.664, 95% CI = 0.514 – 0.814, All = 0.602, 95% CI = 0.538 – 0.666), and sensitivity (Boruta = 0.442, 95% CI = 0.176 – 0.708, All = 0.298, 0.139 – 0.457), while all variables provided better specificity (Boruta = 0.892, 95% CI = 0.795 – 0.989, All = 0.902, 95% CI = 0.803 – 1.0). Moreover, all these nine variables showed significant univariate statistically significant differences between cLBP patients and asymptomatic controls in the questionnaire, clinical, and MRI datasets (Fig. 4A). Utilising a Wilcoxon-Mann-Whitney test revealed reduced scores of the SF36 for social function (u = 183751, z = -7.08, p < 0.001, effect size r = -.225) and psychological well-being (u = 178506.5, z = -7.74, p < 0.001, r = -0.247) in people suffering from cLBP. In addition, the occurrence of hip pain was also altered comparing people with and without cLBP (χ^2^ (3, N=986) = 40.17, p = 0.004, ω = 0.202). Comparing clinical examinations further revealed reduced cervical axial rotation to the left (u = 221595, z = -7.98, p < 0.001, r = -0.243), reduced sit to stand 30 second repetition (u = 226818.5, z = -6.83, p < 0.001, r = -0.208), as well as altered general mobility (χ2 (2, N=1080) = 46.81, p = 0.006, ω = 0.208) in cLBP patients. Regarding MRI investigations, increased IVD degeneration at L2-L3 (u = 147168, z = 3.38, p = 0.011, r = 0.120) and L4-L5 (u = 147691, z = 3.43, p = 0.010, r = 0.121), as well as increased disc herniation at L4-L5 (u = 153470.5, z = 6.02, p < 0.001, r = 0.213) were found in people suffering from cLBP compared to asymptomatic controls. Univariate statistical results for all variable can be found in Supplementary Table S40 – S47.

**Figure 4.**
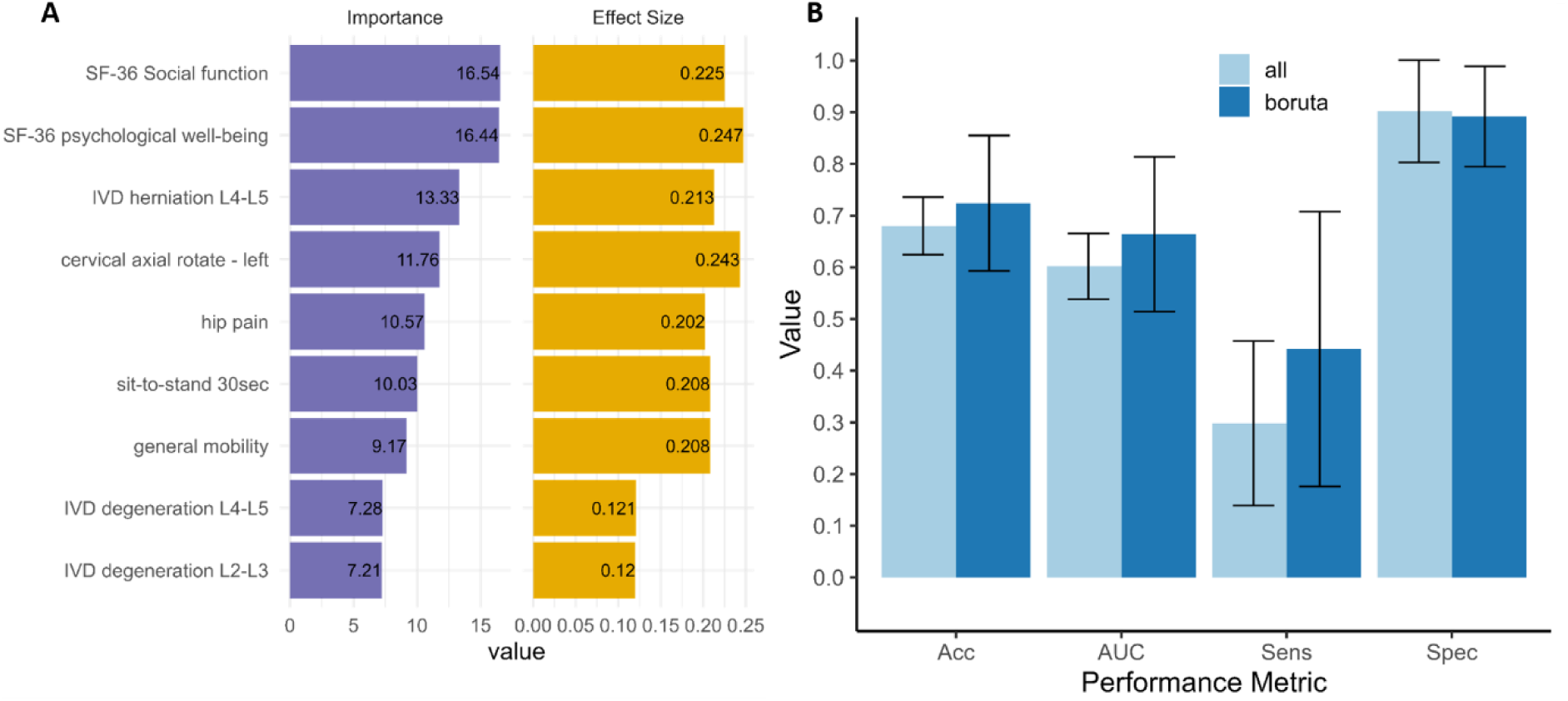
Most robust and important variables for chronic low back pain classification performance. A – Presents a bar plot of the nine most robust variables in order of average Boruta importance score (left). The right bar plot shows the absolute effect size (Cohen’s r or ω depending on data type) comparing controls and cLBP patients of the nine robust variables. B – Column plot showing RF classification performance as mean of five-fold train-test iterations in hold-out set using Boruta selected and all variables. Column plot error bars represent 95% CI. IVD – intervertebral disc, SF-36 – short form 36 health status questionnaire, Acc – accuracy, AUC – Area under the receiver operating characteristic curve, Sens – sensitivity, Spec – specificity.

## Discussion

This study employes a large multi-modal dataset and a machine learning workflow to demonstrate the importance of using data from different domains in cLBP patient delineation. Increasing the number of modalities generally lead to a model performance improvement although it seems the inclusion of back shape and motion data resulted in little to no performance improvement. Utilising Boruta in our iterative selection workflow resulted in considerable variable reduction across all datasets (median = 62.7%), while model performance remained comparable. This may reflect many variables showing little difference between patients and controls, or the underlying machanisms are more robustly captured by a small subset of variables. Both the best performing modality model (Q + C + M) and the most robust variables (Fig. 4), show the importance of measuring psychosocial factors, cervical axial rotation, general mobility, hip flexion, and lower lumbar spine disc degeneration and herniation ratings in cLBP patients.

The questionnaires probing the psychosocial factors, social function and psychological well-being, showed highest importance among all variables (Fig. 4A) and were the most important variables in the best sparse model (Boruta reduced questionnaire, Fig. 3A). Moreover, the quesionnaires represent the most cost effective modality to the examiner, highlighting the clinical importance of psychosocial factors in cLBP diagnosis and treatment.

Social functioning describes the ability of a person to engage in social activities, which we have shown to be an important marker for delineating cLBP patients from asymptomatic controls. Using cross-sectional data from 180 chronic low back pain patients, Ge and colleagues ^19^ showed that these patients reported more limitations in performing (major life tasks and) social activities as compared to subjects without cLBP, even after adjusting for influencing factors, such as socio-demographics, lifestyle and number of diseases. Furthermore, Tagliaferri et al. ^20^ were able to separate 4156 chronic back pain patients from the UK Biobank dataset into five sub-groups based on their scores of social isolation and depressive symptoms. Interestingly, increased social isolation was only a variable of three sub-groups, encompassing 26% of all back pain patients (n = 1085), while the remaining subgroups showed either no changes (4.1%; n = 776) or a reduction in social isolation scores (12%; n = 2296). This prevalence in patients with LBP may indicate that reduced social functioning was identified by some studies, while others did not find similar changes as compared to asymptomatic controls ^21^. However, as levels of social function (here: social participation) were found to be correlated with self-perceived physical health status ^22^, a direct impact of social functioning on personal functional impairments remains feasible.

In addition to social function, the psychological health or well-being was shown to be an important variable in cLBP patient delineation. Using longitudinal data from the SwePain cohort, including 9361 participants with and without chronic pain, psychological well-being scores at baseline were able to predict pain intensity after 2 years ^23^. Within this study, positive well-being was predictive of lower pain severity in participants without and with chronic pain. Similar conclusions were drawn from the comparison of back pain patients with different levels of mental distress, in which patients with higher mental distress showed, among other things, reduced psychological well-being and social function, and higher severe pain than patients with lower mental distress ^24^. Furthermore, patients suffering from chronic pain exhibit significantly lower quality of life scores across all sub-domains, including psychological well-being ^25^. Alterations in quality of life are stronger associated with changes in social functioning and psychological well-being (via pain catastrophizing) than pain intensity it self ^26^, indicating the high importance of psycho-social aspects for daily living with painful conditions such as back pain.

Our findings highlight the potential importance of psychosocial factors in cLBP and suggest a need for change in clinical practice. In Berlin, and more widely, the integration of psychosocial assessment into routine cLBP care could improve treatment effectiveness. This is consistent with previous meta-analytic evidence ^27^ supporting a combined biopsychosocial therapeutic approach to optimise treatment efficacy. Early identification of psychosocial barriers such as anxiety, depression or stress should be prioritised, with interventions such as cognitive behavioural therapy, stress management and mental health support used alongside traditional physical therapies.

We demonstrated that spinal herniation and degeneration observed on MRI may contribute to pain mechanisms in the bio-psychosocial model of cLBP, consistent with previous meta-analyses ^28^. However, it is clear that MRI findings alone do not fully explain pain presence in cLBP, highlighting the need for caution when interpreting MRI results at the individual patient level. Interestingly, cervical spine rotation but not lumbar back motion assessments were shown to be robust important examinations for cLBP delineation. This contradictory finding is likely a result of two factors. First, the poor cervical rotation may be the result of neck pain that has high comorbidity with cLBP ^29^ and can lead to decreased axial rotation ^30^. Second, poor psychological health has been associated with neck pain ^31^, which we found to also present high importance in patient stratification and relates to the bio-physical-psychosocial interplay present in cLBP patients ^5^. A systematic review on hip mobility in LBP patients ^32^ showed small to no changes in hip flexion compared to controls. As all studies had less than 110 subjects, they were likely under powered to uncover the decreased mobility we show here and may represent a diagnostic test for LBP. Previous studies have shown that clinical kinematic data can effectively stratify cLBP patients into high, low and intermediate risk groups ^33^, suggesting that pain correlates with reduced physical function. Persistent nociceptive input from aggravated spinal joints/muscles may lead to reduced motor output and spinal cord excitability^34^, potentially resulting in a reduced ability to recruit specific muscles and necessitating compensatory movement strategies. Our findings underscore that detailed movement analysis could serve as a diagnostic biomarker for LBP, potentially rivalling medical imaging in diagnostic accuracy and improving patient care by identifying sub-populations likely to respond well to specific therapies or at risk of adverse outcomes.

Classification of cLBP patients has often been conducted on relatively small sample sizes (< 200) as well as utilising a single data domain ^9^. Performance of such models are subject to overfitting due to their small samples and would likely perform poorly at out-of-sample classification in unseen external datasets ^35^. Furthermore, several studies have established classification models with high accuracy (> 0.8) at determining particular LBP symptoms ^36–43^, although these lack clinical applicability in understanding the most appropriate variables and modalities in classification of cLBP as well as the classification of the disorder in general. Classification models created using large datasets (n > 1000) either contained psychosocial and demographic variables, without imaging and physical variables ^44,45^. On the other hand, Jin-Heekun and colleagues ^46^ employed only physical variables without considering important psychosocial factors, which have shown to be important in previous research ^5,20,47,48^ as well as in our current study (Fig. 4). Our best model (Boruta – Q + C + M) performed slightly worse than Parsaeian et al. ^45^ (AUC 0.693 – 0.75) and compared to Shim et al. ^44^ (AUC, 0.693 – 0.716), we utilised more plentiful data points per subject in a significantly smaller sample size (approximately 34x and 6x smaller respectively) to address the clinically relevant question of what modalities and variables a best suited for cLBP classification.

The multimodal nature of the data utilised and the amount of subjects that have participated in physical, imaging, and questionnaire measurements are major strengths in our current study. Utilising the “Berlin Back Study” dataset that contains more than 500 subjects in the different domains highlighted as lacking in a recent review by our group ^9^, enabling us to robustly investigate the importance of different modalities as well as specific variables in cLBP patient stratification. The large sample size enabled us to minimise model overfitting through cross-validation and hold-out testing of good sample size and distributions. A more accurate representation of a models performance is provided by out-of-sample testing that uses a new sample population containing comparable variables. This provides a test set with minimised sampling and dataset bias greatly improving the generalisability and applicability of the findings.

The results of our study offer valuable insights into the potential of using a multimodal machine learning approach for the classification of individuals with chronic low back pain (cLBP). However, despite the promising nature of our dataset, the classification performance was moderate. One possible explanation is the size and diversity of the dataset. Although the sample size was substantial, it may not fully encompass the complex and multifactorial nature of cLBP, especially when considering the broad spectrum of psychosocial, environmental, socio-economic, and biological factors that contribute to the condition. Incorporating detailed information regarding socio-economic status, education, and other social factors could potentially improve model performance by capturing the broader context in which cLBP manifests.

While our study highlights the importance of psychosocial factors in the classification of cLBP, we acknowledge that the analysis of MRI data was limited. Several key MRI phenotypes, including disc bulging, spondylolisthesis, osteophytes, Modic lesions, endplate abnormalities, and high-intensity zones, were not included in the final analysis. It is also important to note that while psychosocial factors showed strong predictive power, the MRI analysis was not exhaustive, and a more complete MRI dataset would be necessary to make a fair comparison. Future studies should aim to expand both the MRI and psychosocial data to provide a more comprehensive understanding of the relative contributions of anatomical and psychosocial factors in cLBP.

A notable limitation of this study is the absence of key social determinants of health, such as income, education, and health insurance status, which are known to significantly influence both the risk of developing cLBP and its persistence. While we included job-related factors, such as posture during work, these social determinants were not available in our dataset. Incorporating these factors into future models could enhance the accuracy of our predictions and offer a more nuanced understanding of how socioeconomic and environmental factors intersect with clinical and psychosocial elements in the context of cLBP. The inclusion of such variables should be a priority in subsequent research to improve both model performance and clinical applicability.

We acknowledge the challenge of translating these findings into clinical practice. The complexity and costs associated with collecting such comprehensive data may not be feasible in all healthcare settings. However, the value of this research lies in its potential to inform the development of more personalized, evidence-based treatment strategies. In future work, we aim to explore how these multimodal data could contribute to treatment recommendations or patient phenotyping, providing clinicians with more precise tools to tailor interventions to individual patient needs. By advancing the field toward personalized care, we believe the clinical utility of this approach will become more apparent. Furthermore, our study provides a cross-sectional investigation of cLBP, whereas a prospective design would be better suited to examine the causality of the disorder. Two different types of interviews were conducted: face-to-face interviews (clinical examination) and electronic interviews (questionnaires). The main difference is that body language, facial expressions and other non-verbal social cues are obvious to the interviewer in face-to-face interviews, whereas these aspects are absent in electronic surveys. As both surveys have advantages and disadvantages, the answers of the study participants were weighted equally in this study. Additionally, the high number of questions could lead to a reduction in the participants’ attention and concentration.

In conclusion, while our current model shows promise, there is room for improvement. Expanding the dataset, incorporating more detailed and diverse data sources, and exploring alternative machine learning models are potential next steps that could enhance the predictive power of the system. A key future application will be the use of baseline data to predict treatment response and the transition from acute to chronic pain. This predictive capability could enable early identification of high-risk patients, improving treatment outcomes and advancing personalised medicine in the management of cLBP. We also believe that as more data becomes available and our understanding of the complex interplay of factors contributing to cLBP deepens, the clinical utility of such models will become more apparent, leading to better patient stratification and more personalized treatment approaches.

## Methods

### Quantitative variables and data collection

Patients had to meet all of the following inclusion criteria: written informed consent to participate in the study, asymptomatic (no back pain) or symptomatic (cLBP) caucasian women and men aged 18–67 years, pain duration ≥12 weeks daily (cLBP only), pain localization in the lumbopelvic region (cLBP only). A telephone interview was conducted during recruiting and subjects were excluded if they met any exclusion criteria. However, some subjects came to testing that should have been excluded during the telephone interview, and were then excluded at the testing site (Supplementary Table S3). No minimal threshold for LBP intensity was defined. A list of all variables including number of missing values is shown in Supplementary Table S19.

### Questionnaires

The localization, type, course, possible radiation, intensity, quality, duration, and any factors that may relieve or exacerbate the pain, as well as possible triggers or the patient’s own explanations regarding the cause of the pain has been asked. The patient’s medical history has been recorded, which includes any previous diseases and surgeries, and a detailed pain and general medication history as well as allergies, intolerances, and vaccination status among others. A family and social history (anamnesis) was taken (professional activity, family situation, diseases in the family, stressful situations, etc.). In addition, any past or present use of addictive substances has been asked (alcohol, nicotine, etc.).

The following questionnaires were completed within 30 minutes:

➤ Pain intensity, pain duration, and pain-related disability: von Korff et al. ^49^
➤ Disability Questionnaire: Roland and Morris (RMDQ) ^50^
➤ Short-form 36 Health Status Questionnaire: SF-36 ^51^. The following four domains were considered: general mental health (psychological distress and well-being), limitations in usual role activities because of emotional problems, vitality (energy and fatigue), and general health perceptions.
➤ International Physical Activity Questionnaire (IPAQ) ^52^
➤ Self-Report Behavioural Automaticity Index (SRBAI) ^53^
➤ Behavioural Regulation in Sport Questionnaire (BRSQ) ^54^
➤ Tampa Scale for Kinesiophobia (TSK-GV) ^55^.
➤ Fear-Avoidance Belief Questionnaire (FABQ) ^56^.

The participants primarily answered the questionnaires in digital form using a survey program specially developed for the study. The data were collected under similar conditions (e.g., same room, same computer) for all subjects at the study centre.

### Demographic data

During the clinical assessment, age, sex, body height, body weight, hip diameter, and waist diameter of the subjects were recorded. BMI was chosen instead of waist hip ratio to measure physical body size and health, as the BMI variable contained less missing values compared to waist hip ratio (Supplementary Table S19).

### Clinical examination

The clinical examination included the evaluation of organ functions (inspection, palpation, percussion, and auscultation), the general impression, and the vital parameters of the patient (temperature, heart rate, blood pressure, etc.). Examination was performed by an experienced orthopaedic consultant. The neurological status was assessed by the examination of the coordination, reflexes, sensitivity, and motor function. The evaluation of the functional parameters, that is, the assessment of posture, shape, orientation, and movement of the lumbar spine and pelvis, was based on current clinical standards (e.g., Ott and Schober test, 3-step hyperextension test, passive lumbar extension test, etc.) and self-assessment by the persons investigated. Data were documented according to their dimension using distances in cm, degrees of angle, and number of repetitions per defined time interval or bivalent whether pain provocation occurred. Self-assessment of functional restrictions of the back was recorded according to a scale from 1 (best) – 10 (worst).

### Back shape and function

All study participants received measurements of the back shape in the sagittal and frontal planes during upright standing and sitting using the Idiag M360 (Idiag AG, Fehraltorf, Switzerland). The device measures segmental angles of the thoracic and lumbar spine. In both postures, study participants were measured upright, in flexed, extended, and in left and right lateral bending (3 repetitions, ∼10 sec each). Maximum upper body flexion, extension, as well as left and right lateral bending were performed with extended knees. During extension the arms were crossed in front of the body. The order of performed tasks was randomised. The measurements were performed by trained medical students. The validity and reliability were demonstrated in previous studies ^57–60^.

### Spino-pelvic MRI

MRIs were conducted using a 1.5 MRI scanner. Following sequences were evaluated: 1) Sag T1 (4 mm slices), 2) Sag T2 (4 mm slices), 3) Cor STIR-T2 (4 mm slices) and 4) Axial T2 (3 mm slices). MRIs were evaluated for intervertebral disc degeneration (Pfirrmann classification ^61^), disc herniation (Kramer classification ^62^), facet joint arthrosis (Fujiwara classification ^63^), osteochondrosis intervertebralis ^64^, spondylolisthesis (Meyerding classification ^65^ and spinal canal stenosis (Schizas classification ^66^) at each level of the lumbar spine. The spino-pelvic MRI evaluation was performed blinded by two spine surgeons and a radiologist, all of whom have many years of experience in the evaluation of spinal pathologies. The inter-rater reliability was good-to-excellent for all measurement parameters.

### Data storage

All data files electronically recorded during the study period were stored on a database server folder (SharePoint folder) hosted by Charite-Universitaetsklinikum. A data back-up for the database is run daily. Local study team members signed a non-disclosure agreement. They have access to the database using a personal password and are authorized only for entries depending on their function based on a role concept (investigator, statistician, monitor, administrator etc.). A multilevel data validation plan was developed to guarantee the correctness and consistency of the data. Data were entered only after a check for completeness and plausibility. Furthermore, data were cross-checked for plausibility with previously entered data for each participant. Questionnaires filled out on paper are stored in a lockable cabinet at the university.

### Potential sources of bias and minimisation

To generally reduce the possible location and assessor bias, measurements (clinical physical and questionnaire assessment and back shape and function) were administered by a few trained clinicians in the same room with the same lighting. The self-administered questionnaires (von Korff, RMDQ, SF-36, IPAQ, SRBAI, BRSQ, TSK-GV, and FABQ) were completed by the subjects under supervision by our trained study coordinator who provided explanations for unclear questions and mitigated possible lack of motivation to complete the questions by assuring the subjects of the importance to complete the questionnaires. Furthermore, generic questionnaires (SF-36 or IPAQ) were placed before specific ones (SRBAI, BRSQ) to minimize bias from order effects.

To minimise the bias in our classification and variable selection modelling, as well as the modality comparison, variables directly assessing back pain and questions heavily biased to pain patients were removed. Such variables assessed back pain during particular movements or upon physical manipulation. Furthermore, questions only back pain patients were asked for example, pain intensity, duration, and disability, as well as pain and health biased self-questionnaires (von Korff, RMDQ, TSK-GV, FABQ, therapies, and the SF-36 sub-categories regarding physical function, physical role function, physical pain, health perception, and vitality), and clinician administered questions regarding pain medication intake, previous spinal disorder diagnosis, and participants’ subjective physical health assessment were removed as well to reduce model bias.

### Outcome

The outcome target for our classification model is cLBP patients. All participants were assessed by a clinician and diagnosed as cLBP patient, asymptomatic control, or suffered from LBP in the past but not at present. We used the clinician diagnosis of either current cLBP patient or asymptomatic control as our two-class target outcome. The participants with LBP in the past were removed to enable better distinguishable groups for binary classification and variable importance selection.

### Data handling, preprocessing, cleaning and missing data

Total and sub-scores for the self-administered questionnaires were used after removal of pain biased questionnaires (see *Potential sources of bias and minimisation*). This includes the SF-36 ^51^, the IPAQ ^52^, the SRBAI ^53^, and the BRSQ ^54^. The SF-36 was used to collect statements related to the health domains ‘emotional role limitation, (three items) ‘social functioning’ (two items), and ‘mental health’ (five items). A scoring algorithm was used to convert the raw scores into these three domains. The scores were transformed to range from zero (worst possible health) and 100 (best possible health). The SRBAI and BRSQ collected ratings for multiple statements on a numerical scale from 1 (strongly disagree) to 6 (strongly agree). For the SRBAI, the total score was calculated by summing the numerical values across all 4 statements. In contrast, sub-scores were created for the BRSQ that related to ‘intrinsic motivation’, ‘integrated regulation’ and ‘external regulation’. These scores were calculated by summing the numerical values across two statements per sub-score. The IPAQ, recorded the average time spent per day over the past 7 days while ‘sitting’, ‘walking’, doing ‘moderate activities’ (e.g., heavy lifting, digging, aerobics, or fast bicycling), and ‘vigorous activities’ (e.g. carrying light loads, bicycling at a regular pace, or doubles tennis). To estimate the energy requirements for each activity type, the average time spent per day in minutes was multiplied by MET-score (metabolic equivalents) of 1.5, 3.3, 4, or 8 for sitting, walking, moderate and vigorous activities, respectively. This resulted in MET-minutes scores, describing the amount of energy in kilocalories required for a 60 kilogram person. Finally, the MET-minutes scores were summed across ‘sitting’, ‘walking’, ‘moderate activities’ and ‘vigorous activities’, resulting in a total MET-minutes score per subject.

Modelling preprocessing was conducted by checking variables within each modality for very low variance and collinearity. Variables with close to zero (variance < 1) variance were removed. Furthermore, variables presenting a Spearman correlation greater than 0.9 were also removed to reduce collinearity between variables. The decision on which of the correlated variables to removed, was the variable showing less correlation to the target, cLBP patient status. Following these cleaning steps, single data modalities were joined with demographic data and subsequently joined with other datasets for the dual-and multi-modalities datasets. Subjects having any missing values were removed from our main analyses and MissForest ^18^ imputation was additionally conducted. Imputation was computed for the training and testing samples within each fold independently to avoid data leakage. Following cleaning and preprocessing modality dataset presented different number of subjects and variables with a similar age and sex distribution across cLBP patients and asymptomatic controls (Fig. 1A).

### Univariate statistics

The univariate statistics were carried out separately for continuous, ordinal and nominal data to compare patients suffering from cLBP against asymptomatic controls using R (version 4.3.1; www.r-project.org). As most continuous variables did not follow a normal distribution according to the Anderson-Darling test ^67^, we implemented the non-parametric Wilcoxon-Mann-Whitney test to determine significant difference between cLBP patients and asymptomatic controls for ordinal and continuous data. Hence, u-values, z-values, r-value (effect sizes), as well as the p-value are reported from. Nominal data were compared using the Chi-Square test and reported with Chi^2^ values, Cohen’s ω-values (effect size), and p-values. Statistical significance was determined at p≤0.05 following family wise error (FWE) ^68^ correction for multiple comparisons within each modality (demographics, questionnaires, clinical examinations, superficial spine morphology and motion, and spino-pelvic MRI).

### Machine learning

#### Boruta variable selection

We used the Boruta method ^16^ for importance variable selection. Boruta utilises random forest (RF) classification algorithm ^17^ with both the real variables and set of ‘dummy’ or ‘shadow’ variables, that are created by shuffling the variable values. This creates random variables (dummy features) that have the same distribution as the original variables, although represent the classification accuracy of this variable randomly sampled. As these dummy variables represent random noise, they had their possible correlation to the target (cLBP) removed. All real and dummy variables are used to classify cLBP and variable importance is calculated. Variable importance is calculated as the Z-score of the mean decrease in classification accuracy following the removal of this variable from the model. The importance of the dummy variables can be used as a reference to test the variable importance of the real variables. Through an iterative process, real variables that have significantly greater importance than the maximum dummy variable importance are marked as important. The variables that have a significantly lower importance than the maximum dummy variables are deemed unimportant and removed for the next iteration of selection. Iterations are repeated until the importance is assigned to all variables or a user defined iteration number is reached. We used a max of 2000 iterations with a random forest containing 1000 trees. As there are occasionally a few variables not definitively identified as important or removed after 2000 iterations by Boruta, we only selected the variables that have been confirmed as important in our subsequent modelling and analyses.

#### Random forest classification algorithm

We utilised RF ^17^ implemented using the ranger package ^69^ in R (version 4.3.1, www.r-project.org) to classify cLBP patients and pain-free controls. As RF provides variable importance measures and can handle categorical, ordinal, and continuous data it represents the ideal choice to deal with the different data types present in the Berlin Back dataset. Ten-fold cross validation was conducted during model training and hyper-parameter tuning utilising 1000 tree RF. We conducted hyper-parameter tuning using a tuning parameter search grid, which contained number of variables to be sampled at each split (mtry) of 1 – square root of the number of variables, and a minimum node size of 5 and 10. Therefore, using a gini split rule a grid search was conducted with all combination of hyper-parameters to determine the best. A ten-fold train-test loop (Fig. 1B) was conducted to determine the cLBP classification performance within each of the 15 datasets utilising all and Boruta selected variables independently. Model performance was calculated as follows; accuracy = (TP + TN) / (TP + FP + TN + FN), sensitivity = TP / (TP + FN), specificity = TN / (TN + FP), and AUC (area under the receiver operating characteristic (ROC) curve). Whereby, TP – true positive, TN – true negative, FP – false positive, and FN – false negative. The ROC curve represents the classification performance measured by sensitivity and specificity over a range [0, 1] of classification thresholds.

#### Robust variable selection workflow

Selecting important variables and comparing the 15 different dataset modalities was conducted on 90% (n = 1045, 19 – 72 y/o, mean age = 41.71 ± 12.28, cLBP = 469) of the preprocessed Berlin Back study dataset, with a 10% (n = 116, 20 – 64 y/o, mean age = 43.81 ± 12.12, cLBP = 43) hold-out set used to evaluate the most robust and important variables (Fig. 1B). The hold-out sample comprised of participants that had data for all variables. Each 15 modality datasets went through the iterative ten-fold train-test loop, where Boruta variable selection was conducted on the training set. Followed by RF training using all and Boruta selected variables implemented on the training dataset. Model performance was calculated using the test set. Across the ten loops, the percentage a variable was selected and average importance value was calculated across all 15 datasets. Finally, the variables that were selection within every train-test loop and within every possible dataset were provided as the most robust and important variables for cLBP classification. The demographic variables can be selected a maximum of 15 times, while the modality-specific variables a maximum of 8 times across the 15 dataset modalities. These variables were then used in a five-fold train test loop on the hold-out dataset and compared to a model trained using all variables.

## Data Availability

The raw data of this study will be openly released from the Berlin Back Study as per agreement with the funding agency following the completion of the data acquisition (30.12.2025).

## Disclosures

### Funding

This study is part of the Research Unit FOR 5177 funded by the German Research Foundation (DFG), Hendrik Schmidt: SCHM 2572/11-1, SCHM 2572/12-1, SCHM 2572/13-1; Sandra Reitmaeier: RE 4292/3-1, Matthias Pumberger: PU762/1-1. The analyses and contribution from the Hochschule für Gesundheit were funded, in part, by grant number 50WK2273A (to DLB) from the German AeroSpace Center (DLR).

### Conflicts of interest

All authors declare no conflict of interests.

## Acknowledgments

We would like to thank all patients and healthy participants for their selfless participation in this study and the participating companies for informing their employees about this study.

## Data and code availability

All results in this study are provided in the (Supplementary) tables. The Berlin Back study is currently ongoing (end date 31/12/2025) and therefore the raw data used in this manuscript cannot be provided. The raw data will be openly released from the Berlin Back Study as per agreement with the funding agency following the completion of the data acquisition. A link to the raw data will be provided on the Github repository where the analysis code is located (https://github.com/viko18/BerlinBack_FeatImp/) when it is made available.

## Author Contributions

**CRediT Contributions:**

Conceptualization – SV, HS, DLB

Methodology – SV, HS, DLB, MA, CK

Software – SV, FJ

Validation – SV

Formal Analysis – SV, FJ

Investigation – SV, LAB, NT, MP, SR

Resources – HS, DLB

Data Curation – SV FJ

Writing original draft – SV, RD, FJ, DLB, HS

Writing review & editing – All co-authors

Visualizations – SV, FJ

Supervision – DLB, HS, MA, CK

Project Administration – HS, SV

Funding Acquisition – HS, DLB

## Notes

### Competing Interest Statement

The authors have declared no competing interest.

### Clinical Trial

DRKS00027907

### Author Declarations

The ethics committee of the Charite Universitaetsmedizin Berlin (registry numbers: EA4/011/10, EA1/162/13) gave ethical approval for this work.

### Summary of Updates

- Added extra demographic data tables and figure 2 - computed imputation using MissForest - Added extra limitations and discussion paragraphs - fixed some uncommon language and shortened introduction

## References

1. Andersson, G. B. Epidemiologic aspects on low-back pain in industry. Spine (Phila Pa 1976) 6, 53–60 (1981).

2. Meucci, R. D., Fassa, A. G. & Faria, N. M. X. Prevalence of chronic low back pain: systematic review. Rev Saude Publica 49, 1 (2015).

3. Pastorino, R. et al. Benefits and challenges of Big Data in healthcare: an overview of the European initiatives. Eur J Public Health 29, 23–27 (2019).

4. Shilo, S., Rossman, H. & Segal, E. Axes of a revolution: challenges and promises of big data in healthcare. Nat Med 26, 29–38 (2020).

5. Tagliaferri, S. D. et al. Relative contributions of the nervous system, spinal tissue and psychosocial health to non-specific low back pain: Multivariate meta-analysis. Eur J Pain (2021) doi:10.1002/ejp.1883.

6. Grotle, M. et al. Lumbar spine surgery across 15 years: trends, complications and reoperations in a longitudinal observational study from Norway. BMJ Open 9, e028743 (2019).

7. Lee, W. et al. Identifying and Assessing Interesting Subgroups in a Heterogeneous Population. Biomed Res Int 2015, 462549 (2015).

8. Lötsch, J. & Ultsch, A. Machine learning in pain research. Pain 159, 623–630 (2018).

9. Tagliaferri, S. D. et al. Artificial intelligence to improve back pain outcomes and lessons learnt from clinical classification approaches: three systematic reviews. npj Digital Medicine 3, 93 (2020).

10. Noroozi, Z., Orooji, A. & Erfannia, L. Analyzing the impact of feature selection methods on machine learning algorithms for heart disease prediction. Sci Rep 13, 22588 (2023).

11. Mwangi, B., Tian, T. S. & Soares, J. C. A review of feature reduction techniques in neuroimaging. Neuroinformatics 12, 229–244 (2014).

12. World Medical Association. World Medical Association Declaration of Helsinki: ethical principles for medical research involving human subjects. JAMA 310, 2191–2194 (2013).

13. von Elm, E. et al. The Strengthening the Reporting of Observational Studies in Epidemiology (STROBE) Statement: Guidelines for reporting observational studies. Preventive Medicine 45, 247–251 (2007).

14. Collins, G. S., Reitsma, J. B., Altman, D. G. & Moons, K. G. M. Transparent Reporting of a multivariable prediction model for Individual Prognosis or Diagnosis (TRIPOD): the TRIPOD statement. Ann Intern Med 162, 55–63 (2015).

15. Shadbahr, T. et al. The impact of imputation quality on machine learning classifiers for datasets with missing values. Commun Med 3, 1–15 (2023).

16. Kursa, M. B. & Rudnicki, W. R. Feature Selection with the Boruta Package. Journal of Statistical Software 36, 1–13 (2010).

17. Breiman, L. Random Forests. Machine Learning 45, 5–32 (2001).

18. Stekhoven, D. J. & Bühlmann, P. MissForest—non-parametric missing value imputation for mixed-type data. Bioinformatics 28, 112–118 (2012).

19. Ge, L., Pereira, M. J., Yap, C. W. & Heng, B. H. Chronic low back pain and its impact on physical function, mental health, and health-related quality of life: a cross-sectional study in Singapore. Sci Rep 12, 20040 (2022).

20. Tagliaferri, S. D. et al. Chronic back pain sub-grouped via psychosocial, brain and physical factors using machine learning. Sci Rep 12, 15194 (2022).

21. Iguti, A. M., Guimarães, M. & Barros, M. B. A. Health-related quality of life (SF-36) in back pain: a population-based study, Campinas, São Paulo State, Brazil. Cad Saude Publica 37, e00206019 (2021).

22. Takeyachi, Y. et al. Correlation of low back pain with functional status, general health perception, social participation, subjective happiness, and patient satisfaction. Spine (Phila Pa 1976) 28, 1461–1466; discussion 1467 (2003).

23. Larsson, B., Dragioti, E., Gerdle, B. & Björk, J. Positive psychological well-being predicts lower severe pain in the general population: a 2-year follow-up study of the SwePain cohort. Ann Gen Psychiatry 18, 8 (2019).

24. Hnatešen, D. et al. Quality of Life and Mental Distress in Patients with Chronic Low Back Pain: A Cross-Sectional Study. Int J Environ Res Public Health 19, 10657 (2022).

25. Hadi, M. A., McHugh, G. A. & Closs, S. J. Impact of Chronic Pain on Patients’ Quality of Life: A Comparative Mixed-Methods Study. J Patient Exp 6, 133–141 (2019).

26. Lamé, I. E., Peters, M. L., Vlaeyen, J. W. S., Kleef, M. v & Patijn, J. Quality of life in chronic pain is more associated with beliefs about pain, than with pain intensity. Eur J Pain 9, 15–24 (2005).

27. Kamper, S. J. et al. Multidisciplinary biopsychosocial rehabilitation for chronic low back pain: Cochrane systematic review and meta-analysis. BMJ 350, h444 (2015).

28. Brinjikji, W. et al. MRI Findings of Disc Degeneration are More Prevalent in Adults with Low Back Pain than in Asymptomatic Controls: A Systematic Review and Meta-Analysis. AJNR Am J Neuroradiol 36, 2394–2399 (2015).

29. von der Lippe, E., et al. Prevalence of back and neck pain in Germany. Results from the BURDEN 2020 Burden of Disease Study. J Health Monit 6, 2–14 (2021).

30. Rampazo, É. P. et al. Sensory, Motor, and Psychosocial Characteristics of Individuals With Chronic Neck Pain: A Case Control Study. Physical Therapy 101, pzab104 (2021).

31. Mansfield, M. et al. The association between psychosocial factors and mental health symptoms in cervical spine pain with or without radiculopathy on health outcomes: a systematic review. BMC Musculoskeletal Disorders 24, 235 (2023).

32. Avman, M. A., Osmotherly, P. G., Snodgrass, S. & Rivett, D. A. Is there an association between hip range of motion and nonspecific low back pain? A systematic review. Musculoskeletal Science and Practice 42, 38–51 (2019).

33. Abdollahi, M. et al. Using a Motion Sensor to Categorize Nonspecific Low Back Pain Patients: A Machine Learning Approach. Sensors 20, 3600 (2020).

34. Nijs, J. et al. Nociception affects motor output: a review on sensory-motor interaction with focus on clinical implications. Clin J Pain 28, 175–181 (2012).

35. Rajput, D., Wang, W.-J. & Chen, C.-C. Evaluation of a decided sample size in machine learning applications. BMC Bioinformatics 24, 48 (2023).

36. Al Imran, A., Rifat, M. R. I. & Mohammad, R. Enhancing the classification performance of lower back pain symptoms using genetic algorithm-based feature selection. in 455–469 (Springer, 2020).

37. Abdullah, A. A., Yaakob, A. & Ibrahim, Z. Prediction of Spinal Abnormalities Using Machine Learning Techniques. in 1–6 (IEEE, 2018).

38. Riveros, N. A. M., Espitia, B. A. C. & Pico, L. E. A. Comparison between K-means and self-organizing maps algorithms used for diagnosis spinal column patients. Informatics in Medicine Unlocked 16, 100206 (2019).

39. Sandag, G. A., Tedry, N. E. & Lolong, S. Classification of lower back pain using K-Nearest Neighbor algorithm. in 1–5 (IEEE, 2018).

40. Karabulut, E. M. & Ibrikci, T. Effective automated prediction of vertebral column pathologies based on logistic model tree with SMOTE preprocessing. Journal of Medical Systems 38, 50 (2014).

41. Mathew, B., Norris, D., Hendry, D. & Waddell, G. Artificial intelligence in the diagnosis of low-back pain and sciatica. Spine 13, 168–172 (1988).

42. Vaughn, M. L., Cavill, S. J., Taylor, S. J., Foy, M. A. & Fogg, A. J. Direct explanations for the development and use of a multi-layer perceptron network that classifies low-back-pain patients. International Journal of Neural Systems 11, 335–347 (2001).

43. Zhang, W., et al. Deep learning-based detection and classification of lumbar disc herniation on magnetic resonance images. JOR SPINE 6, e1276 (2023).

44. Shim, J.-G. et al. Machine Learning Approaches to Predict Chronic Lower Back Pain in People Aged over 50 Years. Medicina 57, 1230 (2021).

45. Parsaeian, M., Mohammad, K., Mahmoudi, M. & Zeraati, H. Comparison of logistic regression and artificial neural network in low back pain prediction: second national health survey. Iranian Journal of Public Health 41, 86 (2012).

46. Jin-Heeku. Analysis of sitting posture using wearable sensor data and support vector machine model. Medico-Legal Update 1, 334–338 (2018).

47. Tagliaferri, S. D. et al. Brain structure, psychosocial, and physical health in acute and chronic back pain: a UKBioBank study. Pain 163, 1277–1290 (2022).

48. Tagliaferri, S. D. et al. Towards data-driven biopsychosocial classification of non-specific chronic low back pain: a pilot study. Sci Rep 13, 13112 (2023).

49. Von Korff, M., Ormel, J., Keefe, F. J. & Dworkin, S. F. Grading the severity of chronic pain. PAIN 50, 133 (1992).

50. Roland, M. & Fairbank, J. The Roland–Morris Disability Questionnaire and the Oswestry Disability Questionnaire. Spine 25, 3115 (2000).

51. Ware, J. E. & Sherbourne, C. D. The MOS 36-item short-form health survey (SF-36). I. Conceptual framework and item selection. Med Care 30, 473–483 (1992).

52. Craig, C. L. et al. International Physical Activity Questionnaire: 12-Country Reliability and Validity. Medicine & Science in Sports & Exercise 35, 1381 (2003).

53. Gardner, B., Abraham, C., Lally, P. & de Bruijn, G.-J. Towards parsimony in habit measurement: Testing the convergent and predictive validity of an automaticity subscale of the Self-Report Habit Index. International Journal of Behavioral Nutrition and Physical Activity 9, 102 (2012).

54. Lonsdale, C., Hodge, K. & Rose, E. A. The behavioral regulation in sport questionnaire (BRSQ): Instrument development and initial validity evidence. Journal of Sport & Exercise Psychology 30, 323–355 (2008).

55. Rusu, A. C., Kreddig, N., Hallner, D., Hülsebusch, J. & Hasenbring, M. I. Fear of movement/(Re)injury in low back pain: confirmatory validation of a German version of the Tampa Scale for Kinesiophobia. BMC Musculoskelet Disord 15, 280 (2014).

56. Waddell, G., Newton, M., Henderson, I., Somerville, D. & Main, C. J. A Fear-Avoidance Beliefs Questionnaire (FABQ) and the role of fear-avoidance beliefs in chronic low back pain and disability. PAIN 52, 157 (1993).

57. Dreischarf, B. et al. Comparison of three validated systems to analyse spinal shape and motion. Sci Rep 12, 10222 (2022).

58. Guermazi, M. et al. [Validity and reliability of Spinal Mouse to assess lumbar flexion]. Ann Readapt Med Phys 49, 172–177 (2006).

59. Topalidou, A., Tzagarakis, G., Souvatzis, X., Kontakis, G. & Katonis, P. Evaluation of the reliability of a new non-invasive method for assessing the functionality and mobility of the spine. Acta Bioeng Biomech 16, 117–124 (2014).

60. Barrett, E., McCreesh, K. & Lewis, J. Reliability and validity of non-radiographic methods of thoracic kyphosis measurement: a systematic review. Man Ther 19, 10–17 (2014).

61. Pfirrmann, C. W., Metzdorf, A., Zanetti, M., Hodler, J. & Boos, N. Magnetic resonance classification of lumbar intervertebral disc degeneration. Spine 26, 1873–8 (2001).

62. Kraemer, J. Natural course and prognosis of intervertebral disc diseases. International Society for the Study of the Lumbar Spine Seattle, Washington, June 1994. Spine (Phila Pa 1976) 20, 635–639 (1995).

63. Fujiwara, A. et al. The relationship between facet joint osteoarthritis and disc degeneration of the lumbar spine: an MRI study. Eur Spine J 8, 396–401 (1999).

64. Modic, M. T., Steinberg, P. M., Ross, J. S., Masaryk, T. J. & Carter, J. R. Degenerative disk disease: assessment of changes in vertebral body marrow with MR imaging. Radiology 166, 193–199 (1988).

65. Meyerding, H. W. Spondyloptosis. Surgery, Gynecology & Obstetrics 371–377 (1932).

66. Schizas, C. et al. Qualitative grading of severity of lumbar spinal stenosis based on the morphology of the dural sac on magnetic resonance images. Spine (Phila Pa 1976) 35, 1919–1924 (2010).

67. Anderson, T. W. & Darling, D. A. Asymptotic Theory of Certain ‘Goodness of Fit’ Criteria Based on Stochastic Processes. The Annals of Mathematical Statistics 23, 193– 212 (1952).

68. Holm, S. A Simple Sequentially Rejective Multiple Test Procedure. Scandinavian Journal of Statistics 6, 65–70 (1979).

69. Wright, M. N. & Ziegler, A. ranger: A Fast Implementation of Random Forests for High Dimensional Data in C++ and R. Journal of Statistical Software 77, 1–17 (2017).

