## Supplementary for "Integrating Multidimensional Data Analytics for Precision Diagnosis of Chronic Low Back Pain"

Supplementary Figure S13. Spearman correlation matrix of Boruta selected important features for questionnaire + MRI dataset ... 76
Supplementary Table S30. Boruta selected important features for MRI and clinical assessment modality dataset ... 76
Supplementary Figure S14. Spearman correlation matrix of Boruta selected important features for clinical assessment + MRI dataset ... 78
Supplementary Table S31. Boruta selected important features for MRI and superficial spine morphology dataset ... 78
Supplementary Figure S15. Spearman correlation matrix of Boruta selected important features for back shape and function + MRI dataset ... 80
Supplementary Table S32. Boruta selected important features for questionnaire and superficial spine morphology dataset ... 80
Supplementary Figure S16. Spearman correlation matrix of Boruta selected important features for questionnaire + back shape and function dataset ... 82
Supplementary Table S33. Boruta selected important features for questionnaire clinical assessment, and MRI dataset ... 82
Supplementary Figure S17. Spearman correlation matrix of Boruta selected important features for questionnaire + clinical assessment + MRI dataset ... 84
Supplementary Table S34. Boruta selected important features for clinical assessment, MRI, and superficial spine morphology dataset ... 84
Supplementary Figure S18. Spearman correlation matrix of Boruta selected important features for MRI + clinical assessment + back shape and function dataset ... 86
Supplementary Table S35. Boruta selected important features for questionnaire clinical assessment, and superficial spine morphology dataset ... 86
Supplementary Figure S19. Spearman correlation matrix of Boruta selected important features for questionnaire + clinical assessment + back shape and function dataset ... 88
Supplementary Table S36. Boruta selected important features for questionnaire, superficial spine morphology, and MRI dataset ... 88
Supplementary Figure S20. Spearman correlation matrix of Boruta selected important features for MRI + questionnaire + back shape and function dataset ... 90
Supplementary Table S37. Boruta selected important features for clinical assessment, MRI, questionnaire, and superficial spine morphology dataset ... 90
Supplementary Figure S21. Spearman correlation matrix of Boruta selected important features for questionnaire + clinical assessment + back shape and function + MRI dataset ... 92
Supplementary Table S38. Overall feature importance and robustness across all modality datasets ... 92
**Supplementary Data 4: Univariate statistics** ... 95
Supplementary Table S39: Demographic Wilcoxon-Mann-Whitney test (continuous and ordinal) ... 95
Supplementary Table S40: Demographic Chi2 Test (nominal) ... 95
Supplementary Table S41: Questionnaire Wilcoxon-Mann-Whitney test (continuous and ordinal) ... 95
Supplementary Table S42: Questionnaire Chi2 Test (nominal) ... 96
Supplementary Table S43: Clinical Physcial Assessment Wilcoxon-Mann-Whitney test (continuous and ordinal) ... 96

Supplementary Table S44: Clinical Physcial Assessment Chi2 Test (nominal) ................. 97
Supplementary Table S45: Superficial spine morphology Wilcoxon-Mann-Whitney test (continuous) ........................................................................................................ 97
Supplementary Table S46: Spino-pelvic MRI Wilcoxon-Mann-Whitney test (continuous and ordinal) ........................................................................................................ 99

### Supplementary Data 1: STROBE & Tripod Checklist

Supplementary Table S1: STROBE Checklist.

| | Item No | Recommendation | Section or page number |
|---|---|---|---|
| **Title and abstract** | 1 | (*a*) Indicate the study's design with a commonly used term in the title or the abstract | 1 |
| | | (*b*) Provide in the abstract an informative and balanced summary of what was done and what was found | 2 |
| **Introduction** | | | |
| Background/rationale | 2 | Explain the scientific background and rationale for the investigation being reported | 3-4 |
| Objectives | 3 | State specific objectives, including any prespecified hypotheses | 5 |
| **Methods** | | | |
| Study design | 4 | Present key elements of study design early in the paper | 6 |
| Setting | 5 | Describe the setting, locations, and relevant dates, including periods of recruitment, exposure, follow-up, and data collection | 6 |
| Participants | 6 | (*a*) Give the eligibility criteria, and the sources and methods of selection of participants | 6-7 |
| Variables | 7 | Clearly define all outcomes, exposures, predictors, potential confounders, and effect modifiers. Give diagnostic criteria, if applicable | 7-12 |
| Data sources/ measurement | 8* | For each variable of interest, give sources of data and details of methods of assessment (measurement). Describe comparability of assessment methods if there is more than one group | 7-11 |
| Bias | 9 | Describe any efforts to address potential sources of bias | 11-12 |
| Study size | 10 | Explain how the study size was arrived at | - |
| Quantitative variables | 11 | Explain how quantitative variables were handled in the analyses. If applicable, describe which groupings were chosen and why | 7-11, 12-14 |
| Statistical methods | 12 | (*a*) Describe all statistical methods, including those used to control for confounding | 14 |
| | | (*b*) Describe any methods used to examine subgroups and interactions | 10-11, 13, 29-31 |
| | | (c) Explain how missing data were addressed | 7 |
| | | (*d*) If applicable, describe analytical methods taking account of sampling strategy | Not applicable |
| | | (*e*) Describe any sensitivity analyses | Not applicable |
| **Results** | | | |
| Participants | 13* | (a) Report numbers of individuals at each stage of study—eg numbers potentially eligible, examined for eligibility, confirmed eligible, included in the study, completing follow-up, and analysed | 7-8 |
| | | (b) Give reasons for non-participation at each stage | 7-8 |
| | | (c) Consider use of a flow diagram | - |
| Descriptive data | 14* | (a) Give characteristics of study participants (eg demographic, clinical, social) and information on exposures and potential confounders | 7-8, 12-14 |

| | | | |
|---|---|---|---|
| | | (b) Indicate number of participants with missing data for each variable of interest | 7-8 |
| Outcome data | 15* | Report numbers of outcome events or summary measures | 12 |
| Main results | 16 | (*a*) Give unadjusted estimates and, if applicable, confounder-adjusted estimates and their precision (eg, 95% confidence interval). Make clear which confounders were adjusted for and why they were included | 18-23 |
| | | (*b*) Report category boundaries when continuous variables were categorized | Not applicable |
| | | © If relevant, consider translating estimates of relative risk into absolute risk for a meaningful time period | Not applicable |
| Other analyses | 17 | Report other analyses done—eg analyses of subgroups and interactions, and sensitivity analyses | Not applicable |
| **Discussion** | | | |
| Key results | 18 | Summarise key results with reference to study objectives | 24 |
| Limitations | 19 | Discuss limitations of the study, taking into account sources of potential bias or imprecision. Discuss both direction and magnitude of any potential bias | 28 |
| Interpretation | 20 | Give a cautious overall interpretation of results considering objectives, limitations, multiplicity of analyses, results from similar studies, and other relevant evidence | 24-27 |
| Generalisability | 21 | Discuss the generalisability (external validity) of the study results | 25-28 |
| **Other information** | | | |
| Funding | 22 | Give the source of funding and the role of the funders for the present study and, if applicable, for the original study on which the present article is based | 6 |

#### Supplementary Table S2: TRIPOD Checklist

| Section/Topic | Item | Checklist Item | Page |
|---|---|---|---|
| **Title and abstract** | | | |
| Title | 1 | Identify the study as developing and/or validating a multivariable prediction model, the target population, and the outcome to be predicted. | 1 |
| Abstract | 2 | Provide a summary of objectives, study design, setting, participants, sample size, predictors, outcome, statistical analysis, results, and conclusions. | 2 |
| **Introduction** | | | |
| Background and objectives | 3a | Explain the medical context (including whether diagnostic or prognostic) and rationale for developing or validating the multivariable prediction model, including references to existing models. | 3-4 |
| | 3b | Specify the objectives, including whether the study describes the development or validation of the model or both. | 5 |
| **Methods** | | | |
| Source of data | 4a | Describe the study design or source of data (e.g., randomized trial, cohort, or registry data), separately for the development and validation data sets, if applicable. | 6 |
| | 4b | Specify the key study dates, including start of accrual; end of accrual; and, if applicable, end of follow-up. | 6 |
| Participants | 5a | Specify key elements of the study setting (e.g., primary care, secondary care, general population) including number and location of centres. | 6 |
| | 5b | Describe eligibility criteria for participants. | 6-7 |
| | 5c | Give details of treatments received, if relevant. | - |
| Outcome | 6a | Clearly define the outcome that is predicted by the prediction model, including how and when assessed. | 12 |
| | 6b | Report any actions to blind assessment of the outcome to be predicted. | - |
| | 7a | Clearly define all predictors used in developing or validating the multivariable | 7-10, 11- |

| | | | |
|---|---|---|---|
| Predictors | | prediction model, including how and when they were measured. | 14 |
| | 7b | Report any actions to blind assessment of predictors for the outcome and other predictors. | - |
| Sample size | 8 | Explain how the study size was arrived at. | - |
| Missing data | 9 | Describe how missing data were handled (e.g., complete-case analysis, single imputation, multiple imputation) with details of any imputation method. | 7 |
| Statistical analysis methods | 10a | Describe how predictors were handled in the analyses. | 12-14 |
| | 10b | Specify type of model, all model-building procedures (including any predictor selection), and method for internal validation. | 14-16 |
| | 10d | Specify all measures used to assess model performance and, if relevant, to compare multiple models. | 15-16 |
| Risk groups | 11 | Provide details on how risk groups were created, if done. | - |
| **Results** | | | |
| Participants | 13a | Describe the flow of participants through the study, including the number of participants with and without the outcome and, if applicable, a summary of the follow-up time. A diagram may be helpful. | 7-8 |
| | 13b | Describe the characteristics of the participants (basic demographics, clinical features, available predictors), including the number of participants with missing data for predictors and outcome. | 7-8 |
| Model development | 14a | Specify the number of participants and outcome events in each analysis. | 7, 12-14 |
| | 14b | If done, report the unadjusted association between each candidate predictor and outcome. | - |
| Model specification | 15a | Present the full prediction model to allow predictions for individuals (i.e., all regression coefficients, and model intercept or baseline survival at a given time point). | 18-23 |
| | 15b | Explain how to the use the prediction model. | - |
| Model performance | 16 | Report performance measures (with CIs) for the prediction model. | 18-23 |
| **Discussion** | | | |
| Limitations | 18 | Discuss any limitations of the study (such as nonrepresentative sample, few events per predictor, missing data). | 28 |
| Interpretation | 19b | Give an overall interpretation of the results, considering objectives, limitations, and results from similar studies, and other relevant evidence. | 24-27 |
| Implications | 20 | Discuss the potential clinical use of the model and implications for future research. | 24-27 |
| **Other information** | | | |
| Supplementary information | 21 | Provide information about the availability of supplementary resources, such as study protocol, Web calculator, and data sets. | Supplementary Data |
| Funding | 22 | Give the source of funding and the role of the funders for the present study. | 6 |

### Supplementary Data 2: Study variables, preprocessing, and cleaning

Supplementary Table S3. Exclusion criteria and amount of participants excluded after arriving for testing.

| Exclusion Critera | Number Excluded |
|---|---|
| Professional atheletes | 0 |
| Substance abuse | 0 |
| Pregnant | 0 |
| BMI >28 $kg/m^2$ | 0 |
| Central or peripheral neurologic impairments (e.g., spinal cord injury, radicular symptoms, sensory deficits) | 4 |
| Irritated, inflamed, and infected tissues in the measuring areas of the back | 2 |
| spinal fractures, osteoporosis, tumor diseases and bone metastases, and previous spinal surgery | 21 |
| strong drug therapy (opioids, muscle relaxants, antiepileptics), intake of long-acting antihistamines and/or systemic glucocorticoids, and/or immunosuppressive drugs | 0 |
| Rheumatic diseases and/or active systemic diseases (e.g., tuberculosis, collagenosis, multiple sclerosis, autoimmune diseases, acquired immune deficiency syndrome) | 0 |
| internal diseases that pose a potential risk to the study participants during the measurements (e.g., coronary heart diseases, chronic obstructive pulmonary disease) and/or influence the findings | 0 |
| heart insufficiency and/or malignant hypertension, | 0 |
| malpositions or anomalies of the lower extremities (e.g., knee or hip arthroplasty, arthrodesis) | 2 |

Supplementary Table S4. Demographic data of cleaned questionnaire dataset

| | Asymptomatic | Chronic low back pain |
|---|---|---|
| Sample size | 555 | 431 |
| Females | 299 | 253 |
| Age (years) mean [sd] | 40.45 [12.36] | 43.25 [11.54] |
| Body mass index mean [sd] | 23.56 [2.74] | 23.53 [2.83] |
| Pain duration (weeks) mean [sd] | | 563.67 [544.82] |
| Pain Intensity (VAS) mean [sd] | | 2.88 [1.85] |

Supplementary Table S5. Demographic data of cleaned clinical assessment dataset

| | Asymptomatic | Chronic low back pain |
|---|---|---|

| | | |
|---|---|---|
| Sample size | 596 | 484 |
| Females | 319 | 286 |
| Age (years)<br>mean [sd] | 40.5 [12.61] | 43.58 [11.67] |
| Body mass index<br>mean [sd] | 23.53 [2.74] | 23.57 [2.82] |
| Pain duration (weeks)<br>mean [sd] | | 557.49 [532.35] |
| Pain Intensity (VAS)<br>mean [sd] | | 2.87 [1.84] |

Supplementary Table S6. Demographic data of cleaned back shape and function dataset

| | **Asymptomatic** | **Chronic low back pain** |
|---|---|---|
| Sample size | 624 | 489 |
| Females | 337 | 299 |
| Age (years)<br>mean [sd] | 40.75 [12.59] | 43.68 [11.71] |
| Body mass index<br>mean [sd] | 23.53 [2.76] | 23.55 [2.83] |
| Pain duration (weeks)<br>mean [sd] | | 548.87 [527.87] |
| Pain Intensity (VAS)<br>mean [sd] | | 2.9 [1.86] |

Supplementary Table S7. Demographic data of cleaned MRI dataset

| | **Asymptomatic** | **Chronic low back pain** |
|---|---|---|
| Sample size | 456 | 343 |
| Females | 254 | 206 |
| Age (years)<br>mean [sd] | 41.46 [12.51] | 44.4 [11.51] |
| Body mass index<br>mean [sd] | 23.5 [2.77] | 23.59 [2.72] |
| Pain duration (weeks)<br>mean [sd] | | 570.01 [551.03] |
| Pain Intensity (VAS)<br>mean [sd] | | 2.87 [1.86] |

Supplementary Table S8. Demographic data of cleaned questionnaire + clinic dataset

| | Asymptomatic | Chronic low back pain |
|---|---|---|
| Sample size | 522 | 413 |
| Females | 278 | 240 |
| Age (years) mean [sd] | 40.3 [12.42] | 43.42 [11.5] |
| Body mass index mean [sd] | 23.56 [2.73] | 23.53 [2.82] |
| Pain duration (weeks) mean [sd] | | 562.59 [543.63] |
| Pain Intensity (VAS) mean [sd] | | 2.86 [1.84] |

Supplementary Table S9. Demographic data of cleaned questionnaire + back shape and function dataset

| | Asymptomatic | Chronic low back pain |
|---|---|---|
| Sample size | 530 | 409 |
| Females | 283 | 246 |
| Age (years) mean [sd] | 40.45 [12.36] | 43.25 [11.54] |
| Body mass index mean [sd] | 23.54 [2.74] | 23.54 [2.82] |
| Pain duration (weeks) mean [sd] | | 563.61 [538.73] |
| Pain Intensity (VAS) mean [sd] | | 2.9 [1.86] |

Supplementary Table S10. Demographic data of cleaned questionnaire + MRI dataset

| | Asymptomatic | Chronic low back pain |
|---|---|---|
| Sample size | 383 | 278 |
| Females | 212 | 162 |
| Age (years) mean [sd] | 41.35 [12.22] | 44.19 [11.34] |
| Body mass index mean [sd] | 23.5 [2.77] | 23.6 [2.7] |
| Pain duration (weeks) mean [sd] | | 583.07 [569.01] |

| | | |
|---|---|---|
| Pain Intensity (VAS) mean [sd] | | 2.87 [1.84] |

Supplementary Table S11. Demographic data of cleaned clinic + back shape and function dataset

| | Asymptomatic | Chronic low back pain |
|---|---|---|
| Sample size | 573 | 463 |
| Females | 305 | 280 |
| Age (years) mean [sd] | 40.57 [12.59] | 43.74 [11.74] |
| Body mass index mean [sd] | 23.51 [2.74] | 23.59 [2.84] |
| Pain duration (weeks) mean [sd] | | 555.8 [526.21] |
| Pain Intensity (VAS) mean [sd] | | 2.89 [1.85] |

Supplementary Table S12. Demographic data of cleaned clinic + MRI dataset

| | Asymptomatic | Chronic low back pain |
|---|---|---|
| Sample size | 413 | 325 |
| Females | 228 | 194 |
| Age (years) mean [sd] | 41.26 [12.54] | 44.5 [11.48] |
| Body mass index mean [sd] | 23.44 [2.75] | 23.61 [2.72] |
| Pain duration (weeks) mean [sd] | | 577.98 [551.55] |
| Pain Intensity (VAS) mean [sd] | | 2.87 [1.84] |

Supplementary Table S13. Demographic data of cleaned back shape and function + MRI dataset

| | Asymptomatic | Chronic low back pain |
|---|---|---|
| Sample size | 448 | 334 |
| Females | 247 | 202 |
| Age (years) mean [sd] | 41.53 [12.54] | 44.46 [11.5] |
| Body mass index mean [sd] | 23.5 [2.78] | 23.59 [2.74] |
| Pain duration (weeks) mean [sd] | | 560.32 [537.8] |

| | | |
|---|---|---|
| Pain Intensity (VAS)<br>mean [sd] | | 2.87 [1.87] |

Supplementary Table S14. Demographic data of cleaned questionnaire + clinic + back shape and function dataset

| | Asymptomatic | Chronic low back pain |
|---|---|---|
| Sample size | 499 | 393 |
| Females | 264 | 234 |
| Age (years)<br>mean [sd] | 40.38 [12.4] | 43.56 [11.59] |
| Body mass index<br>mean [sd] | 23.54 [2.73] | 23.56 [2.84] |
| Pain duration (weeks)<br>mean [sd] | | 561.02 [536.44] |
| Pain Intensity (VAS)<br>mean [sd] | | 2.88 [1.85] |

Supplementary Table S15. Demographic data of cleaned questionnaire + clinic + MRI dataset

| | Asymptomatic | Chronic low back pain |
|---|---|---|
| Sample size | 356 | 267 |
| Females | 196 | 155 |
| Age (years)<br>mean [sd] | 41.53 [12.54] | 44.46 [11.5] |
| Body mass index<br>mean [sd] | 23.47 [2.76] | 23.6 [2.71] |
| Pain duration (weeks)<br>mean [sd] | | 581.41 [568.3] |
| Pain Intensity (VAS)<br>mean [sd] | | 2.86 [1.84] |

Supplementary Table S16. Demographic data of cleaned questionnaire + back shape and function + MRI dataset

| | Asymptomatic | Chronic low back pain |
|---|---|---|
| Sample size | 375 | 269 |
| Females | 205 | 158 |
| Age (years)<br>mean [sd] | 41.42 [12.25] | 44.27 [11.3] |

| | | |
|---|---|---|
| Body mass index<br>mean [sd] | 23.5 [2.78] | 23.62 [2.73] |
| Pain duration (weeks)<br>mean [sd] | | 571.48 [533.96] |
| Pain Intensity (VAS)<br>mean [sd] | | 2.87 [1.86] |

Supplementary Table S17. Demographic data of cleaned clinic + back shape and function + MRI dataset

| | **Asymptomatic** | **Chronic low back pain** |
|---|---|---|
| Sample size | 406 | 317 |
| Females | 222 | 190 |
| Age (years)<br>mean [sd] | 41.32 [12.56] | 44.51 [11.52] |
| Body mass index<br>mean [sd] | 23.45 [2.76] | 23.62 [2.74] |
| Pain duration (weeks)<br>mean [sd] | | 566.27 [537.69] |
| Pain Intensity (VAS)<br>mean [sd] | | 2.87 [1.85] |

Supplementary Table S18. Demographic data of cleaned questoinmaire + clinic + back shape and function + MRI dataset

| | **Asymptomatic** | **Chronic low back pain** |
|---|---|---|
| Sample size | 349 | 259 |
| Females | 190 | 151 |
| Age (years)<br>mean [sd] | 41.31 [12.36] | 44.38 [11.32] |
| Body mass index<br>mean [sd] | 23.48 [2.77] | 23.61 [2.74] |
| Pain duration (weeks)<br>mean [sd] | | 567.21 [552.41] |
| Pain Intensity (VAS)<br>mean [sd] | | 2.85 [1.85] |

Supplementary Table S19. List of all variables in the Berlin Back Study.

| **Modality** | **Variable** | **Explanation** | **Units** | **Missing Values** |
|---|---|---|---|---|
| Demographic | age | age of the participant at testing | years | 17 |
| Demographic | sex | sex of the participant at testing | Male, female | 17 |

| | | | | |
|---|---|---|---|---|
| Demographic | clinic chronic LBP [Target] | classification of participant as having LBP, not having LBP, or having LBP in the past as diagnosed by a clinician | Yes, No, in past | 23 |
| Demographic | question chronic LBP | Participant answer of question do they suffer from chronic LBP | Yes, No | 45 |
| Demographic | LBP past pain duration | In past pain duration | days | 1192 |
| Demographic | LBP pain duration | pain duration | weeks | 643 |
| Demographic | height | height of participant as testing | cm | 51 |
| Demographic | weight | weigh tof participant at testing | Kg | 52 |
| Demographic | BMI | Body mass index | Kg/m2 | 52 |
| Demographic | waist | diameter of waist | cm | 108 |
| Demographic | hip | diameter of hip | cm | 107 |
| Demographic | waist - hip ratio | ratio between hip and waist | | 108 |
| Demographic | leg - left | length of left leg | cm | 32 |
| Demographic | leg - right | length of right leg | cm | 32 |
| Demographic | right- left leg ratio | ratio between leg length | | 32 |
| Demographic | pain primary location | Loaction of primary pain site | | 40 |
| Demographic | pain secondary location | Loaction of secondary pain site | | 224 |
| Demographic | current pain | Visual Analoug Scale (VAS) 0-10 | | 68 |
| Questionnaire | pain begin | Participant answer of question how would they rate their pain at the moment? | 1) sudden onset after exertion, 2) slowly increasing, 3) after an incorrect movement, 4) Progressive after previous low back pain, 5) after an accident | 51 |
| Questionnaire | pain coughing sneezing | Participant answer of question is their pain intensified when coughing or sneezing? | Yes, No | 59 |
| Questionnaire | pain standing | Participant answer of question is their pain intensified when standing? | Yes, No | 59 |
| Questionnaire | pain walking | Participant answer of question is their pain intensified when walking? | Yes, No | 61 |
| Questionnaire | pain sitting | Participant answer of question is their pain intensified when sitting? | Yes, No | 62 |
| Questionnaire | pain resting | Participant answer of question is their pain independent of exertion? | Yes, No | 64 |

| | | | | |
|---|---|---|---|---|
| Questionnaire | pain day diff | Participant answer of question do they experience differences in pain intensity depending on the time of day? | Yes, No | 67 |
| Questionnaire | pain when strongest | Participant answer of question when is their pain most severe? | 1) only during the day<br>2) only at night<br>3) always equally strong<br>4) Days > Nights<br>5) Nights > Days | 79 |
| Questionnaire | pain arm | Accompanying symptoms: pain arm | Yes, No | 536 |
| Questionnaire | pain leg | Accompanying symptoms: pain leg | Yes, No | 528 |
| Questionnaire | prior general disease | Symptoms such as fever or a general feeling of illness? | Yes, No | 528 |
| Questionnaire | pain walking distance | Participant answer of question which distance they can walk unrestricted. | unrestricted<br>1000 - 2000 m<br>500 - 1000 m<br>100 - 500 m<br>< 100 m | 388 |
| Questionnaire | pain movment limit | Participant answer of question is their movement restricted. | Yes, No | 491 |
| Questionnaire | pain sensitivity | Participant answer of question whether they have sensory disturbances/feeling disorders? | Yes, No | 496 |
| Questionnaire | pain paralysis | Participant answer of question whether they have sensory disturbances/feeling disorders? | Yes, No | 513 |
| Questionnaire | pain paralysis location | Participant answer of question for pain paralysis location | C1, C2, C3, C4, C5, C6, C7, C8,<br>T1, T2, T3, T4,<br>T5, T6, T7, T8, T9,<br>T10, T11, T12, L1, L2,<br>L3, L4, L5, S1, S2, S3,<br>S4, S5, RH Anesthesia,<br>Multisegmental, Arm,<br>Multisegmental leg | 1282 |
| Questionnaire | sensory symptom | Participant answer of question whether they have sensory symptoms and if yes, where? | Bladder and rectal disorders<br>Genital area<br>Thigh<br>Lower leg<br>Outer edge of foot<br>sole of the foot<br>Instep<br>Toes<br>Hand | 826 |

| | | | | |
|---|---|---|---|---|
| Questionnaire | sensory deficit | Participant answer of question whether they have sensory deficits. | Yes, No | 540 |
| Questionnaire | sensory location | …if yes, where? | C1, C2, C3, C4, C5, C6, C7, C8, T1, T2, T3, T4, T5, T6, T7, T8, T9, T10, T11, T12, L1, L2, L3, L4, L5, S1, S2, S3, S4, S5, RH Anesthesia, Multisegmental, Arm, Multisegmental leg | 1216 |
| Questionnaire | prior diagnosis | Participant answer of question whether they have prior diagnosis. | Yes, No | 40 |
| Questionnaire | prior diagnosis which | …if yes, which? | Intervertebral discs – protrusion, Intervertebral discs – prolapse, Intervertebral discs – sequestration, Spondylolisthesis, Spondylarthrosis, Osteochrondrosis, Spinal canal stenosis, Vertebral body fracture, Osteoporosis, Hip-left, Hip-right, Hip on both sides, Scoliosis, Multiple diagnosis spine | 854 |
| Questionnaire | prior diagnosis back level | …when prior back diagnosis which level? | L1, L2, L3, L4, L5, L1-L2, L2-L3, L3-L4, L4-L5, L5-S1, T1-T2, T2-T3, T3-T4, T4-T5, T5-T6, T6-T7, T7-T8, T8-T9, T9-T10, T10-T11, T11-T12, Cervical | 1081 |
| Questionnaire | prior surgeries | Participant answer of question whether they have prior surgeries. | Yes, No | 96 |

| | | | | |
|---|---|---|---|---|
| Questionnaire | prior surgeries which | …if yes, which? | Spondylodesis, decompression surgery, disc surgery, vertebroplasty, kyphoplasty, vertebral body stenting, hip-left, hip-right, hip-bilateral, knee-left, knee-right, knee-bilateral, ankle-left, ankle-right, ankle-bilateral, foot-left, foot-right, foot-bilateral, Multiple pre-ops | 1161 |
| Questionnaire | prior surgeries spine level | …when prior spine surgeries which level? | L1, L2, L3, L4, L5, L1-L2, L2-L3, L3-L4, L4-L5, L5-S1, T1-T2, T2-T3, T3-T4, T4-T5, T5-T6, T6-T7, T7-T8, T8-T9, T9-T10, T10-T11, T11-T12 | 1293 |
| Questionnaire | physically active for at least 150 min / week | Participant answer of question whether they are physically active for at least 150 min / week | 1) No, and I don't plan to<br>2) No, but I am thinking about becoming more active<br>3) No, but I am determined to do so<br>4) Yes, but I find it difficult<br>5) Yes, and I find it very easy | 38 |
| Questionnaire | covid19 activity | Participant answer of question whether covid 19 affected their everyday functionality | substantial, moderate, hardly, not | 24 |
| Questionnaire | alcohol frequency | Participant answer of question how often do they drink alcohol. | never, less than once a month, two to four times a month, two to three times a week, four or more times a week | 23 |
| Questionnaire | alcohol number glasses | … if yes, how many glasses do they drink in a day? | 1-2, 3-4, 5-6, 7-9, 10 or more | 26 |
| Questionnaire | alcohol restriction | Participants' response to the question of how often their alcohol consumption has prevented them from doing what was expected of them in the last 12 months. | never, less than once a month, once a month, once a week, daily or almost daily | 28 |
| Questionnaire | smoking | Participants' response to the question do they currently smoke? | Yes, No | 24 |

| | | | | |
|---|---|---|---|---|
| Questionnaire | smoking pack years | …if yes, how many pack years | Cigarette packs per year | 417 |
| Questionnaire | medication 1 registration | Participants' response to the question what medication they take regularly | No medication, NSAIDs: Acetylsalicylic acid, NSAIDs: Ibuprofen, NSAIDs: Diclofenac, NSAIDs: Indomethacin, NSAIDs: Aspirin, NSAIDs: Coxibe, NSAIDs: Novaminsulfon, Analgesics: Paracetamol, Corticosteroids: hydrocortisone, Corticosteroids: prednisolone, Local anesthetics, Local anesthetics plus corticosteroids, Muscle relaxants: Tizanidine, muscle relaxants: Methocarbamol, Flupirtine, Opioids: morphine, Opioids: fentanyl, Opioids: fentanyl patches, Opioids: tramadol, Opioids: tilidine, Opioids: capsaicin patches and creams, Opioids: oxycondon, Opioids: tapentadol, Antidepressants: amitriptyline, Antidepressants: doxepin, Antidepressants: clomipramine, Antidepressants: escitalopram, Antidepressants: citalopram, Antidepressants: sertraline, Antidepressants: mirtazapine, Antidepressants: not named, Neur analgesics: Gabapentin, Neur analgesics: Pregabalin, Benzodiazepine, | 130 |

| | | | | |
|---|---|---|---|---|
| | | | sleeping pills, Combination preparations: keltican, intigrin antagonist: α4β7 integrin antagonist, herbal remedy: devil's claw, herbal remedy: green-lipped mussel | |
| Questionnaire | medication 1 dosis mg | …if yes what dose? | | 1255 |
| Questionnaire | medication 1 morning | …if yes what dose? | | 1259 |
| Questionnaire | medication 1 midday | …if yes what dose? | | 1271 |
| Questionnaire | medication 1 noon | …if yes what dose? | | 1265 |

| | | | | |
|---|---|---|---|---|
| Questionnaire | medication 2 registration | Participants' response to the question what second medication they take regulary | No medication, NSAIDs: Acetylsalicylic acid, NSAIDs: Ibuprofen, NSAIDs: Diclofenac, NSAIDs: Indomethacin, NSAIDs: Aspirin, NSAIDs: Coxibe, NSAIDs: Novaminsulfon, Analgesics: Paracetamol, Corticosteroids: hydrocortisone, Corticosteroids: prednisolone, Local anesthetics, Local anesthetics plus corticosteroids, Muscle relaxants: Tizanidine, muscle relaxants: Methocarbamol, Flupirtine, Opioids: morphine, Opioids: fentanyl, Opioids: fentanyl patches, Opioids: tramadol, Opioids: tilidine, Opioids: capsaicin patches and creams, Opioids: oxycondon, Opioids: tapentadol, Antidepressants: amitriptyline, Antidepressants: doxepin, Antidepressants: clomipramine, Antidepressants: escitalopram, Antidepressants: citalopram, Antidepressants: sertraline, Antidepressants: mirtazapine, Antidepressants: not named, Neur analgesics: Gabapentin, Neur analgesics: Pregabalin, Benzodiazepine, sleeping pills, Combination | 310 |

| | | | | |
|---|---|---|---|---|
| | | | preparations: keltican, intigrin antagonist: α4β7 integrin antagonist, herbal remedy: devil's claw, herbal remedy: green-lipped mussel | |
| Questionnaire | medication 2 dosis mg | …if yes what dose? | | 1287 |
| Questionnaire | medication 2 morning | …if yes what dose? | | 1288 |
| Questionnaire | medication 2 midday | …if yes what dose? | | 1290 |
| Questionnaire | medication 2 noon | …if yes what dose? | | 1289 |

| | | | | |
|---|---|---|---|---|
| Questionnaire | medication 1 demand | Participants' response to the question what medication they take on request | No medication, NSAIDs: Acetylsalicylic acid, NSAIDs: Ibuprofen, NSAIDs: Diclofenac, NSAIDs: Indomethacin, NSAIDs: Aspirin, NSAIDs: Coxibe, NSAIDs: Novaminsulfon, Analgesics: Paracetamol, Corticosteroids: hydrocortisone, Corticosteroids: prednisolone, Local anesthetics, Local anesthetics plus corticosteroids, Muscle relaxants: Tizanidine, muscle relaxants: Methocarbamol, Flupirtine, Opioids: morphine, Opioids: fentanyl, Opioids: fentanyl patches, Opioids: tramadol, Opioids: tilidine, Opioids: capsaicin patches and creams, Opioids: oxycondon, Opioids: tapentadol, Antidepressants: amitriptyline, Antidepressants: doxepin, Antidepressants: clomipramine, Antidepressants: escitalopram, Antidepressants: citalopram, Antidepressants: sertraline, Antidepressants: mirtazapine, Antidepressants: not named, Neur analgesics: Gabapentin, Neur analgesics: Pregabalin, Benzodiazepine, sleeping pills, Combination | 130 |

| | | | | |
|---|---|---|---|---|
| | | | preparations: keltican, intigrin antagonist: α4β7 integrin antagonist, herbal remedy: devil's claw, herbal remedy: green-lipped mussel | |
| Questionnaire | medication 1 demand dosis mg | …if yes what dose? | | 624 |
| Questionnaire | medication 1 demand month | …if yes what dose? | | 814 |

| | | | | |
|---|---|---|---|---|
| Questionnaire | medication 2 demand | Participants' response to the question what medication they take on request | No medication, NSAIDs: Acetylsalicylic acid, NSAIDs: Ibuprofen, NSAIDs: Diclofenac, NSAIDs: Indomethacin, NSAIDs: Aspirin, NSAIDs: Coxibe, NSAIDs: Novaminsulfon, Analgesics: Paracetamol, Corticosteroids: hydrocortisone, Corticosteroids: prednisolone, Local anesthetics, Local anesthetics plus corticosteroids, Muscle relaxants: Tizanidine, muscle relaxants: Methocarbamol, Flupirtine, Opioids: morphine, Opioids: fentanyl, Opioids: fentanyl patches, Opioids: tramadol, Opioids: tilidine, Opioids: capsaicin patches and creams, Opioids: oxycondon, Opioids: tapentadol, Antidepressants: amitriptyline, Antidepressants: doxepin, Antidepressants: clomipramine, Antidepressants: escitalopram, Antidepressants: citalopram, Antidepressants: sertraline, Antidepressants: mirtazapine, Antidepressants: not named, Neur analgesics: Gabapentin, Neur analgesics: Pregabalin, Benzodiazepine, sleeping pills, Combination | 413 |

| | | | | |
|---|---|---|---|---|
| | | | preparations: keltican, intigrin antagonist: α4β7 integrin antagonist, herbal remedy: devil's claw, herbal remedy: green-lipped mussel | |
| Questionnaire | medication 2 demand dosis mg | …if a second medication is taken what dose? | | 1195 |
| Questionnaire | medication 2 demand month | …if a second medication is taken which duration? | months | 1222 |
| Questionnaire | family back pain | Participants' response to the question is there back pain in their family? | Yes, No | 51 |

| | | | | |
|---|---|---|---|---|
| Questionnaire | job field | Participants' response to the question in which professional field do they work? | Construction, architecture, surveying<br>Services<br>Electrical<br>Healthcare<br>IT, computers<br>art, culture, design<br>Agriculture, nature, environment<br>media,<br>metal, mechanical engineering<br>Natural sciences<br>Production, manufacturing<br>Social affairs, education<br>Technology, technology fields<br>Transportation, logistics<br>Economy, administration<br>Unemployed<br>Disabled/retired<br>Parental leave<br>Student<br>Pupil | 22 |
| Questionnaire | job posture | Participants' response to the question what is the primary posture that characterizes the professional field. | sitting, standing, sitting + standing, standing + walking, unemployed | 25 |
| Questionnaire | job posture duration | Participants' response to the question how the duration of the primary posture is. | Hours per day | 321 |
| Questionnaire | job load 1 | Participants' response to the question of how they would characterize the primary physical workload in this area. | low load, moderate load, heavy load, vibrations, vibrations + heavy load, unemployed | 34 |
| Questionnaire | job load 1 duration | Participants' response to the question of how long the workload lasts. | Hours per day | 62 |
| Questionnaire | job load 2 | Participants' response to the question of how they would characterize the secondary physical workload in this area. | low load, moderate load, heavy load, vibrations, vibrations + heavy load, unemployed | 1019 |
| Questionnaire | job load 2 duration | Participants' response to the question of how long the workload lasts. | Hours per day | 1027 |

| | | | | |
|---|---|---|---|---|
| Questionnaire | job duration years | Participants' response to the question of how long they have been working in this area. | Years | 59 |
| Questionnaire | pyschological stress | Participants' response to the question of whether there are signs of psychosocial stress or strain due to their family situation. | Yes, No | 23 |
| Questionnaire | living situation | Participants' response to the question of whether they live alone or in a larger community. | alone, family, shared flat, home | 23 |
| Questionnaire | family conflicts | Participants' response to the question of whether family conflicts currently exist. | Yes, No | 23 |
| Questionnaire | self assessment | Participants' response to the question about their back functionality on a scale from 1 (very good) to 10 (severely limited). | 1 to 10 | 23 |
| Clinical | cervical inclination | The participants are asked to bend their cervical spine forward. | Degree | 22 |
| Clinical | cervical reclination | The participants are asked to bend their cervical spine backward. | Degree | 22 |
| Clinical | chin sternum distance | | mm | 25 |
| Clinical | cervical lateral bend - left | The participants are asked to bend their cervical spine to the left. | Degree | 23 |
| Clinical | cervical lateral bend - right | The participants are asked to bend their cervical spine to the right. | Degree | 23 |
| Clinical | cervical axial rotate - left | The participants are asked to rotate their cervical spine to the left. | Degree | 23 |
| Clinical | cervical axial rotate - right | The participants are asked to rotate their cervical spine to the left. | Degree | 24 |
| Clinical | thoracic lumbar inclination | The participants are asked to perform maximum flexion. | Degree | 22 |
| Clinical | thoracic lumbar reclination | The participants are asked to perform maximum extension. | Degree | 26 |
| Clinical | finger floor distance | The participants are asked to perform maximum flexion. | mm | 22 |
| Clinical | hip flexion | The participants are asked to perform maximum flexion. | Degree | 300 |

| | | | | |
|---|---|---|---|---|
| Clinical | thoracic lumbar lateral bending - left | The participants are asked to perform maximum lateral bending to the left. | Degree | 24 |
| Clinical | thoracic lumbar lateral bending - right | The participants are asked to perform maximum lateral bending to the right. | Degree | 23 |
| Clinical | thoracic lumbar axial rotation - left | The participants are asked to perform maximum axial rotation to the left. | Degree | 28 |
| Clinical | thoracic lumbar axial rotation - right | The participants are asked to perform maximum axial rotation to the right. | Degree | 28 |
| Clinical | motion OTT | The participants are asked to perform maximum flexion. | mm | 24 |
| Clinical | shober | The participants are asked to perform maximum flexion. | mm | 23 |
| Clinical | square shoulders | Participants are asked to stand upright. | Straight line, Right > Left, Left > Right | 23 |
| Clinical | plump line | Non-specific parameter with possible indication of scoliosis (spinal curvature), postural asymmetry, etc. Participants are asked to stand upright. | Yes, No | 22 |
| Clinical | rib hump | The ribs on one side of the spine are pushed backwards. Participants are asked to stand upright. | No, Right, Left | 22 |
| Clinical | lumbar bulge | When the upper body is bent forward, the back should look the same on the right and left side. The participants are asked to perform maximum flexion. | No, Right, Left | 22 |
| Clinical | asym waist triangle | Non-specific parameter with possible indication of scoliosis (spinal curvature), postural asymmetry, etc. Participants are asked to stand upright. | Yes, No | 22 |

| | | | | |
|---|---|---|---|---|
| Clinical | back form | Normal findings: harmonious lumbar and thoracic spine curvature, flat back: reduced lumbar and thoracic spine curvature, gibbus: short-arched vertebral hump, hyperlordosis: increased lumbar curvature, hyperkyphosis: increased curvature of the thoracic spine, scoliosis: lateral deviation of the spine from the longitudinal axis with rotation (twisting) of the vertebrae around the longitudinal axis and torsion of the vertebral bodies. Participants are asked to stand upright. | Normal findings, Flat back, Gibbus, Hyperlordosis, Hyperkyphosis, Scoliosis, | 22 |
| Clinical | roussoly type | Lateral assessment of the spinal profile according to Roussouly. Participants are asked to stand upright. | 1, 2, 3, 4 | 24 |
| Clinical | sagittal balance | Lateral assessment of the spinal profile to determine whether the perpendicular of the center of the 7th cervical vertebra is approximately in line with the upper edge of the sacrum (part of the posterior pelvic ring). Participants are asked to stand upright. | Yes, No | 22 |
| Clinical | coronal balance | Assessment of the spinal profile from the front to determine whether the perpendicular of the center of the 7th cervical vertebra represents the center of the body (the head and cervical spine are in a central position). Participants are asked to stand upright. | Yes, No | 25 |
| Clinical | back head wall distance | Remaining distance between the back of the head and the wall when the heels/buttocks/shoulders are in contact with the wall as an indication of restricted spinal mobility, standard value: 0 cm. Participants are asked to stand upright at a wall. | cm | 39 |

| | | | | |
|---|---|---|---|---|
| Clinical | Muscular hard tension | Hardened/tense actual muscle parts | No,<br>Cervical,<br>Thoracic,<br>Lumbar,<br>Several back sections | 26 |
| Clinical | pain pressure | Pressure pain over the spinous processes of the vertebral bodies as an indication of functional or structural changes or tension. | No,<br>Cervical,<br>Thoracic,<br>Lumbar,<br>Several back sections | 22 |
| Clinical | pain knocking | Tapping pain over the spinous processes of the vertebral bodies as an indication of functional or structural changes or tensio | No,<br>Cervical,<br>Thoracic,<br>Lumbar,<br>Several back sections | 22 |
| Clinical | pain spinous process shake | Pain in the spinous processes of the vertebral bodies as an indication of functional or structural changes or tension. | No,<br>Cervical,<br>Thoracic,<br>Lumbar,<br>Several back sections | 22 |
| Clinical | pain mennell sign | Indication of functional or structural change of the ilio-sacral joint | Negative<br>Right Positive<br>Left positive<br>Positive on both sides | 25 |
| Clinical | pain jumping | Test to examine the mobility of the vertebrae in relation to each other | Yes, No | 38 |
| Clinical | General mobility | Rating of general mobility into three categories | Normal mobility,<br>Hypomobile,<br>Hypermobile, | 37 |
| Clinical | toe stand | Orienting test to examine for reduced strength or signs of paralysis of the calf muscles | Yes, No | 22 |
| Clinical | heel stand | Orientation test to examine for reduced strength or signs of paralysis of the foot lifter muscles | Yes, No | 22 |
| Clinical | pain heel drop | Pain provocation due to compression of the axial skeleton as an indication of functional or structural changes or inflammation | Yes, No | 23 |
| Clinical | trendelenburg sign | Sinking of the opposite hip when standing on one leg as a sign of insufficiency of the gluteal muscles (gluteus medius/minimus) | Negative<br>Right Positive<br>Left positive<br>Positive on both sides | 23 |

| | | | | |
|---|---|---|---|---|
| Clinical | straight leg raise | Pain provocation in the back leg in the range of 20-60° hip flexion with the leg extended due to nerve stretching/stretching of the nerve roots as an indication of nerve (root) compression | Negative<br>Right Positive<br>Left positive<br>Positive on both sides | 24 |
| Clinical | pseudo lasegue - left | Pain provocation in the back leg in the range of 20-60° hip flexion with the leg extended due to nerve stretching/stretching of the nerve roots as an indication of nerve (root) compression | Degree | 310 |
| Clinical | pseudo lasegue - right | Pain provocation in the back leg in the range of 20-60° hip flexion with the leg extended due to nerve stretching/stretching of the nerve roots as an indication of nerve (root) compression | Degree | 311 |
| Clinical | gait pattern | Fluent gait: normal findings, Limping gait: non-specific gait asymmetry, Gait ataxia: unsteady gait, Stepper gait: altered gait due to foot drop palsy, Wernicke-Mann gait: altered gait after a central disorder (e.g. stroke), Duchenne limp: gait alteration due to muscular insufficiency of the small gluteal muscles | luid gait<br>Limping gait<br>gait ataxia<br>stepping gait<br>Wernicke-Mann gait pattern<br>Duchenne limp | 24 |
| Clinical | reflex quadricep - left | Quadriceps tendon reflex: Tensing of the anterior thigh muscle after striking the patellar tendon with a reflex hammer | Areflexia<br>Hyporeflexia<br>Normoreflexia<br>hyperreflexia | 23 |
| Clinical | reflex quadricep - right | Quadriceps tendon reflex: Tensing of the anterior thigh muscle after striking the patellar tendon with a reflex hammer | Areflexia<br>Hyporeflexia<br>Normoreflexia<br>hyperreflexia | 24 |
| Clinical | reflex achilles - left | Achilles tendon reflex: Tensing of the calf muscles after striking the Achilles tendon with a reflex hammer | Areflexia<br>Hyporeflexia<br>Normoreflexia<br>hyperreflexia | 24 |
| Clinical | reflex achilles - right | Achilles tendon reflex: Tensing of the calf muscles after striking the Achilles tendon with a reflex hammer | Areflexia<br>Hyporeflexia<br>Normoreflexia<br>hyperreflexia | 27 |
| Questionnaire | pain hip | Pain at the hip | No<br>Left<br>Right<br>Both sides | 41 |

| | | | | |
|---|---|---|---|---|
| Clinical | hip flexion left | Flexion: hip flexion - left | Degree | 36 |
| Clinical | hip flexion - right | Flexion: hip flexion - right | Degree | 36 |
| Clinical | hip extension - left | Extension: hip extension - left | Degree | 39 |
| Clinical | hip extension - right | Extension: hip extension - right | Degree | 41 |
| Clinical | hip external rotate - left | External rotation: outward rotation of the hip joint - left | Degree | 37 |
| Clinical | hip external rotate - right | External rotation: outward rotation of the hip joint - right | Degree | 36 |
| Clinical | hip internal rotate - left | Internal rotation: inward rotation of the hip joint - left | Degree | 37 |
| Clinical | hip internal rotate - right | Internal rotation: inward rotation of the hip joint - right | Degree | 36 |
| Clinical | hip abduction - left | movement of the leg away from the midline of the body | Degree | 36 |
| Clinical | hip abduction - right | movement of the leg away from the midline of the body | Degree | 36 |
| Clinical | hip adduction - left | Abduction: spreading apart in the hip joint | Degree | 37 |
| Clinical | hip adduction - right | Adduction: spreading in the hip joint | Degree | 37 |
| Clinical | thomas handle - left | Thomas grip: indicative test for shortening of the hip flexor muscles | Positive, Negative | 37 |
| Clinical | thomas handle - right | Thomas grip: indicative test for shortening of the hip flexor muscles | Positive, Negative | 39 |
| Clinical | pelvic tilt frontal | Frontal pelvic position: Indication of leg length difference/postural asymmetry | Straight position<br>Left > Right<br>Right > Left | 42 |
| Clinical | pelvic tilt sagittal | Sagittal pelvic position: Different standard variants of the "pelvic tilt" viewed from the side | Straight position<br>Pool forward tilt<br>Pool back tilt | 39 |
| Clinical | sit to stand 30sec | The participant is instructed to complete as many full stands as possible within the 30 seconds | Number of complete stands | 55 |
| Clinical | heart rate | | / min | 67 |
| Clinical | breath rate | | / min | 172 |
| Clinical | heart systolic pressure | | mmHg | 46 |
| Clinical | heart diastolic pressure | | mmHg | 48 |
| Questionnaire | sf36 physical function | Heatlth questionnaire. Transformed into a 0-100 scale. The lower the score the more disability. The higher the score the less disability | 0-100 | 41 |

| | | | | |
|---|---|---|---|---|
| Questionnaire | sf36 physical role function | Heatlth questionnaire. Transformed into a 0-100 scale. The lower the score the more disability. The higher the score the less disability | 0-100 | 39 |
| Questionnaire | sf36 physical pain | Heatlth questionnaire. Transformed into a 0-100 scale. The lower the score the more disability. The higher the score the less disability | 0-100 | 36 |
| Questionnaire | sf36 health perception | Heatlth questionnaire. Transformed into a 0-100 scale. The lower the score the more disability. The higher the score the less disability | 0-100 | 34 |
| Questionnaire | sf36 vitality | Heatlth questionnaire. Transformed into a 0-100 scale. The lower the score the more disability. The higher the score the less disability | 0-100 | 36 |
| Questionnaire | sf36 social function | Heatlth questionnaire. Transformed into a 0-100 scale. The lower the score the more disability. The higher the score the less disability | 0-100 | 42 |
| Questionnaire | sf36 emotional role function | Heatlth questionnaire. Transformed into a 0-100 scale. The lower the score the more disability. The higher the score the less disability | 0-100 | 36 |
| Questionnaire | sf36 psychological well being | Heatlth questionnaire. Transformed into a 0-100 scale. The lower the score the more disability. The higher the score the less disability | 0-100 | 36 |
| Questionnaire | therapy 1 | Have you received pain treatment so far? | Yes, No | 77 |
| Questionnaire | therapy 2 | Medications | Yes, No | 105 |
| Questionnaire | therapy 2 - 1 | Effectiveness | Yes, partially, no | 483 |
| Questionnaire | therapy 3 | Infusion | Yes, No | 119 |
| Questionnaire | therapy 3 - 1 | Efectiveness | Yes, partially, no | 852 |
| Questionnaire | therapy 4 | Injections into the area of pain, nerve blocks | Yes, No | 109 |
| Questionnaire | therapy 4 - 1 | Efectiveness | Yes, partially, no | 660 |
| Questionnaire | therapy 5 | Injections in the spinal cord (e.g. epidural) | Yes, No | 119 |
| Questionnaire | therapy 5 - 1 | Efectiveness | Yes, partially, no | 852 |
| Questionnaire | therapy 6 | Spinal cord probe (SCS) or pump systems | Yes, No | 121 |

| | | | | |
|---|---|---|---|---|
| Questionnaire | therapy 6 - 1 | Efectiveness | Yes, partially, no | 891 |
| Questionnaire | therapy 7 | Physical therapy | Yes, No | 93 |
| Questionnaire | therapy 7 - 1 | Efectiveness | Yes, partially, no | 348 |
| Questionnaire | therapy 8 | Massages, baths, cold/heat therapy | Yes, No | 101 |
| Questionnaire | therapy 8 - 1 | Efectiveness | Yes, partially, no | 403 |
| Questionnaire | therapy 9 | Electrical nerve stimulation (TENS) | Yes, No | 113 |
| Questionnaire | therapy 9 - 1 | Efectiveness | Yes, partially, no | 729 |
| Questionnaire | therapy 10 | Acupuncture | Yes, No | 108 |
| Questionnaire | therapy 10 - 1 | Efectiveness | Yes, partially, no | 704 |
| Questionnaire | therapy 11 | Chiropratic | Yes, No | 109 |
| Questionnaire | therapy 11 - 1 | Efectiveness | Yes, partially, no | 685 |
| Questionnaire | therapy 12 | Psychotherapy | Yes, No | 115 |
| Questionnaire | therapy 12 - 1 | Efectiveness | Yes, partially, no | 784 |
| Questionnaire | therapy 13 | Relaxation techniques, hypnosis, biofeedback | Yes, No | 116 |
| Questionnaire | therapy 13 - 1 | Efectiveness | Yes, partially, no | 809 |
| Questionnaire | therapy 14 | Medication withdrawal | Yes, No | 119 |
| Questionnaire | therapy 14 - 1 | Efectiveness | Yes, partially, no | 896 |
| Questionnaire | therapy 15 | Spa/rehab treatment | Yes, No | 113 |
| Questionnaire | therapy 15 - 1 | Efectiveness | Yes, partially, no | 782 |
| Questionnaire | employment currently | Are you currently working? (also applies if you are currently unable to work) | Yes, No | 62 |
| Questionnaire | employment unable work | Are you currently unable to work? | Yes, No | 61 |
| Questionnaire | employment unable work could return | If unable to work, do you think you will be able to return to your old job? Able to? | Yes, No | 766 |
| Questionnaire | employment unable work last 3 months | How many days have you been unable to work in the last 3 months? | 0-92 | 151 |
| Questionnaire | employment pension application | Do you intend to submit a pension application or an application to change your pension? | Yes, No | 83 |
| Questionnaire | employment pension application not decided | Have you submitted a pension application / application for a pension change that has not yet been decided? | Yes, No | 86 |
| Questionnaire | employment pension application rejected | Has a pension application already been rejected? | Yes, No | 105 |
| Questionnaire | employment pension application opposition | Is a pension application currently in the objection procedure? | Yes, No | 104 |

| | | | | |
|---|---|---|---|---|
| Questionnaire | employment pension | Do you currently recieve a pension? | Yes, No | 83 |
| Questionnaire | employment pension for time | If Yes, for a fixed time? | Yes, No | 780 |
| Questionnaire | employment pension final | If Yes, for good? | Yes, No | 786 |
| Questionnaire | employment pension type | If so, what type of retirement? | Early retirement pension<br>Partial reduction in earning capacity<br>Full reduction in earning capacity<br>Occupational disability<br>Disability<br>Accident pension<br>Reaching the age limit | 898 |
| Questionnaire | employment disability | Do you have a recognized degree of disability (e.g. by the Office for Pension Matters)? | Yes, No | 119 |
| Questionnaire | employment disability grade | If so, how high is the disability grade | 1-… | 859 |
| Questionnaire | chron 1 | Which of the statements best applies to your pain in the last 12 weeks? | Constant pain with slight fluctuations<br>Continuous pain with strong fluctuations<br>Pain attacks in between pain-free<br>Pain attacks also pain in between<br>Had no pain in the last 12 weeks | 236 |
| Questionnaire | chron 2 | If you suffer from pain attacks, please answer the following questions: 1) How often do these attacks occur on average? | several times a day<br>once a day<br>several times a week<br>once a week<br>several times a month<br>once a month | 593 |
| Questionnaire | chron 3 | 2) How long do these attacks last on average? | Seconds<br>minutes<br>hours<br>up to 3 days<br>longer than 3 days | 589 |
| Questionnaire | ipaq met vigorous | MET minutes of vigorous activity in a day | MET minutes | 218 |
| Questionnaire | ipaq met moderate | MET minutes of moderate activity in a day | MET minutes | 197 |
| Questionnaire | ipaq met walking | MET minutes of walking activity in a day | MET minutes | 136 |
| Questionnaire | ipaq met sitting | MET minutes of sitting activity in a day | MET minutes | 67 |

| | | | | |
|---|---|---|---|---|
| Questionnaire | ipaq met sum | sum total of MET minutes in a day | MET minutes | 37 |
| Questionnaire | kinesiophobia | Total kinesiophobia value by summing the answers to 11 Tampa Scale for Kinesiophobia questions with values of 1-4 for do not agree at all, more or less disagree, more or less agree, completely agree | 11 - 44 summation score | 332 |
| Questionnaire | srbai sum | Behavioural Automaticity summation score of 4 questionaire (1-6) | 4 - 24 summation score | 0 |
| Questionnaire | brsq intrinsic motivation | summation score of BRSQ questions 1 & 3:<br>Q1 - I am happy and satisfied when I am physically active.<br>Q3 - I am physically active because I enjoy it. | 1 - Does not apply at all<br>2 - Does not apply<br>3 - Does not really apply<br>4 - Rather true<br>5 - applies<br>6 - fully applies | 33 |
| Questionnaire | brsq integrated regulation | summation score of BRSQ questions 2 & 4:<br>Q2 - I consider physical activity to be an important part of me.<br>Q4 - I see physical activity as part of my identity. | 1 - Does not apply at all<br>2 - Does not apply<br>3 - Does not really apply<br>4 - Rather true<br>5 - applies<br>6 - fully applies | 34 |
| Questionnaire | brsq external regulation | summation score of BRSQ questions 5 & 6:<br>Q5 - I consider physical activity to be an important part of me.<br>Q6 - I see physical activity as part of my identity. | 1 - Does not apply at all<br>2 - Does not apply<br>3 - Does not really apply<br>4 - Rather true<br>5 - applies<br>6 - fully applies | 34 |
| Questionnaire | stress days | Summation of stress day questions 1 - 4:<br>Q1 - Little interest or pleasure in your activities<br>Q2 - Depression, melancholy or hopelessness<br>Q3 - Nervousness, anxiety or tension<br>Q4 - Not being able to stop or control worries | 1 - not at all<br>2 - on individual days<br>3 - on more than half of the days<br>4 - almost every day | 330 |

| | | | | | |
|---|---|---|---|---|---|
| Questionnaire | stress score | Summation of stress day questions 5 - 8:<br>Q5 -How often in the last month have you had the feeling that you couldn't influence important things in your life?<br>Q6 - How often in the last month did you feel confident in dealing with personal tasks and problems?<br>Q7 - How often in the last month have you had the feeling that things are going your way?<br>Q8 - How often in the last month have you had the feeling that your problems have piled up to such an extent that you can no longer cope with them? | 1 - never<br>2 - rarely<br>3 - sometimes<br>4 - often<br>5 - very often | | 330 |
| Questionnaire | korff pain intensity | Average rating (1-10) from 3 Korff questionnaire questions:<br>Q2 - How would you classify your pain as it is at this moment?<br>Q3 - If you think about the days you have been in pain in the last three months, how would you rate your worst pain?<br>Q4 - If you think about the days you have had pain in the last three months, how would you rate the average intensity of the pain? | | 1:10 | 307 |

| | | | | | |
|---|---|---|---|---|---|
| Questionnaire | korff disability | Average rating (1-10) from 4 Korff questionnaire questions:<br>Q5 - To what extent has your pain affected your daily activities in the last three months?<br>Q6 - To what extent has the pain affected your ability to participate in family or leisure activities in the last three months?<br>Q7 - To what extent has the pain affected your ability to do your work/homework in the last three months?<br>Q8 - To what extent has the pain affected your physical activities in your leisure time in the last three months? | | 1:10 | 327 |
| Questionnaire | korff disability grading | Korff disability grading:<br>< 3 -> 0<br>3 - 5 -> 1<br>5-7 -> 2<br>> 7 -> 3 | 0-3 | | 327 |
| Questionnaire | korff days grading | Grading of days not able to carry out regular activities in the past 3 months:<br>< 4 -> 0<br>4 - 8 -> 1<br>8 - 16 -> 2<br>> 16 -> 3 | 0-3 | | 306 |
| Questionnaire | korff disability score | Summation score Korff disability grading and Korff days garding | 0-6 | | 335 |
| Questionnaire | korff grading | Grading based on combination of Korff pain intensity (PI) and disability score (DS):<br>PI < 50 & DS < 3 -> 1<br>PI > 50 & DS < 3 -> 2<br>PI > 50 & DS 3 - 5 -> 3<br>PI > 50 & DS > 5 -> 4 | 1-4 | | 335 |
| Questionnaire | RM disability | Rolland Morris questionnaire summation score across 26 questions:<br>1 - True<br>0 - false | 0-26 | | 305 |
| MRI | intervertebral disc oberservers | Intravertabral disc Pfirrmann classification rater code | 1-3 | | 408 |

| | | | | |
|---|---|---|---|---|
| MRI | intervertebral disc degeneration L1 - L2 | Intravertabral disc L1-L2 Pfirrmann classification score | 1-5 | 431 |
| MRI | intervertebral disc degeneration L2 - L3 | Intravertabral disc L2-L3 Pfirrmann classification score | 1-5 | 431 |
| MRI | intervertebral disc degeneration L3 - L4 | Intravertabral disc L3-L4 Pfirrmann classification score | 1-5 | 431 |
| MRI | intervertebral disc degeneration L4 - L5 | Intravertabral disc L4-L5 Pfirrmann classification score | 1-5 | 431 |
| MRI | intervertebral disc degeneration L5 - S1 | Intravertabral disc L5-S1 Pfirrmann classification score | 1-5 | 431 |
| MRI | intervertebral disc degeneration L6 - S1 | Intravertabral disc L6-S1 Pfirrmann classification score | 1-5 | 1287 |
| MRI | intervertebral disc herniation oberservers | intervertebral disc Kramer classification rater code | 1-3 | 408 |
| MRI | intervertebral disc herniation L1 - L2 | intervertebral disc L1-L2 Kramer classification score | 1-5 | 431 |
| MRI | intervertebral disc herniation L2 - L3 | intervertebral disc L2-L3 Kramer classification score | 1-5 | 431 |
| MRI | intervertebral disc herniation L3 - L4 | intervertebral disc L3-L4 Kramer classification score | 1-5 | 431 |
| MRI | intervertebral disc herniation L4 - L5 | intervertebral disc L4-L5 Kramer classification score | 1-5 | 431 |
| MRI | intervertebral disc herniation L5 - S1 | intervertebral disc L5-S1 Kramer classification score | 1-5 | 431 |
| MRI | intervertebral disc herniation L6 - S1 | intervertebral disc L6-S1 Kramer classification score | 1-5 | 1288 |
| MRI | facet joint oberservers | Facet joint arthosis with Fujiwara classification rate code | 1-3 | 408 |
| MRI | facet joint L1 - L2 left | Facet joint L1-L2 (left) arthosis Fujiwara classification score | 1-4 | 432 |
| MRI | facet joint L1 - L2 right | Facet joint L1-L2 (right) arthosis Fujiwara classification score | 1-4 | 432 |
| MRI | facet joint L2 - L3 left | Facet joint L2-L3 (left) arthosis Fujiwara classification score | 1-4 | 432 |
| MRI | facet joint L2 - L3 right | Facet joint L2-L3 (right) arthosis Fujiwara classification score | 1-4 | 432 |
| MRI | facet joint L3 - L4 left | Facet joint L3-L4 (left) arthosis Fujiwara classification score | 1-4 | 431 |
| MRI | facet joint L3 - L4 right | Facet joint L3-L4 (right) arthosis Fujiwara classification score | 1-4 | 431 |

| | | | | |
|---|---|---|---|---|
| MRI | facet joint L4 - L5 left | Facet joint L4-L5 (left) arthosis Fujiwara classification score | 1-4 | 431 |
| MRI | facet joint L4 - L5 right | Facet joint L4-L5 (right) arthosis Fujiwara classification score | 1-4 | 431 |
| MRI | facet joint L5/S1 - L5/L6 left | Facet joint L5/S1-L5/L6 (left) arthosis Fujiwara classification score | 1-4 | 431 |
| MRI | facet joint L5/S1 - L5/L6 right | Facet joint L5/S1-L5/L6 (right) arthosis Fujiwara classification score | 1-4 | 431 |
| MRI | facet joint L6 - S1 left | Facet joint L6-S1 (left) arthosis Fujiwara classification score | 1-4 | 1287 |
| MRI | facet joint L6 - S1 right | Facet joint L6-S1 (right) arthosis Fujiwara classification score | 1-4 | 1287 |
| MRI | osteochondrosis intervertebralis oberservers | osteochondrosis intervertebralis rater | 1-3 | 408 |
| MRI | osteochondrosis intervertebralis L1 - L2 | osteochondrosis intervertebralis L1-L2 score | 0-2 | 431 |
| MRI | osteochondrosis intervertebralis L2 - L3 | osteochondrosis intervertebralis L2 L3 score | 0-2 | 431 |
| MRI | osteochondrosis intervertebralis L3 - L4 | osteochondrosis intervertebralis L3-L4 score | 0-2 | 431 |
| MRI | osteochondrosis intervertebralis L4 - L5 | osteochondrosis intervertebralis L4-L5 score | 0-2 | 431 |
| MRI | osteochondrosis intervertebralis L5 - S1 | osteochondrosis intervertebralis L5-S1 score | 0-2 | 431 |
| MRI | osteochondrosis intervertebralis L6 - S1 | osteochondrosis intervertebralis L6-S1 score | 0-2 | 1295 |
| MRI | spinal canal stenosis oberservers | spinal canal stenosis rater | 1-3 | 408 |
| MRI | spinal canal stenosis L1 | spinal canal stenosis diameter | 10-27mm | 431 |
| MRI | spinal canal stenosis L1 | spinal canal stenosis diameter | 10-27mm | 431 |
| MRI | spinal canal stenosis L2 | spinal canal stenosis diameter | 10-27mm | 431 |
| MRI | spinal canal stenosis L3 | spinal canal stenosis diameter | 10-27mm | 431 |
| MRI | spinal canal stenosis L4 | spinal canal stenosis diameter | 10-27mm | 431 |
| MRI | spinal canal stenosis L5 | spinal canal stenosis diameter | 10-27mm | 1289 |
| MRI | schizas oberservers | schizas spinal canal rater | 1-3 | 408 |
| MRI | schizas L1 | schizas spinal canal L1 | 1-3 | 1004 |

| | | | | |
|---|---|---|---|---|
| MRI | schizas L2 | schizas spinal canal L2 | 1-3 | 1004 |
| MRI | schizas L3 | schizas spinal canal L3 | 1-3 | 1004 |
| MRI | schizas L4 | schizas spinal canal L4 | 1-3 | 1004 |
| MRI | schizas L5 | schizas spinal canal L5 | 1-3 | 1004 |
| MRI | schizas L6 | schizas spinal canal L6 | 1-3 | 1296 |
| MRI | lordosis oberservers | Lordosis grading rater | 1-3 | 408 |
| MRI | lordosis L1 - S1 | Lordosis L1-S1 | degrees | 529 |
| MRI | lordosis L1 - L2 | Lordosis L1-L2 | degrees | 529 |
| MRI | lordosis L2 - L3 | Lordosis L2-L3 | degrees | 529 |
| MRI | lordosis L3 - L4 | Lordosis L3-L4 | degrees | 529 |
| MRI | lordosis L4 - L5 | Lordosis L4-L5 | degrees | 529 |
| MRI | lordosis L5 - S1 | Lordosis L5-L6 | degrees | 529 |
| MRI | lordosis L6 - S1 | Lordosis L6-S1 | degrees | 1290 |
| Back shape & function | sacral frontal sit straight | sacral sipine segment angle during frontal sit straight motion | degrees | 86 |
| Back shape & function | thoracic value frontal sit straight | thoracic spine segment angle during frontal sit straight motion | degrees | 86 |
| Back shape & function | lumbar value frontal sit straight | lumbar spine segment angle during frontal sit straight motion | degrees | 86 |
| Back shape & function | inclination value frontal sit straight | inlcination value during frontal sit straight motion | degrees | 86 |
| Back shape & function | spine length frontal sit straight | spine length measured by spine mouse during frontal sit straight motion | mm | 86 |
| Back shape & function | surface length frontal sit straight | spine surface area measured by spine mouse during frontal sit straight motion | mm2 | 86 |
| Back shape & function | sacral frontal sit left | sacral sipine segment angle during sit left motion | degrees | 86 |
| Back shape & function | thoracic value frontal sit left | thoracic spine segment angle during frontal sit left motion | degrees | 86 |
| Back shape & function | lumbar value frontal sit left | lumbar spine segment angle during frontal sit left motion | degrees | 86 |
| Back shape & function | inclination value frontal sit left | inlcination value during frontal sit left motion | degrees | 86 |
| Back shape & function | spine length frontal sit left | spine length measured by spine mouse during frontal sit left motion | mm | 86 |
| Back shape & function | surface length frontal sit left | spine surface area measured by spine mouse during frontal sit left motion | mm2 | 86 |
| Back shape & function | sacral frontal sit right | sacral sipine segment angle during sit right motion | degrees | 86 |
| Back shape & function | thoracic value frontal sit right | thoracic spine segment angle during frontal sit right motion | degrees | 86 |
| Back shape & function | lumbar value frontal sit right | lumbar spine segment angle during frontal sit right motion | degrees | 86 |

| | | | | |
|---|---|---|---|---|
| Back shape & function | inclination value frontal sit right | inlcination value during frontal sit right motion | degrees | 86 |
| Back shape & function | spine length frontal sit right | spine length measured by spine mouse during frontal sit right motion | mm | 86 |
| Back shape & function | surface length frontal sit right | spine surface area measured by spine mouse during frontal sit right motion | mm2 | 86 |
| Back shape & function | sacral frontal stand straight | sacral sipine segment angle during stand straight motion | degrees | 86 |
| Back shape & function | thoracic value frontal stand straight | thoracic spine segment angle during frontal stand straight motion | degrees | 86 |
| Back shape & function | lumbar value frontal stand straight | lumbar spine segment angle during frontal stand straight motion | degrees | 86 |
| Back shape & function | inclination value frontal stand straight | inlcination value during frontal stand straight motion | degrees | 86 |
| Back shape & function | spine length frontal stand straight | spine length measured by spine mouse during frontal stand straight motion | mm | 86 |
| Back shape & function | surface length frontal stand straight | spine surface area measured by spine mouse during frontal stand straight motion | mm2 | 86 |
| Back shape & function | sacral frontal stand left | sacral sipine segment angle during stand left motion | degrees | 86 |
| Back shape & function | thoracic value frontal stand left | thoracic spine segment angle during frontal stand left motion | degrees | 86 |
| Back shape & function | lumbar value frontal stand left | lumbar spine segment angle during frontal stand left motion | degrees | 86 |
| Back shape & function | inclination value frontal stand left | inlcination value during frontal stand left motion | degrees | 86 |
| Back shape & function | spine length frontal stand left | spine length measured by spine mouse during frontal stand left motion | mm | 86 |
| Back shape & function | surface length frontal stand left | spine surface area measured by spine mouse during frontal stand left motion | mm2 | 86 |
| Back shape & function | sacral frontal stand right | sacral sipine segment angle during stand right motion | degrees | 86 |
| Back shape & function | thoracic value frontal stand right | thoracic spine segment angle during frontal stand right motion | degrees | 86 |
| Back shape & function | lumbar value frontal stand right | lumbar spine segment angle during frontal stand right motion | degrees | 86 |
| Back shape & function | inclination value frontal stand right | inlcination value during frontal stand right motion | degrees | 86 |

| | | | | |
|---|---|---|---|---|
| Back shape & function | spine length frontal stand right | spine length measured by spine mouse during frontal stand right motion | mm | 86 |
| Back shape & function | surface length frontal stand right | spine surface area measured by spine mouse during frontal stand right motion | mm2 | 86 |
| Back shape & function | sacral sagittal sit straight | sacral sipine segment angle during sagittal sit straight motion | degrees | 86 |
| Back shape & function | thoracic value sagittal sit straight | thoracic spine segment angle during sagittal sit straight motion | degrees | 86 |
| Back shape & function | lumbar value sagittal sit straight | lumbar spine segment angle during sagittal sit straight motion | degrees | 86 |
| Back shape & function | inclination value sagittal sit straight | inlcination value during sagittal sit straight motion | degrees | 86 |
| Back shape & function | spine length sagittal sit straight | spine length measured by spine mouse during frontal sit straight motion | mm | 86 |
| Back shape & function | surface length sagittal sit straight | spine surface area measured by spine mouse during frontal sit straight motion | mm2 | 86 |
| Back shape & function | sacral sagittal sit extension | sacral sipine segment angle during sagittal sit extension motion | degrees | 86 |
| Back shape & function | thoracic value sagittal sit extension | thoracic spine segment angle during sagittal sit extension motion | degrees | 86 |
| Back shape & function | lumbar value sagittal sit extension | lumbar spine segment angle during sagittal sit extension motion | degrees | 86 |
| Back shape & function | inclination value sagittal sit extension | inlcination value during sagittal sit extension motion | degrees | 86 |
| Back shape & function | spine length sagittal sit extension | spine length measured by spine mouse during frontal sit extension motion | mm | 86 |
| Back shape & function | surface length sagittal sit extension | spine surface area measured by spine mouse during frontal sit extension motion | mm2 | 86 |
| Back shape & function | sacral sagittal sit flexion | sacral sipine segment angle during sagittal sit flexion motion | degrees | 86 |
| Back shape & function | thoracic value sagittal sit flexion | thoracic spine segment angle during sagittal sit flexion motion | degrees | 86 |
| Back shape & function | lumbar value sagittal sit flexion | lumbar spine segment angle during sagittal sit flexion motion | degrees | 86 |

| | | | | |
|---|---|---|---|---|
| Back shape & function | inclination value sagittal sit flexion | inlcination value during sagittal sit flexion motion | degrees | 86 |
| Back shape & function | spine length sagittal sit flexion | spine length measured by spine mouse during frontal sit flexion motion | mm | 86 |
| Back shape & function | surface length sagittal sit flexion | spine surface area measured by spine mouse during frontal sit flexion motion | mm2 | 86 |
| Back shape & function | sacral sagittal stand straight | sacral sipine segment angle during sagittal stand straight motion | degrees | 86 |
| Back shape & function | thoracic value sagittal stand straight | thoracic spine segment angle during sagittal stand straight motion | degrees | 86 |
| Back shape & function | lumbar value sagittal stand straight | lumbar spine segment angle during sagittal stand straight motion | degrees | 86 |
| Back shape & function | inclination value sagittal stand straight | inlcination value during sagittal stand straight motion | degrees | 86 |
| Back shape & function | spine length sagittal stand straight | spine length measured by spine mouse during sagittal stand straight motion | mm | 86 |
| Back shape & function | surface length sagittal stand straight | spine surface area measured by spine mouse during sagittal stand straight motion | mm2 | 86 |
| Back shape & function | sacral sagittal stand extension | sacral sipine segment angle during sagittal stand extension motion | degrees | 86 |
| Back shape & function | thoracic value sagittal stand extension | thoracic spine segment angle during sagittal stand extension motion | degrees | 86 |
| Back shape & function | lumbar value sagittal stand extension | lumbar spine segment angle during sagittal stand extension motion | degrees | 86 |
| Back shape & function | inclination value sagittal stand extension | inlcination value during sagittal stand extension motion | degrees | 86 |
| Back shape & function | spine length sagittal stand extension | spine length measured by spine mouse during sagittal stand extension motion | mm | 86 |
| Back shape & function | surface length sagittal stand extension | spine surface area measured by spine mouse during sagittal stand extension motion | mm2 | 86 |
| Back shape & function | sacral sagittal stand flexion | sacral sipine segment angle during sagittal stand flexion motion | degrees | 86 |

| | | | | |
|---|---|---|---|---|
| Back shape & function | thoracic value sagittal stand flexion | thoracic spine segment angle during sagittal stand flexion motion | degrees | 86 |
| Back shape & function | lumbar value sagittal stand flexion | lumbar spine segment angle during sagittal stand flexion motion | degrees | 86 |
| Back shape & function | inclination value sagittal stand flexion | inlcination value during sagittal stand flexion motion | degrees | 86 |
| Back shape & function | spine length sagittal stand flexion | spine length measured by spine mouse during sagittal stand flexion motion | mm | 86 |
| Back shape & function | surface length sagittal stand flexion | spine surface area measured by spine mouse during sagittal stand flexion motion | mm2 | 86 |

Supplementary Table S20. List of all variables removed during preprocessing and the reason.

| Modality | Variable | Reason |
|---|---|---|
| Demographic | height | BMI was used instead of this related variable |
| Demographic | hip | BMI was used instead of this related variable |
| Demographic | LBP pain duration | Question only asked to LBP patients |
| Demographic | LBP past pain duration | Question only asked to LBP patients |
| Demographic | leg left | Not relevant to LBP classification as used for movement analyses |
| Demographic | leg right | Not relevant to LBP classification as used for movement analyses |
| Demographic | question chronic LBP | Clinical diagnosis was used as target |
| Demographic | right left leg ratio | Not relevant to LBP classification as used for movement analyses |
| Demographic | waist | BMI was used instead of this related variable |
| Demographic | weight | BMI was used instead of this related variable |
| Demographic | waist hip ratio | BMI was used instead of this related variable |
| Questionnaire | medication 1 demand | Medication clinical questions heavily biased towards LBP patients |
| Questionnaire | medication 1 demand dosis mg | Medication clinical questions heavily biased towards LBP patients |
| Questionnaire | medication 1 demand month | Medication clinical questions heavily biased towards LBP patients |
| Questionnaire | medication 1 dosis mg | Medication clinical questions heavily biased towards LBP patients |
| Questionnaire | medication 1 midday | Medication clinical questions heavily biased towards LBP patients |
| Questionnaire | medication 1 morning | Medication clinical questions heavily biased towards LBP patients |
| Questionnaire | medication 1 noon | Medication clinical questions heavily biased towards LBP patients |
| Questionnaire | medication 1 registration | Medication clinical questions heavily biased towards LBP patients |

| | | |
|---|---|---|
| Questionnaire | medication 2 demand | Medication clinical questions heavily biased towards LBP patients |
| Questionnaire | medication 2 demand dosis mg | Medication clinical questions heavily biased towards LBP patients |
| Questionnaire | medication 2 demand month | Medication clinical questions heavily biased towards LBP patients |
| Questionnaire | medication 2 dosis mg | Medication clinical questions heavily biased towards LBP patients |
| Questionnaire | medication 2 midday | Medication clinical questions heavily biased towards LBP patients |
| Questionnaire | medication 2 morning | Medication clinical questions heavily biased towards LBP patients |
| Questionnaire | medication 2 noon | Medication clinical questions heavily biased towards LBP patients |
| Questionnaire | medication 2 registration | Medication clinical questions heavily biased towards LBP patients |
| Questionnaire | pain arm | Pain questions and tests on back heavily biased towards LBP pateints |
| Questionnaire | pain begin | Pain questions and tests on back heavily biased towards LBP pateints |
| Questionnaire | pain coughing sneezing | Pain questions and tests on back heavily biased towards LBP pateints |
| Questionnaire | pain day diff | Pain questions and tests on back heavily biased towards LBP pateints |
| Questionnaire | pain heel drop | Pain questions and tests on back heavily biased towards LBP pateints |
| Questionnaire | pain jumping | Pain questions and tests on back heavily biased towards LBP pateints |
| Questionnaire | pain knocking | Pain questions and tests on back heavily biased towards LBP pateints |
| Questionnaire | pain leg | Pain questions and tests on back heavily biased towards LBP pateints |
| Questionnaire | pain mennell sign | Pain questions and tests on back heavily biased towards LBP pateints |
| Questionnaire | pain movment limit | Pain questions and tests on back heavily biased towards LBP pateints |
| Questionnaire | pain paralysis | Pain questions and tests on back heavily biased towards LBP pateints |
| Questionnaire | pain paralysis location | Pain questions and tests on back heavily biased towards LBP pateints |
| Questionnaire | pain pressure | Pain questions and tests on back heavily biased towards LBP pateints |
| Questionnaire | pain primary location | Pain questions and tests on back heavily biased towards LBP pateints |
| Questionnaire | pain resting | Pain questions and tests on back heavily biased towards LBP pateints |
| Questionnaire | pain secondary location | Pain questions and tests on back heavily biased towards LBP pateints |
| Questionnaire | pain sensitivity | Pain questions and tests on back heavily biased towards LBP pateints |
| Questionnaire | pain sitting | Pain questions and tests on back heavily biased towards LBP pateints |
| Questionnaire | pain spinous process shake | Pain questions and tests on back heavily biased towards LBP pateints |

| | | |
|---|---|---|
| Questionnaire | pain standing | Pain questions and tests on back heavily biased towards LBP pateints |
| Questionnaire | pain walking | Pain questions and tests on back heavily biased towards LBP pateints |
| Questionnaire | pain walking distance | Pain questions and tests on back heavily biased towards LBP pateints |
| Questionnaire | pain when strongest | Pain questions and tests on back heavily biased towards LBP pateints |
| Questionnaire | prior diagnosis | Prior diagnoses and surgery clincal questions bias towards LBP pateints |
| Questionnaire | prior diagnosis back level | Prior diagnoses and surgery clincal questions bias towards LBP pateints |
| Questionnaire | prior diagnosis which | Prior diagnoses and surgery clincal questions bias towards LBP pateints |
| Questionnaire | prior general disease | Prior diagnoses and surgery clincal questions bias towards LBP pateints |
| Questionnaire | prior surgeries spine level | Prior diagnoses and surgery clincal questions bias towards LBP pateints |
| Questionnaire | prior surgeries which | Very high correlation (> 0.9) |
| Questionnaire | alcohol restrict | Very low variance (0.48) |
| Questionnaire | body self assessment | Questions biased towards LBP patients |
| Questionnaire | job duration years | Job duration has high correlation with age |
| Questionnaire | job load 2 | Questions with large amount of missing values |
| Questionnaire | job load 2 duration | Questions with large amount of missing values |
| Questionnaire | job posture duration | Questions with large amount of missing values |
| Questionnaire | smoking | Very high correlation (> 0.9) |
| Questionnaire | chron 1 | Questions only asked of LBP patients |
| Questionnaire | chron 2 | Questions only asked of LBP patients |
| Questionnaire | chron 3 | Questions only asked of LBP patients |
| Questionnaire | employment currently | Questions only asked of LBP patients |
| Questionnaire | employment disability | Questions only asked of LBP patients |
| Questionnaire | employment disability grade | Questions only asked of LBP patients |
| Questionnaire | employment pension | Questions only asked of LBP patients |
| Questionnaire | employment pension application | Questions only asked of LBP patients |
| Questionnaire | employment pension application not decided | Questions only asked of LBP patients |
| Questionnaire | employment pension application opposition | Questions only asked of LBP patients |
| Questionnaire | employment pension application rejected | Questions only asked of LBP patients |
| Questionnaire | employment pension final | Questions only asked of LBP patients |
| Questionnaire | employment pension for time | Questions only asked of LBP patients |
| Questionnaire | employment pension type | Questions only asked of LBP patients |
| Questionnaire | employment unable work | Questions only asked of LBP patients |
| Questionnaire | employment unable work could return | Questions only asked of LBP patients |
| Questionnaire | employment unable work last 3 months | Questions only asked of LBP patients |
| Questionnaire | fabq physical activity | Questions only asked of LBP patients |
| Questionnaire | fabq workload | Questions only asked of LBP patients |
| Questionnaire | ipaq met moderate | Only used IPAQ sum score and not individual sub-scores |

| | | |
|---|---|---|
| Questionnaire | ipaq met sitting | Only used IPAQ sum score and not individual sub-scores |
| Questionnaire | ipaq met vigorous | Only used IPAQ sum score and not individual sub-scores |
| Questionnaire | ipaq met walking | Only used IPAQ sum score and not individual sub-scores |
| Questionnaire | kinesiophobia | Questions only asked of LBP patients |
| Questionnaire | korff days grading | Questions only asked of LBP patients |
| Questionnaire | korff disability | Questions only asked of LBP patients |
| Questionnaire | korff disability grading | Questions only asked of LBP patients |
| Questionnaire | korff disability score | Questions only asked of LBP patients |
| Questionnaire | korff grading | Questions only asked of LBP patients |
| Questionnaire | korff pain intensity | Questions only asked of LBP patients |
| Questionnaire | RM disability | Questions only asked of LBP patients |
| Questionnaire | SF36 health perception | Questions biased towards LBP patients |
| Questionnaire | SF36 physical function | Questions biased towards LBP patients |
| Questionnaire | SF36 physical pain | Questions biased towards LBP patients |
| Questionnaire | SF36 physical role function | Questions biased towards LBP patients |
| Questionnaire | SF36 vitality | Questions biased towards LBP patients |
| Questionnaire | stress days | Questions only asked of LBP patients |
| Questionnaire | stress score | Questions only asked of LBP patients |
| Questionnaire | therapy 1 | Questions biased towards LBP patients |
| Questionnaire | therapy 10 | Questions biased towards LBP patients |
| Questionnaire | therapy 10 1 | Questions biased towards LBP patients |
| Questionnaire | therapy 11 | Questions biased towards LBP patients |
| Questionnaire | therapy 11 1 | Questions biased towards LBP patients |
| Questionnaire | therapy 12 | Questions biased towards LBP patients |
| Questionnaire | therapy 12 1 | Questions biased towards LBP patients |
| Questionnaire | therapy 13 | Questions biased towards LBP patients |
| Questionnaire | therapy 13 1 | Questions biased towards LBP patients |
| Questionnaire | therapy 14 | Questions biased towards LBP patients |
| Questionnaire | therapy 14 1 | Questions biased towards LBP patients |
| Questionnaire | therapy 15 | Questions biased towards LBP patients |
| Questionnaire | therapy 15 1 | Questions biased towards LBP patients |
| Questionnaire | therapy 2 | Questions biased towards LBP patients |
| Questionnaire | therapy 2 1 | Questions biased towards LBP patients |
| Questionnaire | therapy 3 | Questions biased towards LBP patients |
| Questionnaire | therapy 3 1 | Questions biased towards LBP patients |
| Questionnaire | therapy 4 | Questions biased towards LBP patients |
| Questionnaire | therapy 4 1 | Questions biased towards LBP patients |
| Questionnaire | therapy 5 | Questions biased towards LBP patients |
| Questionnaire | therapy 5 1 | Questions biased towards LBP patients |
| Questionnaire | therapy 6 | Questions biased towards LBP patients |
| Questionnaire | therapy 6 1 | Questions biased towards LBP patients |
| Questionnaire | therapy 7 | Questions biased towards LBP patients |
| Questionnaire | therapy 7 1 | Questions biased towards LBP patients |
| Questionnaire | therapy 8 | Questions biased towards LBP patients |
| Questionnaire | therapy 8 1 | Questions biased towards LBP patients |
| Questionnaire | therapy 9 | Questions biased towards LBP patients |

| | | |
|---|---|---|
| Questionnaire | therapy 9 1 | Questions biased towards LBP patients |
| Clinical | 3 spine mouse measure | Conducting spine mouse 3x not related to LBP |
| Clinical | back head wall distance | Near zero variance (< 0.1) |
| Clinical | heel stand | Near zero variance (< 0.1) |
| Clinical | hip flexion | Large amount of missing values (> 250) |
| Clinical | toe stand | Near zero variance (< 0.1) |
| Clinical | breath rate | Heart rate, breath rate, and blood pressure not related to LBP |
| Clinical | heart diastolic pressure | Heart rate, breath rate, and blood pressure not related to LBP |
| Clinical | heart rate | Heart rate, breath rate, and blood pressure not related to LBP |
| Clinical | heart systolic pressure | Heart rate, breath rate, and blood pressure not related to LBP |
| Clinical | coronal balance | Near zero variance (< 0.1) |
| Clinical | gait pattern | Near zero variance (< 0.1) |
| Clinical | pseudo lasegue left | Large amount of missing values (> 250) |
| Clinical | pseudo lasegue right | Large amount of missing values (> 250) |
| Clinical | reflex achilles left | Near zero variance (< 0.1) |
| Clinical | reflex achilles right | Near zero variance (< 0.1) |
| Clinical | reflex quadricep left | Near zero variance (< 0.1) |
| Clinical | reflex quadricep right | Near zero variance (< 0.1) |
| Clinical | straight leg raise | Near zero variance (< 0.1) |
| Clinical | trendelenburg sign | Near zero variance (< 0.1) |
| Back shape & function | lumbar value sagittal stand flexion | High correlation (> 0.9) |
| Back shape & function | spine length frontal sit left | High correlation (> 0.9) |
| Back shape & function | spine length frontal sit right | High correlation (> 0.9) |
| Back shape & function | spine length frontal sit straight | High correlation (> 0.9) |
| Back shape & function | spine length frontal stand left | High correlation (> 0.9) |
| Back shape & function | spine length frontal stand right | High correlation (> 0.9) |
| Back shape & function | spine length frontal stand straight | High correlation (> 0.9) |
| Back shape & function | spine length sagittal sit extension | High correlation (> 0.9) |
| Back shape & function | spine length sagittal sit flexion | High correlation (> 0.9) |
| Back shape & function | spine length sagittal sit straight | High correlation (> 0.9) |
| Back shape & function | spine length sagittal stand flexion | High correlation (> 0.9) |
| Back shape & function | spine length sagittal stand straight | High correlation (> 0.9) |
| Back shape & function | surface length frontal sit left | High correlation (> 0.9) |
| Back shape & function | surface length frontal sit right | High correlation (> 0.9) |

| | | |
|---|---|---|
| Back shape & function | surface length frontal sit straight | High correlation (> 0.9) |
| Back shape & function | surface length frontal stand left | High correlation (> 0.9) |
| Back shape & function | surface length frontal stand right | High correlation (> 0.9) |
| Back shape & function | surface length frontal stand straight | High correlation (> 0.9) |
| Back shape & function | surface length sagittal sit extension | High correlation (> 0.9) |
| Back shape & function | surface length sagittal sit flexion | High correlation (> 0.9) |
| Back shape & function | surface length sagittal sit straight | High correlation (> 0.9) |
| Back shape & function | surface length sagittal stand extension | High correlation (> 0.9) |
| Back shape & function | surface length sagittal stand flexion | High correlation (> 0.9) |
| Back shape & function | surface length sagittal stand straight | High correlation (> 0.9) |
| MRI | intervertebral disc herniation L1 - L2 | Near zero variance (< 0.2) |
| MRI | intervertebral disc herniation L6 - S1 | Very few (< 10) subjects had values for L6-S1 |
| MRI | intervertebral disc herniation oberservers | radiologist rater info not relevant |
| MRI | facet joint L6 - S1 left | Very few (< 10) subjects had values for L6-S1 |
| MRI | facet joint L6 - S1 right | Very few (< 10) subjects had values for L6-S1 |
| MRI | facet joint oberservers | radiologist rater info not relevant |
| MRI | intervertebral disc degeneration L6 - S1 | Very few (< 10) subjects had values for L6-S1 |
| MRI | intervertebral disc oberservers | radiologist rater info not relevant |
| MRI | lordosis L1 - S1 | Lordosis rating has > 100 missing values compared to other ratings |
| MRI | lordosis L1 - L2 | Lordosis rating has > 100 missing values compared to other ratings |
| MRI | lordosis L2 - L3 | Lordosis rating has > 100 missing values compared to other ratings |
| MRI | lordosis L3 - L4 | Lordosis rating has > 100 missing values compared to other ratings |
| MRI | lordosis L4 - L5 | Lordosis rating has > 100 missing values compared to other ratings |
| MRI | lordosis L5 - S1 | Lordosis rating has > 100 missing values compared to other ratings |
| MRI | lordosis L6 - S1 | Very few (< 10) subjects had values for L6-S1 |
| MRI | lordosis oberservers | radiologist rater info not relevant |
| MRI | osteochondrosis intervertebralis L1 - L2 | Near zero variance (< 0.2) |
| MRI | osteochondrosis intervertebralis L2 - L3 | Near zero variance (< 0.2) |
| MRI | osteochondrosis intervertebralis L3 - L4 | Near zero variance (< 0.2) |
| MRI | osteochondrosis intervertebralis L6 - S1 | Very few (< 10) subjects had values for L6-S1 |

| | | |
|---|---|---|
| MRI | osteochondrosis intervertebralis oberservers | radiologist rater info not relevant |
| MRI | schizas L1 | Schizas rating present in < 10 subjects |
| MRI | schizas L2 | Schizas rating present in < 10 subjects |
| MRI | schizas L3 | Schizas rating present in < 10 subjects |
| MRI | schizas L4 | Schizas rating present in < 10 subjects |
| MRI | schizas L5 | Schizas rating present in < 10 subjects |
| MRI | schizas L6 | Very few (< 10) subjects had values for L6-S1 |
| MRI | schizas oberservers | radiologist rater info not relevant |
| MRI | spinal canal stenosis L6 | Very few (< 10) subjects had values for L6-S1 |
| MRI | spinal canal stenosis oberservers | radiologist rater info not relevant |

#### Supplementary Table S21: List of all variables remaing after preprocessing and cleaning for modelling

| Modality | Variable | Data Type |
|---|---|---|
| Demographic | age | continuous |
| Demographic | sex | nominal |
| Demographic | clinic chronic LBP [Target] | nominal |
| Demographic | BMI | continuous |
| Questionnaire | prior surgeries | nominal |
| Questionnaire | physical activity | ordinal |
| Questionnaire | covid19 activity | ordinal |
| Questionnaire | alcohol freq | ordinal |
| Questionnaire | alcohol glasses | ordinal |
| Questionnaire | smoking pack years | continuous |
| Questionnaire | family back pain | nominal |
| Questionnaire | job field | nominal |
| Questionnaire | job posture | nominal |
| Questionnaire | job load 1 | nominal |
| Questionnaire | job load 1 duration | continuous |
| Questionnaire | pyschological stress | nominal |
| Questionnaire | living situation | nominal |
| Questionnaire | family conflicts | nominal |
| Clinical | cervical inclination | continuous |
| Clinical | cervical reclination | continuous |
| Clinical | chin sternum distance | nominal |
| Clinical | cervical lat bend left | continuous |
| Clinical | cervical lat bend right | continuous |
| Clinical | cervical axial rotate left | continuous |
| Clinical | cervical axial rotate right | continuous |
| Clinical | thoracic lumbar inclination | continuous |
| Clinical | thoracic lumbar reclination | continuous |
| Clinical | finger floor distance | continuous |
| Clinical | thoracic lumbar lateral bend left | continuous |
| Clinical | thoracic lumbar lateral bend right | continuous |
| Clinical | thoracic lumbar axial rotation left | continuous |
| Clinical | thoracic lumbar axial rotation right | continuous |

| | | |
|---|---|---|
| Clinical | OTT | continuous |
| Clinical | shober | continuous |
| Clinical | square shoulders | nominal |
| Clinical | plump line | nominal |
| Clinical | rib hump | nominal |
| Clinical | lumbar bulge | nominal |
| Clinical | asym waist triangle | nominal |
| Clinical | back form | nominal |
| Clinical | roussoly type | nominal |
| Clinical | sagittal balance | nominal |
| Clinical | rigid muscle | nominal |
| Clinical | general mobility | nominal |
| Questionnaire | hip pain | nominal |
| Clinical | hip flexion left | continuous |
| Clinical | hip flexion right | continuous |
| Clinical | hip extension left | continuous |
| Clinical | hip extension right | continuous |
| Clinical | hip external rotate left | continuous |
| Clinical | hip external rotate right | continuous |
| Clinical | hip internal rotate left | continuous |
| Clinical | hip internal rotate right | continuous |
| Clinical | hip abduction left | continuous |
| Clinical | hip abduction right | continuous |
| Clinical | hip adduction left | continuous |
| Clinical | hip adduction right | continuous |
| Clinical | thomas handle left | nominal |
| Clinical | thomas handle right | nominal |
| Clinical | pelvic tilt frontal | nominal |
| Clinical | pelvic tilt sagittal | nominal |
| Clinical | sit to stand 30sec | continuous |
| Questionnaire | SF36 social function | continuous |
| Questionnaire | SF36 emotional role function | continuous |
| Questionnaire | SF36 psychological well being | continuous |
| Questionnaire | IPAQ met sum | continuous |
| Questionnaire | SRBAI sum | continuous |
| Questionnaire | BRSQ intrinsic motivation | continuous |
| Questionnaire | BRSQ integrated regulation | continuous |
| Questionnaire | BRSQ external regulation | continuous |
| MRI | intervertebral disc degeneration L1 - L2 | nominal |
| MRI | intervertebral disc degeneration L2 - L3 | nominal |
| MRI | intervertebral disc degeneration L3 - L4 | nominal |
| MRI | intervertebral disc degeneration L4 - L5 | nominal |
| MRI | intervertebral disc degeneration L5 - S1 | nominal |
| MRI | intervertebral disc herniation L2 - L3 | nominal |
| MRI | intervertebral disc herniation L3 - L4 | nominal |
| MRI | intervertebral disc herniation L4 - L5 | nominal |
| MRI | intervertebral disc herniation L5 - S1 | nominal |
| MRI | facet joint L1 - L2 left | nominal |
| MRI | facet joint L1 - L2 right | nominal |

| | | |
|---|---|---|
| MRI | facet joint L2 - L3 left | nominal |
| MRI | facet joint L2 - L3 right | nominal |
| MRI | facet joint L3 - L4 left | nominal |
| MRI | facet joint L3 - L4 right | nominal |
| MRI | facet joint L4 - L5 left | nominal |
| MRI | facet joint L4 - L5 right | nominal |
| MRI | facet joint L5/S1 - L5/L6 left | nominal |
| MRI | facet joint L5/S1 - L5/L6 right | nominal |
| MRI | osteochondrosis intervertebralis L4 - L5 | nominal |
| MRI | osteochondrosis intervertebralis L5 - S1 | nominal |
| MRI | spinal canal stenosis L1 | continuous |
| MRI | spinal canal stenosis L1 | continuous |
| MRI | spinal canal stenosis L2 | continuous |
| MRI | spinal canal stenosis L3 | continuous |
| MRI | spinal canal stenosis L4 | continuous |
| MRI | spinal canal stenosis L5 | continuous |
| Back shape & function | thoracic value frontal sit straight | continuous |
| Back shape & function | lumbar value frontal sit straight | continuous |
| Back shape & function | inclination value frontal sit straight | continuous |
| Back shape & function | sacral frontal sit left | continuous |
| Back shape & function | thoracic value frontal sit left | continuous |
| Back shape & function | lumbar value frontal sit left | continuous |
| Back shape & function | inclination value frontal sit left | continuous |
| Back shape & function | sacral frontal sit right | continuous |
| Back shape & function | thoracic value frontal sit right | continuous |
| Back shape & function | lumbar value frontal sit right | continuous |
| Back shape & function | inclination value frontal sit right | continuous |
| Back shape & function | sacral frontal stand straight | continuous |
| Back shape & function | thoracic value frontal stand straight | continuous |
| Back shape & function | lumbar value frontal stand straight | continuous |
| Back shape & function | inclination value frontal stand straight | continuous |
| Back shape & function | sacral frontal stand left | continuous |
| Back shape & function | thoracic value frontal stand left | continuous |
| Back shape & function | lumbar value frontal stand left | continuous |
| Back shape & function | inclination value frontal stand left | continuous |
| Back shape & function | sacral frontal stand right | continuous |
| Back shape & function | thoracic value frontal stand right | continuous |
| Back shape & function | lumbar value frontal stand right | continuous |
| Back shape & function | inclination value frontal stand right | continuous |
| Back shape & function | sacral sagittal sit straight | continuous |
| Back shape & function | thoracic value sagittal sit straight | continuous |
| Back shape & function | lumbar value sagittal sit straight | continuous |
| Back shape & function | inclination value sagittal sit straight | continuous |
| Back shape & function | sacral sagittal sit extension | continuous |
| Back shape & function | thoracic value sagittal sit extension | continuous |
| Back shape & function | lumbar value sagittal sit extension | continuous |
| Back shape & function | inclination value sagittal sit extension | continuous |
| Back shape & function | sacral sagittal sit flexion | continuous |

| | | |
|---|---|---|
| Back shape & function | thoracic value sagittal sit flexion | continuous |
| Back shape & function | lumbar value sagittal sit flexion | continuous |
| Back shape & function | inclination value sagittal sit flexion | continuous |
| Back shape & function | sacral sagittal stand straight | continuous |
| Back shape & function | thoracic value sagittal stand straight | continuous |
| Back shape & function | lumbar value sagittal stand straight | continuous |
| Back shape & function | inclination value sagittal stand straight | continuous |
| Back shape & function | sacral sagittal stand extension | continuous |
| Back shape & function | thoracic value sagittal stand extension | continuous |
| Back shape & function | lumbar value sagittal stand extension | continuous |
| Back shape & function | inclination value sagittal stand extension | continuous |
| Back shape & function | spine length sagittal stand extension | continuous |
| Back shape & function | sacral sagittal stand flexion | continuous |
| Back shape & function | thoracic value sagittal stand flexion | continuous |
| Back shape & function | inclination value sagittal stand flexion | continuous |

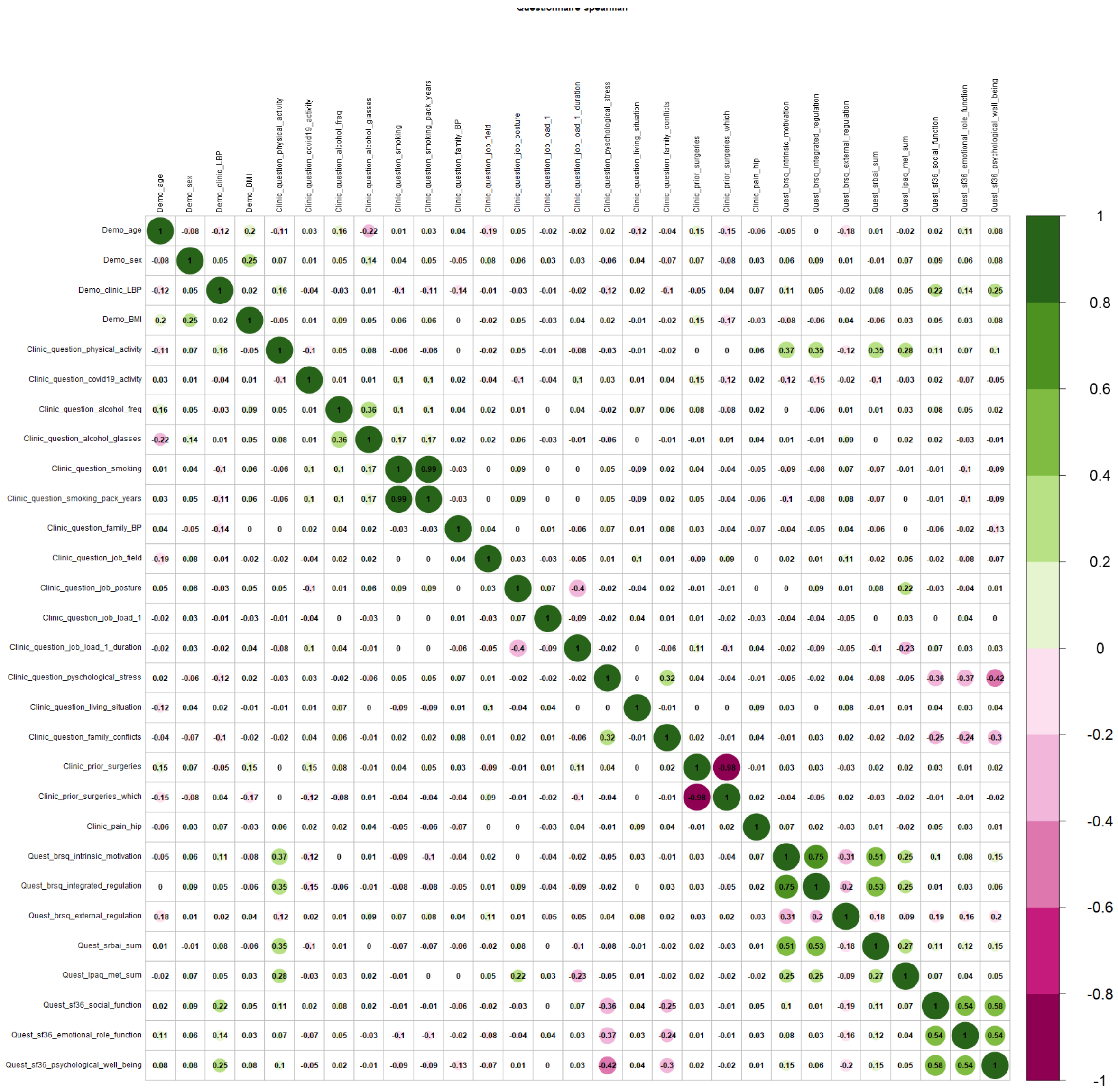

Supplementary Figure S1. Spearman correlation matrix of questionnaire dataset before removal of highly correlated variables

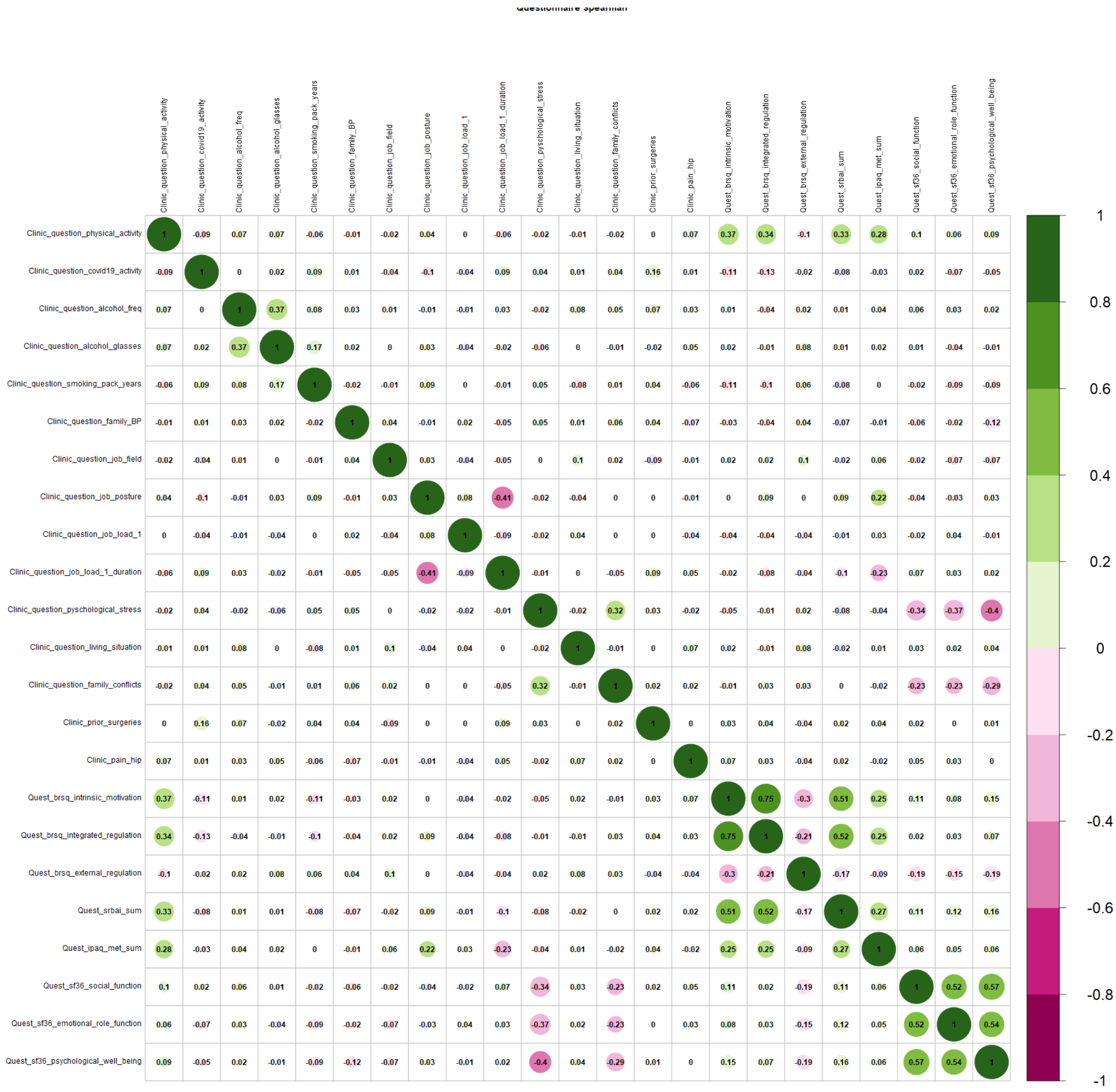

Supplementary Figure S2. Spearman correlation matrix of questionnaire dataset after removal of highly correlated variables

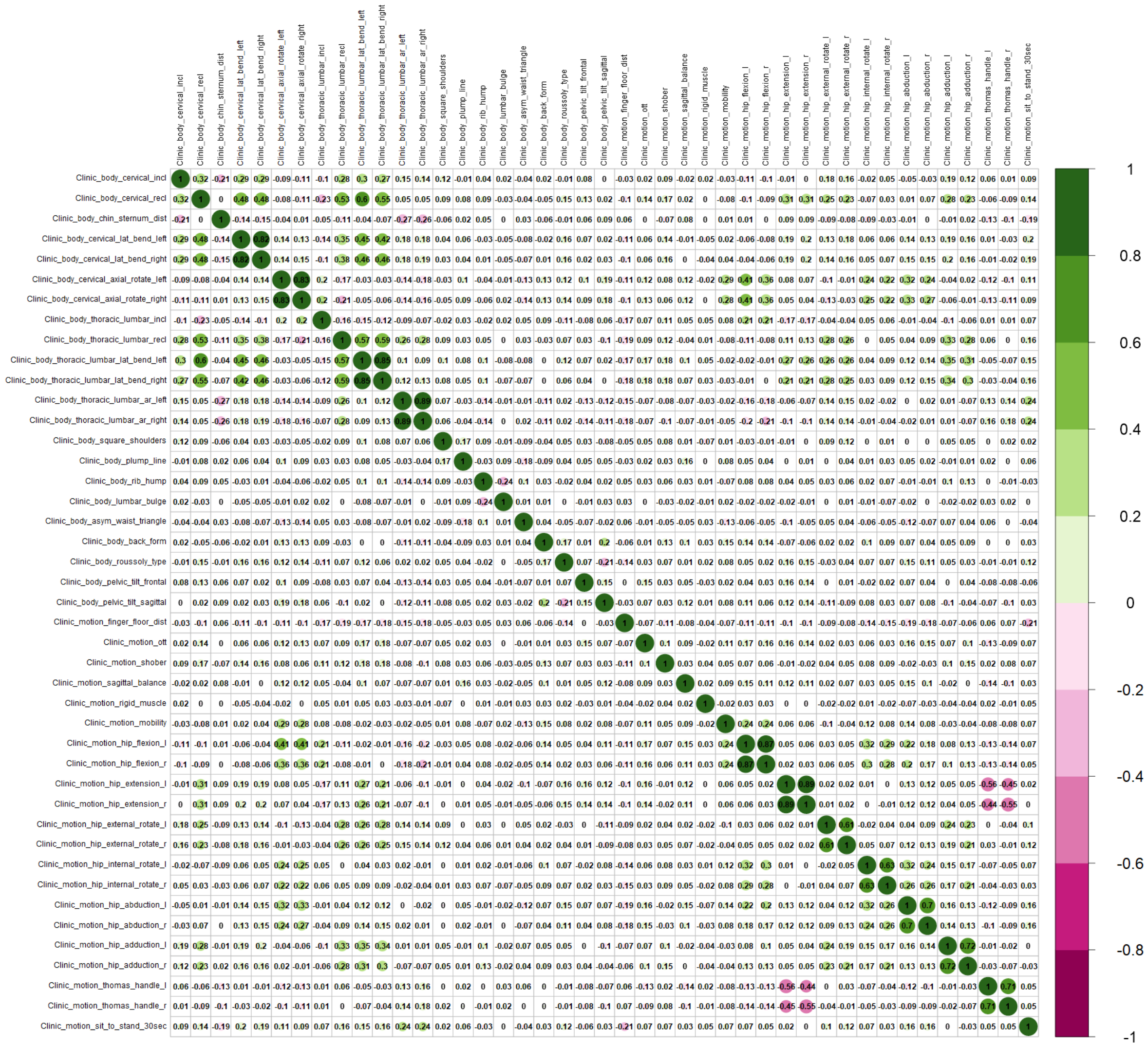

Supplementary Figure S3. Spearman correlation matrix of clinical assessment dataset

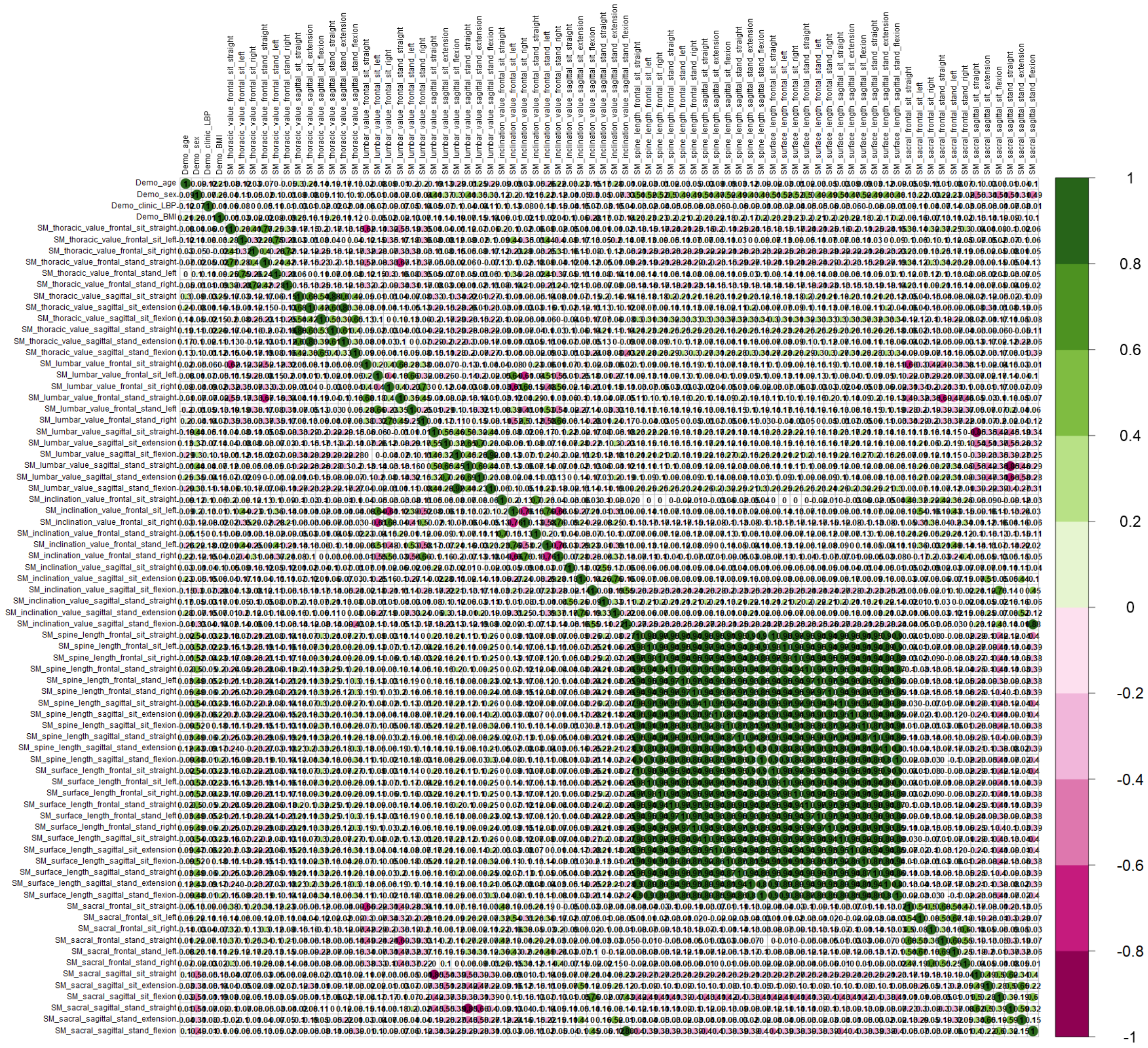

Supplementary Figure S4. Spearman correlation matrix of back shape and function dataset before removal of highly correlated variables

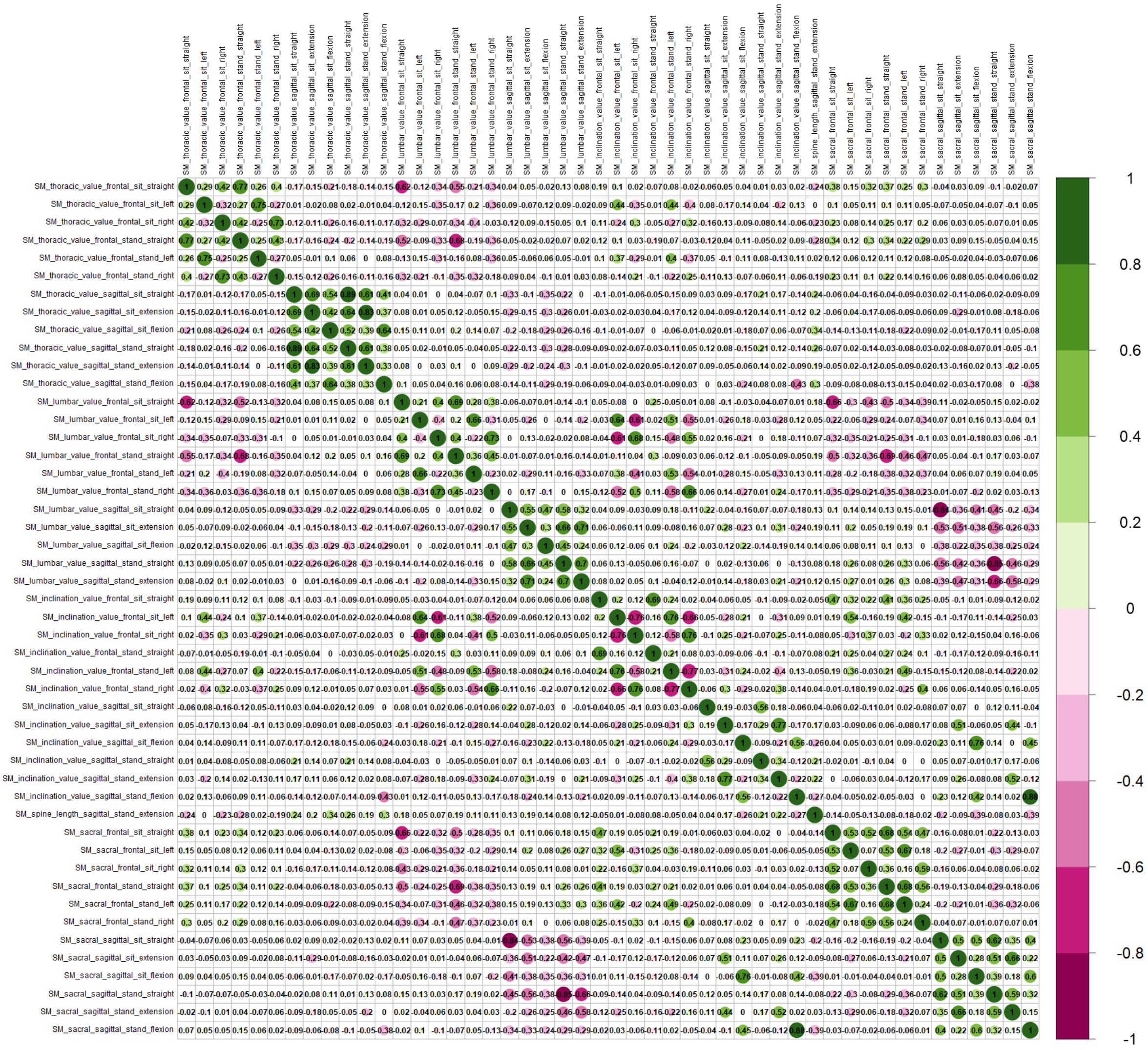

Supplementary Figure S5. Spearman correlation matrix of back shape and function dataset after removal of highly correlated variables

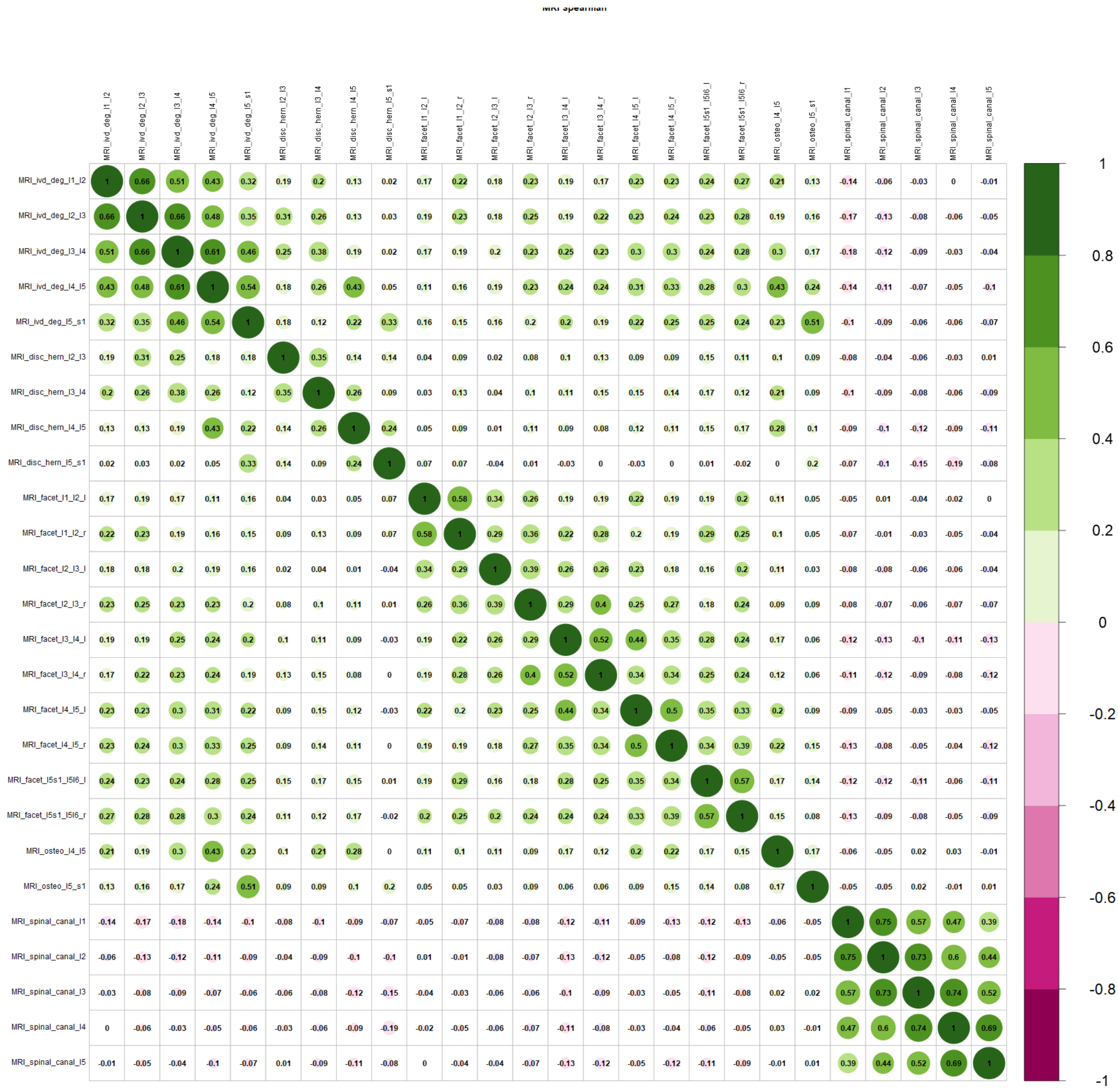

Supplementary Figure S6. Spearman correlation matrix of MRI dataset

### Supplementary Data 3. Classification performance and feature reduction

Supplementary Table S22. Random forest classification performance.

Performance metrics based on classification using test sample across all 15 datasets using all features and boruta importance features. 95% Confidence interval (CI) is measured across 10 train test splits.

| Modality | Method | Sample size | AUC [95% CI] | Accuracy [95% CI] | Sensitivity [95% CI] | Specificity [95% CI] | Feature Number [95% CI] |
|---|---|---|---|---|---|---|---|
| Q + C + M | boruta | 507 | 0.699 [0.669 - 0.729] | 0.709 [0.679 - 0.739] | 0.622 [0.568 - 0.676] | 0.778 [0.74 - 0.816] | 23.8 [22.988 - 24.612] |
| C + M | boruta | 622 | 0.679 [0.633 - 0.725] | 0.683 [0.638 - 0.728] | 0.593 [0.532 - 0.654] | 0.762 [0.721 - 0.803] | 24.9 [23.92 - 25.88] |
| Q + C + S + M | all | 492 | 0.676 [0.628 - 0.724] | 0.695 [0.65 - 0.74] | 0.508 [0.417 - 0.599] | 0.84 [0.799 - 0.881] | 144 [144 - 144] |
| Q + M | boruta | 545 | 0.674 [0.623 - 0.725] | 0.689 [0.638 - 0.74] | 0.585 [0.51 - 0.66] | 0.766 [0.699 - 0.833] | 15.8 [14.988 - 16.612] |
| Q + C | boruta | 819 | 0.674 [0.639 - 0.709] | 0.684 [0.65 - 0.718] | 0.573 [0.511 - 0.635] | 0.778 [0.743 - 0.813] | 19 [18.416 - 19.584] |
| Q + C + M | all | 507 | 0.672 [0.634 - 0.71] | 0.686 [0.65 - 0.722] | 0.555 [0.485 - 0.625] | 0.791 [0.748 - 0.834] | 96 [96 - 96] |
| Q + M | all | 545 | 0.671 [0.623 - 0.719] | 0.688 [0.638 - 0.738] | 0.54 [0.486 - 0.594] | 0.799 [0.736 - 0.862] | 53 [53 - 53] |
| Q + C + S | all | 776 | 0.671 [0.638 - 0.704] | 0.684 [0.651 - 0.717] | 0.531 [0.494 - 0.568] | 0.809 [0.765 - 0.853] | 118 [118 - 118] |
| C + M | all | 622 | 0.67 [0.62 - 0.72] | 0.678 [0.63 - 0.726] | 0.568 [0.502 - 0.634] | 0.769 [0.717 - 0.821] | 73 [73 - 73] |
| Q + C + S | boruta | 776 | 0.67 [0.633 - 0.707] | 0.68 [0.64 - 0.72] | 0.569 [0.526 - 0.612] | 0.769 [0.72 - 0.818] | 22.8 [21.547 - 24.053] |
| Q + C + S + M | boruta | 492 | 0.667 [0.629 - 0.705] | 0.675 [0.639 - 0.711] | 0.585 [0.521 - 0.649] | 0.749 [0.698 - 0.8] | 24.3 [22.992 - 25.608] |
| C + S + M | all | 607 | 0.665 [0.624 - 0.706] | 0.677 [0.637 - 0.717] | 0.552 [0.49 - 0.614] | 0.783 [0.739 - 0.827] | 121 [121 - 121] |
| Q + C | all | 819 | 0.658 [0.637 - 0.679] | 0.669 [0.648 - 0.69] | 0.55 [0.503 - 0.597] | 0.767 [0.716 - 0.818] | 70 [70 - 70] |
| S + M | boruta | 666 | 0.652 [0.628 - 0.676] | 0.669 [0.645 - 0.693] | 0.522 [0.471 - 0.573] | 0.782 [0.743 - 0.821] | 20.1 [19.01 - 21.19] |
| C + S + M | boruta | 607 | 0.65 [0.605 - 0.695] | 0.66 [0.617 - 0.703] | 0.552 [0.472 - 0.632] | 0.752 [0.697 - 0.807] | 23.8 [21.304 - 26.296] |
| Q + S + M | all | 528 | 0.648 [0.586 - 0.71] | 0.674 [0.615 - 0.733] | 0.474 [0.367 - 0.581] | 0.824 [0.767 - 0.881] | 101 [101 - 101] |
| Q + S + M | boruta | 528 | 0.646 [0.585 - 0.707] | 0.665 [0.607 - 0.723] | 0.512 [0.431 - 0.593] | 0.782 [0.72 - 0.844] | 17.1 [14.877 - 19.323] |

| | | | | | | | |
|---|---|---|---|---|---|---|---|
| S + M | all | 666 | 0.646 [0.619 - 0.673] | 0.668 [0.641 - 0.695] | 0.473 [0.407 - 0.539] | 0.819 [0.766 - 0.872] | 78 [78 - 78] |
| M | boruta | 683 | 0.645 [0.618 - 0.672] | 0.657 [0.636 - 0.678] | 0.549 [0.468 - 0.63] | 0.74 [0.701 - 0.779] | 15.9 [14.863 - 16.937] |
| Q + S | all | 823 | 0.645 [0.604 - 0.686] | 0.662 [0.621 - 0.703] | 0.493 [0.431 - 0.555] | 0.798 [0.748 - 0.848] | 75 [75 - 75] |
| C + S | all | 920 | 0.644 [0.615 - 0.673] | 0.658 [0.628 - 0.688] | 0.493 [0.446 - 0.54] | 0.796 [0.76 - 0.832] | 95 [95 - 95] |
| M | all | 683 | 0.637 [0.599 - 0.675] | 0.65 [0.615 - 0.685] | 0.533 [0.438 - 0.628] | 0.741 [0.679 - 0.803] | 30 [30 - 30] |
| Q | boruta | 870 | 0.631 [0.61 - 0.652] | 0.641 [0.617 - 0.665] | 0.529 [0.481 - 0.577] | 0.732 [0.69 - 0.774] | 7.8 [7.061 - 8.539] |
| C + S | boruta | 920 | 0.623 [0.594 - 0.652] | 0.634 [0.607 - 0.661] | 0.504 [0.45 - 0.558] | 0.742 [0.712 - 0.772] | 19 [17.348 - 20.652] |
| Q | all | 870 | 0.623 [0.596 - 0.65] | 0.638 [0.612 - 0.664] | 0.489 [0.434 - 0.544] | 0.758 [0.708 - 0.808] | 27 [27 - 27] |
| C | all | 964 | 0.618 [0.586 - 0.65] | 0.628 [0.596 - 0.66] | 0.498 [0.439 - 0.557] | 0.737 [0.673 - 0.801] | 47 [47 - 47] |
| Q + S | boruta | 823 | 0.617 [0.577 - 0.657] | 0.631 [0.592 - 0.67] | 0.488 [0.423 - 0.553] | 0.747 [0.697 - 0.797] | 14.7 [13.871 - 15.529] |
| C | boruta | 964 | 0.61 [0.577 - 0.643] | 0.62 [0.588 - 0.652] | 0.499 [0.437 - 0.561] | 0.722 [0.675 - 0.769] | 15.4 [13.674 - 17.126] |
| S | all | 997 | 0.581 [0.549 - 0.613] | 0.6 [0.569 - 0.631] | 0.402 [0.345 - 0.459] | 0.76 [0.726 - 0.794] | 52 [52 - 52] |
| S | boruta | 997 | 0.569 [0.538 - 0.6] | 0.587 [0.557 - 0.617] | 0.417 [0.373 - 0.461] | 0.723 [0.69 - 0.756] | 11.5 [9.909 - 13.091] |

CI - Confidence interval, Q - Questionnaire, C - Clincal physical assessment, M - Spino-pelvic MRI, S - Back shape and function

#### Supplementary Table S23. Random forest classification performance (Imputaion).

| Modality | Method | Sample Size | AUC [95% CI] | Accuracy [95% CI] | Sensitivity [95% CI] | Specificity [95% CI] | Feature Number [95% CI] |
|---|---|---|---|---|---|---|---|
| Q + C + M | boruta | 1045 | 0.673 [0.654 - 0.692] | 0.685 [0.664 - 0.706] | 0.556 [0.526 - 0.586] | 0.788 [0.742 - 0.834] | 24.6 [23.471 - 25.729] |

| | | | | | | | |
|---|---|---|---|---|---|---|---|
| Q + C | boruta | 1045 | 0.668 [0.635 - 0.701] | 0.682 [0.65 - 0.714] | 0.537 [0.47 - 0.604] | 0.796 [0.748 - 0.844] | 20.7 [19.228 - 22.172] |
| Q + C + S + M | all | 1045 | 0.662 [0.625 - 0.699] | 0.682 [0.645 - 0.719] | 0.513 [0.46 - 0.566] | 0.817 [0.77 - 0.864] | 154 [154 - 154] |
| Q + C + S + M | boruta | 1045 | 0.661 [0.627 - 0.695] | 0.677 [0.643 - 0.711] | 0.531 [0.483 - 0.579] | 0.791 [0.745 - 0.837] | 28 [26.455 - 29.545] |
| Q + C + M | all | 1045 | 0.659 [0.629 - 0.689] | 0.675 [0.645 - 0.705] | 0.524 [0.464 - 0.584] | 0.793 [0.741 - 0.845] | 106 [106 - 106] |
| Q + C | all | 1045 | 0.658 [0.629 - 0.687] | 0.672 [0.64 - 0.704] | 0.526 [0.475 - 0.577] | 0.786 [0.73 - 0.842] | 74 [74 - 74] |
| Q + C + S | all | 1045 | 0.656 [0.624 - 0.688] | 0.675 [0.64 - 0.71] | 0.488 [0.442 - 0.534] | 0.824 [0.762 - 0.886] | 122 [122 - 122] |
| Q + C + S | boruta | 1045 | 0.654 [0.623 - 0.685] | 0.668 [0.637 - 0.699] | 0.534 [0.479 - 0.589] | 0.777 [0.725 - 0.829] | 25 [23.216 - 26.784] |
| Q + S + M | all | 1045 | 0.648 [0.617 - 0.679] | 0.666 [0.636 - 0.696] | 0.484 [0.446 - 0.522] | 0.812 [0.779 - 0.845] | 108 [108 - 108] |
| C + S + M | all | 1045 | 0.643 [0.606 - 0.68] | 0.662 [0.624 - 0.7] | 0.478 [0.429 - 0.527] | 0.808 [0.759 - 0.857] | 130 [130 - 130] |
| C + S + M | boruta | 1045 | 0.641 [0.602 - 0.68] | 0.655 [0.617 - 0.693] | 0.516 [0.452 - 0.58] | 0.768 [0.719 - 0.817] | 24.1 [23.063 - 25.137] |
| C + M | all | 1045 | 0.638 [0.62 - 0.656] | 0.657 [0.637 - 0.677] | 0.476 [0.44 - 0.512] | 0.8 [0.755 - 0.845] | 82 [82 - 82] |
| Q + S + M | boruta | 1045 | 0.638 [0.615 - 0.661] | 0.656 [0.634 - 0.678] | 0.484 [0.448 - 0.52] | 0.791 [0.763 - 0.819] | 21.9 [20.71 - 23.09] |
| C | boruta | 1045 | 0.637 [0.598 - 0.676] | 0.653 [0.613 - 0.693] | 0.494 [0.434 - 0.554] | 0.781 [0.724 - 0.838] | 15 [14.416 - 15.584] |
| C + S | all | 1045 | 0.635 [0.595 - 0.675] | 0.655 [0.615 - 0.695] | 0.452 [0.383 - 0.521] | 0.816 [0.761 - 0.871] | 98 [98 - 98] |
| C + S | boruta | 1045 | 0.633 [0.591 - 0.675] | 0.648 [0.606 - 0.69] | 0.5 [0.433 - 0.567] | 0.765 [0.701 - 0.829] | 19.9 [18.375 - 21.425] |
| C | all | 1045 | 0.631 [0.597 - 0.665] | 0.65 [0.616 - 0.684] | 0.469 [0.418 - 0.52] | 0.794 [0.74 - 0.848] | 50 [50 - 50] |
| Q + M | boruta | 1045 | 0.631 [0.592 - 0.67] | 0.646 [0.612 - 0.68] | 0.494 [0.417 - 0.571] | 0.77 [0.727 - 0.813] | 16.5 [15.32 - 17.68] |

| | | | | | | | |
|---|---|---|---|---|---|---|---|
| Q + S | all | 1045 | 0.631 [0.6 - 0.662] | 0.651 [0.619 - 0.683] | 0.447 [0.41 - 0.484] | 0.811 [0.767 - 0.855] | 76 [76 - 76] |
| C + M | boruta | 1045 | 0.63 [0.592 - 0.668] | 0.645 [0.609 - 0.681] | 0.498 [0.436 - 0.56] | 0.763 [0.719 - 0.807] | 17.8 [16.299 - 19.301] |
| Q | boruta | 1045 | 0.627 [0.588 - 0.666] | 0.641 [0.604 - 0.678] | 0.507 [0.459 - 0.555] | 0.747 [0.693 - 0.801] | 9.6 [8.695 - 10.505] |
| Q | all | 1045 | 0.616 [0.591 - 0.641] | 0.631 [0.607 - 0.655] | 0.482 [0.429 - 0.535] | 0.747 [0.715 - 0.779] | 28 [28 - 28] |
| Q + S | boruta | 1045 | 0.616 [0.589 - 0.643] | 0.631 [0.604 - 0.658] | 0.473 [0.426 - 0.52] | 0.756 [0.717 - 0.795] | 19.1 [17.818 - 20.382] |
| S + M | all | 1045 | 0.613 [0.57 - 0.656] | 0.636 [0.594 - 0.678] | 0.418 [0.36 - 0.476] | 0.808 [0.771 - 0.845] | 84 [84 - 84] |
| Q + M | all | 1045 | 0.612 [0.589 - 0.635] | 0.631 [0.609 - 0.653] | 0.453 [0.391 - 0.515] | 0.771 [0.724 - 0.818] | 60 [60 - 60] |
| S + M | boruta | 1045 | 0.606 [0.579 - 0.633] | 0.624 [0.598 - 0.65] | 0.44 [0.396 - 0.484] | 0.772 [0.732 - 0.812] | 16.7 [15.349 - 18.051] |
| S | boruta | 1045 | 0.596 [0.557 - 0.635] | 0.614 [0.576 - 0.652] | 0.437 [0.386 - 0.488] | 0.754 [0.715 - 0.793] | 14 [12.314 - 15.686] |
| S | all | 1045 | 0.583 [0.55 - 0.616] | 0.608 [0.575 - 0.641] | 0.368 [0.318 - 0.418] | 0.798 [0.751 - 0.845] | 52 [52 - 52] |
| M | boruta | 1045 | 0.58 [0.547 - 0.613] | 0.61 [0.576 - 0.644] | 0.346 [0.293 - 0.399] | 0.817 [0.761 - 0.873] | 18.1 [16.27 - 19.93] |
| M | all | 1045 | 0.571 [0.543 - 0.599] | 0.599 [0.569 - 0.629] | 0.314 [0.266 - 0.362] | 0.826 [0.775 - 0.877] | 36 [36 - 36] |

CI - Confidence interval, Q - Questionnaire, C - Clincal physical assessment, M - Spino-pelvic MRI, S - Back shape and function

#### Supplementary Table S24. Boruta selected important features in the questionnaire modality dataset.

| **Modality** | **Assessment** | **Selected (%)** | **Importance (mean)** |
|---|---|---|---|
| questionnaire | SF-36 social function | 100 | 21.42 |
| questionnaire | SF-36 psychological well being | 100 | 20.02 |
| questionnaire | hip pain | 100 | 15 |
| questionnaire | smoking pack years | 100 | 11.8 |
| questionnaire | family back pain | 100 | 7.27 |
| demographic | age | 80 | 6.15 |
| questionnaire | physical activity 150min/week | 70 | 6.04 |

| | | | |
|---|---|---|---|
| questionnaire | job load | 70 | 5.25 |
| questionnaire | living situation | 10 | 4.59 |
| questionnaire | BRSQ intrinsic motivation | 30 | 4.53 |
| questionnaire | BRSQ integrated regulation | 20 | 3.86 |

BRSQ - Behavioural Regulation in Sport Questionnaire, SF-36 - Short-form 36 Health Status Questionnaire.

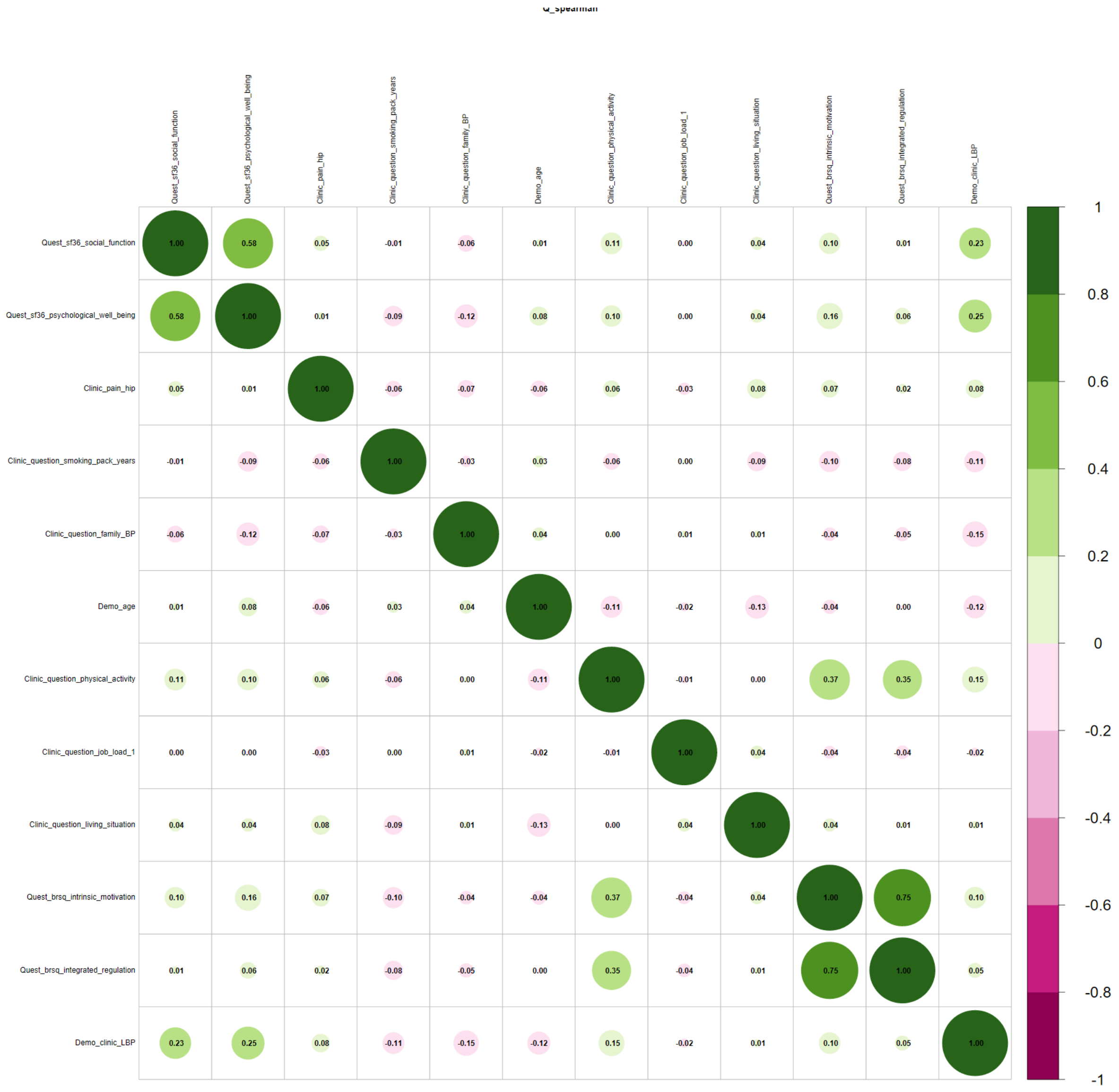

Supplementary Figure S7. Spearman correlation matrix of Boruta selected important features for questionnaire dataset

Supplementary Table S25. Boruta selected important features for clinical physical assessment dataset

| Modality | Variable | Selected (%) | Importance (mean) |
|---|---|---|---|
| Clinical | cervical axial rotate left | 100 | 14.19 |
| Clinical | sit to stand 30sec | 100 | 13.61 |
| Clinical | general mobility | 100 | 13.39 |
| Clinical | rigid muscle | 100 | 11.22 |
| Clinical | cervical axial rotate right | 100 | 7.82 |
| Clinical | hip flexion left | 100 | 6.29 |
| Demographic | age | 100 | 5.61 |
| Clinical | hip flexion right | 90 | 5.53 |
| Clinical | hip abduction left | 90 | 5.5 |
| Clinical | hip internal rotate right | 60 | 5.17 |
| Clinical | shober | 50 | 4.68 |
| Clinical | chin sternum distance | 80 | 4.58 |
| Clinical | thoracic lumbar lateral bend right | 70 | 4.42 |
| Clinical | cervical lateral bend left | 60 | 4.35 |
| Clinical | cervical inclination | 10 | 4.33 |
| Clinical | hip internal rotate left | 40 | 4.29 |
| Clinical | back form | 90 | 4.25 |
| Clinical | hip extension right | 10 | 4.16 |
| Clinical | thoracic lumbar inclination | 30 | 4.08 |
| Clinical | thomas handle right | 10 | 4.03 |
| Clinical | hip abduction right | 70 | 3.99 |
| Clinical | lumbar bulge | 10 | 3.81 |
| Demographic | sex | 10 | 3.76 |
| Clinical | roussoly type | 10 | 3.67 |
| Clinical | cervical reclination | 40 | 3.59 |
| Clinical | asymmetrical waist triangle | 10 | 3.42 |

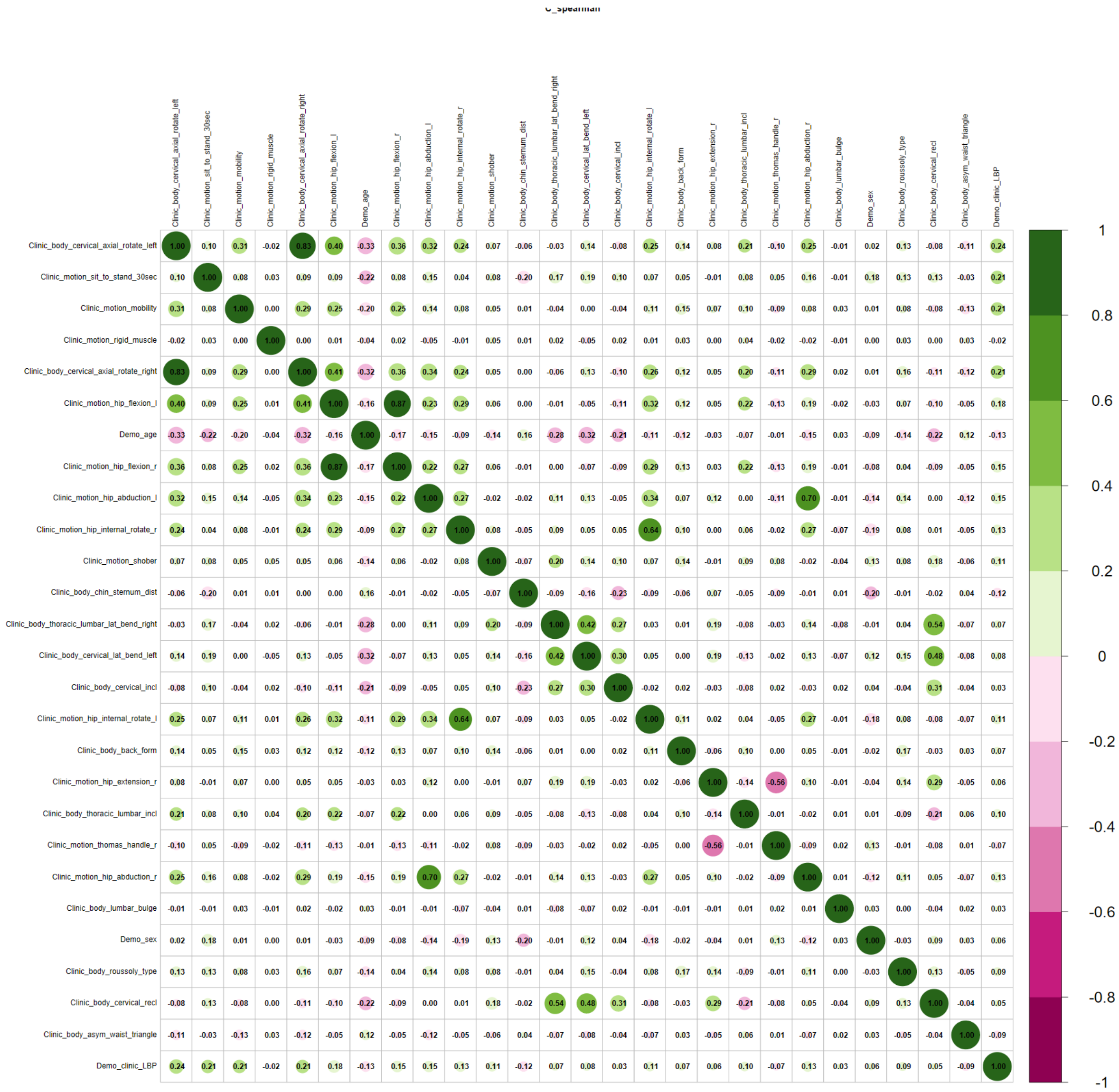

Supplementary Figure S8. Spearman correlation matrix of Boruta selected important features for clinical assessment dataset

Supplementary Table S26. Boruta selected important features for superficial spine morphology modality dataset

| **Modality** | **Variable** | **selected (%)** | **Importance (mean)** |
|---|---|---|---|
| Demographic | age | 100 | 7.8 |
| Back shape & function | sacral frontal stand left | 100 | 6.51 |
| Back shape & function | inclination value frontal stand left | 100 | 6.26 |
| Back shape & function | inclination value sagittal sit extension | 80 | 5.22 |
| Back shape & function | thoracic value sagittal stand extension | 90 | 4.96 |

| | | | |
|---|---|---|---|
| Back shape & function | thoracic value frontal stand left | 70 | 4.86 |
| Back shape & function | lumbar value frontal stand right | 100 | 4.81 |
| Back shape & function | sacral frontal sit left | 70 | 4.78 |
| Back shape & function | inclination value sagittal stand extension | 30 | 4.38 |
| Back shape & function | lumbar value sagittal sit flexion | 80 | 4.19 |
| Back shape & function | spine length sagittal stand extension | 70 | 4.15 |
| Back shape & function | inclination value frontal stand right | 70 | 3.92 |
| Back shape & function | thoracic value frontal sit left | 70 | 3.92 |
| Back shape & function | inclination value frontal sit left | 60 | 3.9 |
| Back shape & function | sacral frontal stand straight | 10 | 3.66 |
| Back shape & function | lumbar value sagittal sit extension | 40 | 3.48 |
| Back shape & function | lumbar value sagittal stand extension | 10 | 3.46 |

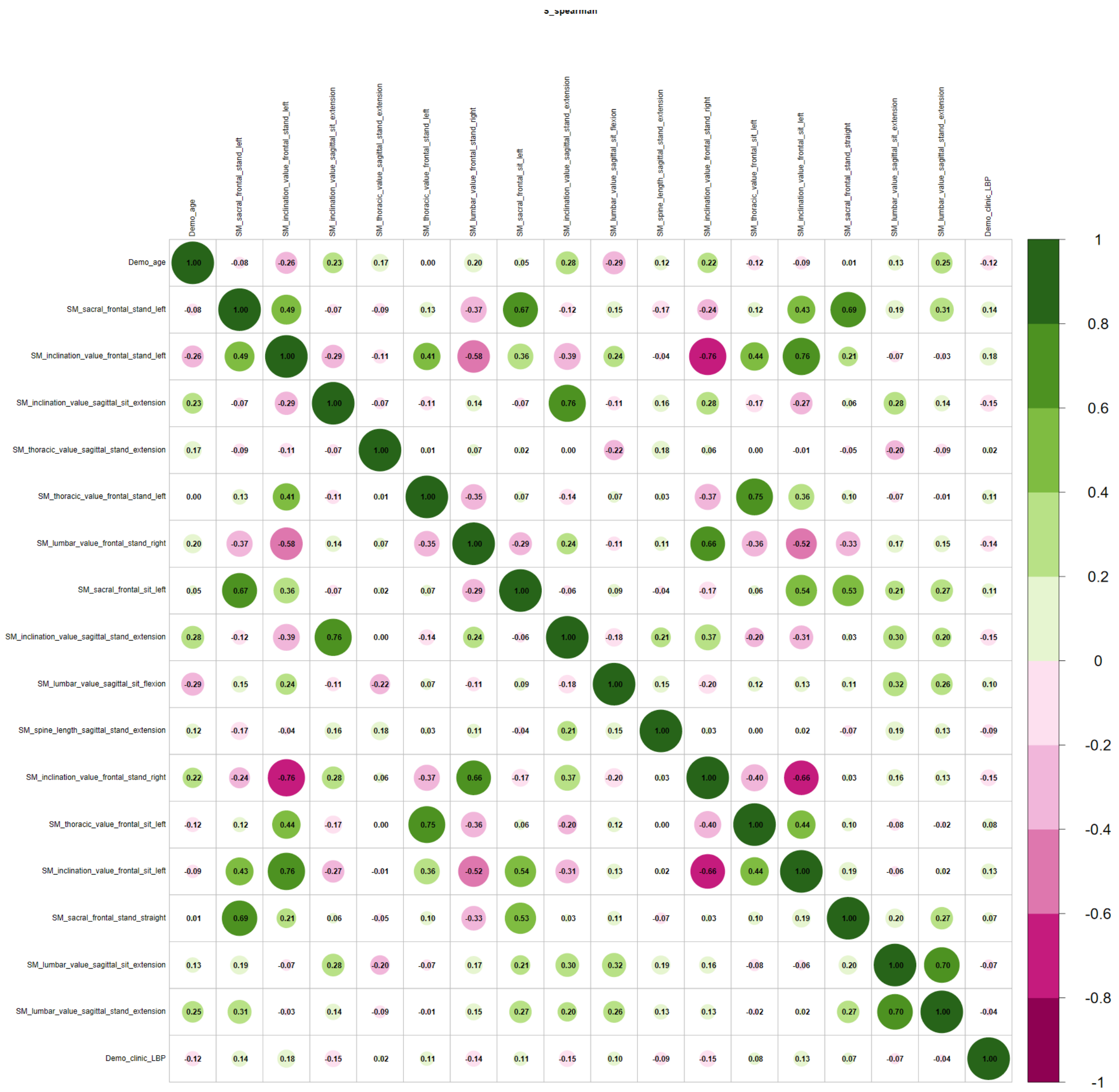

Supplementary Figure S9. Spearman correlation matrix of Boruta selected important features for back shape and function dataset

Supplementary Table S27. Boruta selected important features for MRI modality dataset

| Modality | Assessment | Selected (%) | Importance (mean) |
|---|---|---|---|
| MRI | IVD herniation L4 - L5 | 100 | 19.49 |
| MRI | spinal canal width L2 | 100 | 9.94 |
| MRI | IVD degeneration L3 - L4 | 100 | 9.46 |
| MRI | IVD degeneration L4 - L5 | 100 | 9.14 |
| MRI | spinal canal width L1 | 100 | 8.13 |
| MRI | osteochondrosis L4 - L5 | 100 | 8.12 |
| MRI | IVD herniation L5 - S1 | 100 | 7.38 |
| MRI | IVD degeneration L2 - L3 | 100 | 6.85 |
| MRI | facet joint L3 - L4 right | 100 | 6.81 |
| MRI | facet joint L5/S1 - L5/L6 right | 100 | 6.56 |
| MRI | facet joint L3 - L4 left | 90 | 5.53 |
| Demographic | age | 30 | 5.29 |
| MRI | facet joint L1 - L2 right | 30 | 5.12 |
| MRI | IVD degeneration L5 - S1 | 80 | 4.97 |
| MRI | osteochondrosis L5 - S1 | 60 | 4.66 |
| MRI | IVD degeneration L1 - L2 | 100 | 4.64 |
| MRI | facet joint L1 - L2 left | 20 | 4.59 |
| MRI | facet joint L5/S1 - L5/L6 left | 80 | 4.34 |
| MRI | IVD herniation L2 - L3 | 60 | 4.16 |
| MRI | IVD herniation L3 - L4 | 30 | 4.08 |
| MRI | spinal canal width L4 | 10 | 3.77 |

Boruta selected important variables in MRI modality dataset across ten-fold feature selection.
IVD – intervertebral disc

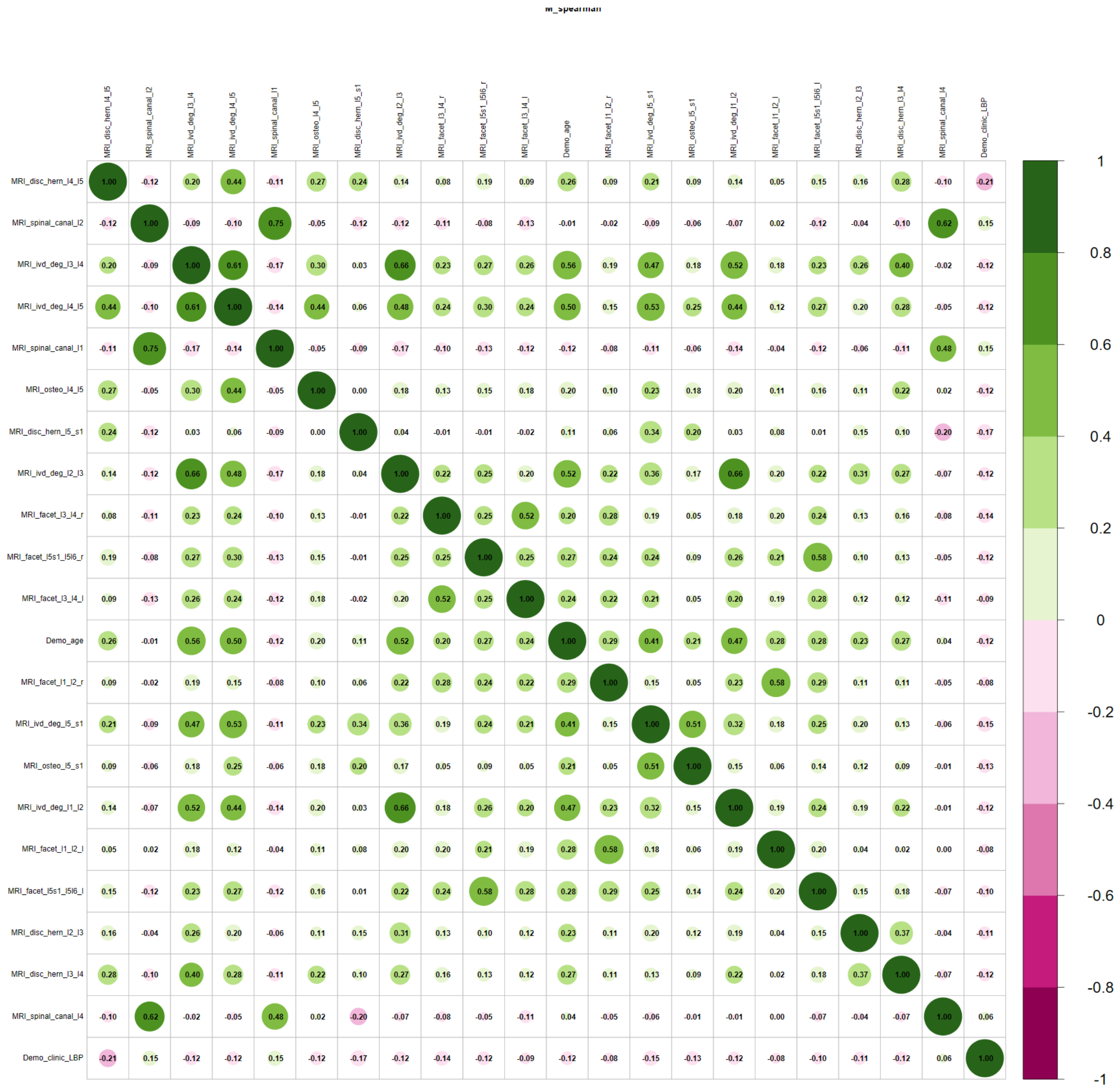

Supplementary Figure S10. Spearman correlation matrix of Boruta selected important features for MRI dataset

Supplementary Table S28. Boruta selected important features for questionnaire and clinical physical assessment dataset

| Modality | Variable | selected (%) | Importance (mean) |
|---|---|---|---|
| Questionnaire | SF-36 psychological well being | 100 | 17.04 |
| Questionnaire | SF-36 social function | 100 | 15.02 |
| Clinical | Hip pain | 100 | 12.39 |
| Clinical | general mobility | 100 | 11.39 |
| Clinical | cervical axial rotate left | 100 | 10.62 |

| | | | |
|---|---|---|---|
| Clinical | sit to stand 30sec | 100 | 9.78 |
| Questionnaire | smoking pack years | 100 | 9.62 |
| Clinical | rigid muscle | 100 | 8.52 |
| Questionnaire | family back pain | 100 | 6.53 |
| Clinical | hip abduction left | 100 | 6 |
| Questionnaire | physical activity 150min/week | 100 | 5.88 |
| Demographic | age | 100 | 5.45 |
| Clinical | cervical axial rotate right | 100 | 5.42 |
| Clinical | hip abduction right | 90 | 5.23 |
| Clinical | hip internal rotate right | 80 | 5.02 |
| Clinical | thoracic lumbar lat bend right | 60 | 4.72 |
| Clinical | back form | 100 | 4.34 |
| Clinical | hip flexion right | 100 | 4.27 |
| Clinical | shober | 10 | 4.2 |
| Clinical | square shoulders | 10 | 4.05 |
| Clinical | cervical lat bend left | 30 | 4.04 |
| Clinical | hip flexion left | 70 | 3.76 |
| Questionnaire | BRSQ intrinsic motivation | 20 | 3.63 |
| Clinical | finger floor distance | 10 | 3.59 |
| Questionnaire | job load 1 | 10 | 3.48 |
| Clinical | hip adduction right | 10 | 3.34 |

BRSQ - Behavioural Regulation in Sport Questionnaire, SF-36 - Short-form 36 Health Status Questionnaire.

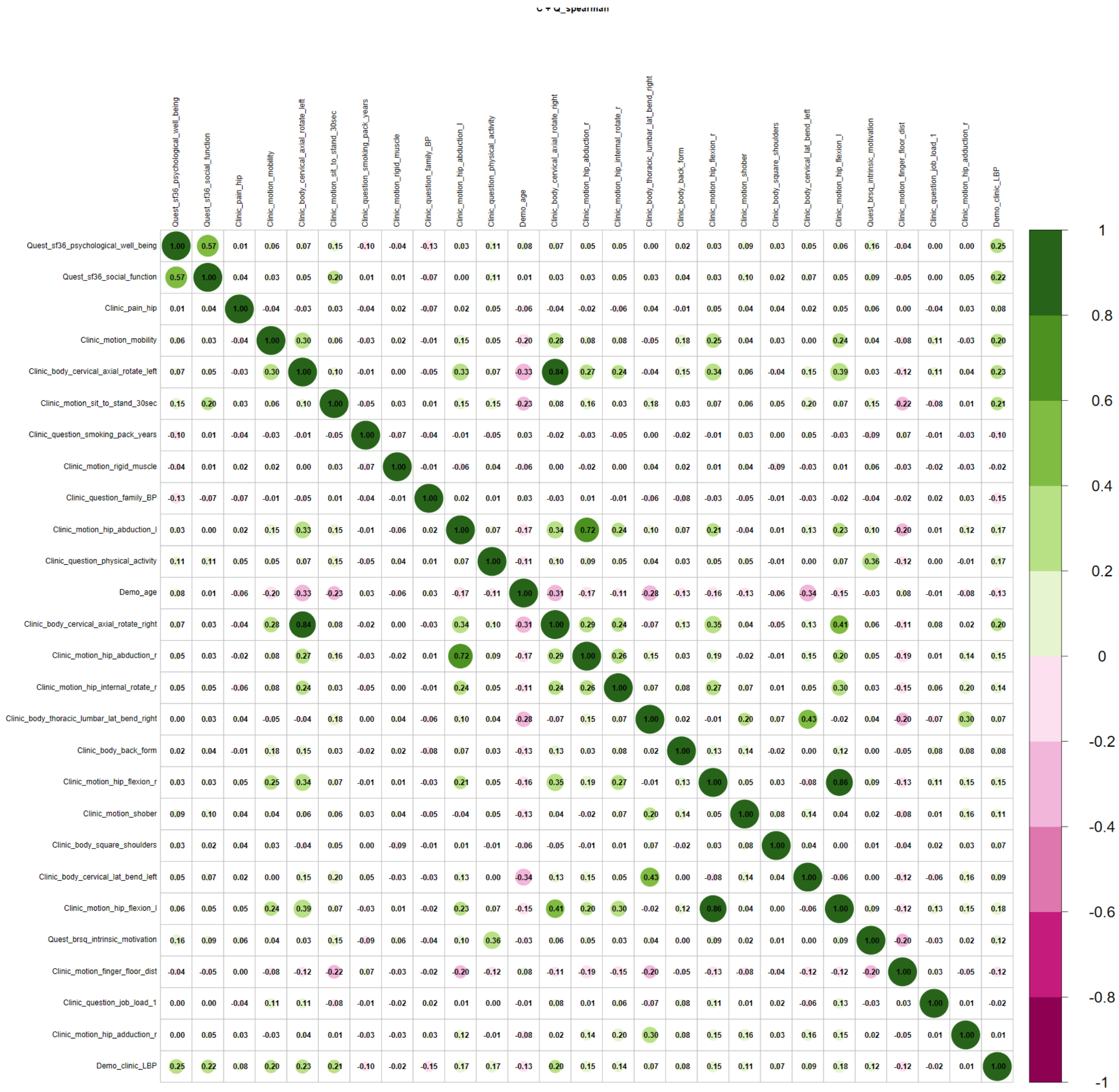

Supplementary Figure S11. Spearman correlation matrix of Boruta selected important features for questionnaire + clinic dataset

Supplementary Table S29. Boruta selected important features for clinic physical assessment and back shape and function dataset

| Modality | Variable | selected (%) | Importance (mean) |
|---|---|---|---|
| Clinical | cervical axial rotate left | 100 | 14.2 |
| Clinical | general mobility | 100 | 11.83 |
| Clinical | sit to stand 30sec | 100 | 11.53 |
| Clinical | rigid muscle | 100 | 9.97 |
| Clinical | cervical axial rotate right | 100 | 7 |
| Clinical | hip flexion left | 100 | 5.99 |
| Demogrpahic | age | 100 | 5.82 |

| | | | |
|---|---|---|---|
| Back shape & function | inclination value frontal stand left | 90 | 5.31 |
| Back shape & function | sacral frontal stand left | 90 | 5.12 |
| Back shape & function | thoracic value sagittal stand extension | 90 | 5.04 |
| Clinical | hip abduction left | 90 | 4.7 |
| Clinical | hip flexion right | 100 | 4.68 |
| Clinical | cervical lateral bend left | 20 | 4.54 |
| Clinical | back form | 70 | 4.49 |
| Back shape & function | lumbar value frontal sit straight | 10 | 4.26 |
| Back shape & function | inclination value sagittal stand extension | 10 | 4.23 |
| Back shape & function | sacral sagittal sit straight | 10 | 4.11 |
| Back shape & function | lumbar value frontal sit right | 20 | 4.05 |
| Back shape & function | lumbar value sagittal stand extension | 10 | 3.97 |
| Back shape & function | inclination value frontal stand right | 60 | 3.95 |
| Back shape & function | inclination value frontal sit left | 40 | 3.91 |
| Back shape & function | lumbar value frontal stand right | 70 | 3.81 |
| Clinical | thoracic lumbar inclination | 40 | 3.8 |
| Back shape & function | lumbar value frontal stand straight | 30 | 3.79 |
| Clinical | shober | 20 | 3.79 |
| Back shape & function | inclination value sagittal sit extension | 30 | 3.78 |
| Clinical | hip abduction right | 30 | 3.74 |
| Clinical | thoracic lumbar lateral bend right | 60 | 3.72 |
| Back shape & function | sacral frontal sit left | 50 | 3.71 |
| Back shape & function | sacral frontal sit straight | 50 | 3.59 |
| Back shape & function | thoracic value sagittal sit extension | 20 | 3.58 |
| Back shape & function | thoracic value frontal stand right | 10 | 3.46 |
| Back shape & function | thoracic value frontal stand left | 40 | 3.44 |
| Back shape & function | sacral sagittal stand straight | 20 | 3.29 |
| Clinical | chin sternum distance | 10 | 3.29 |
| Back shape & function | inclination value frontal sit right | 10 | 3.21 |

284

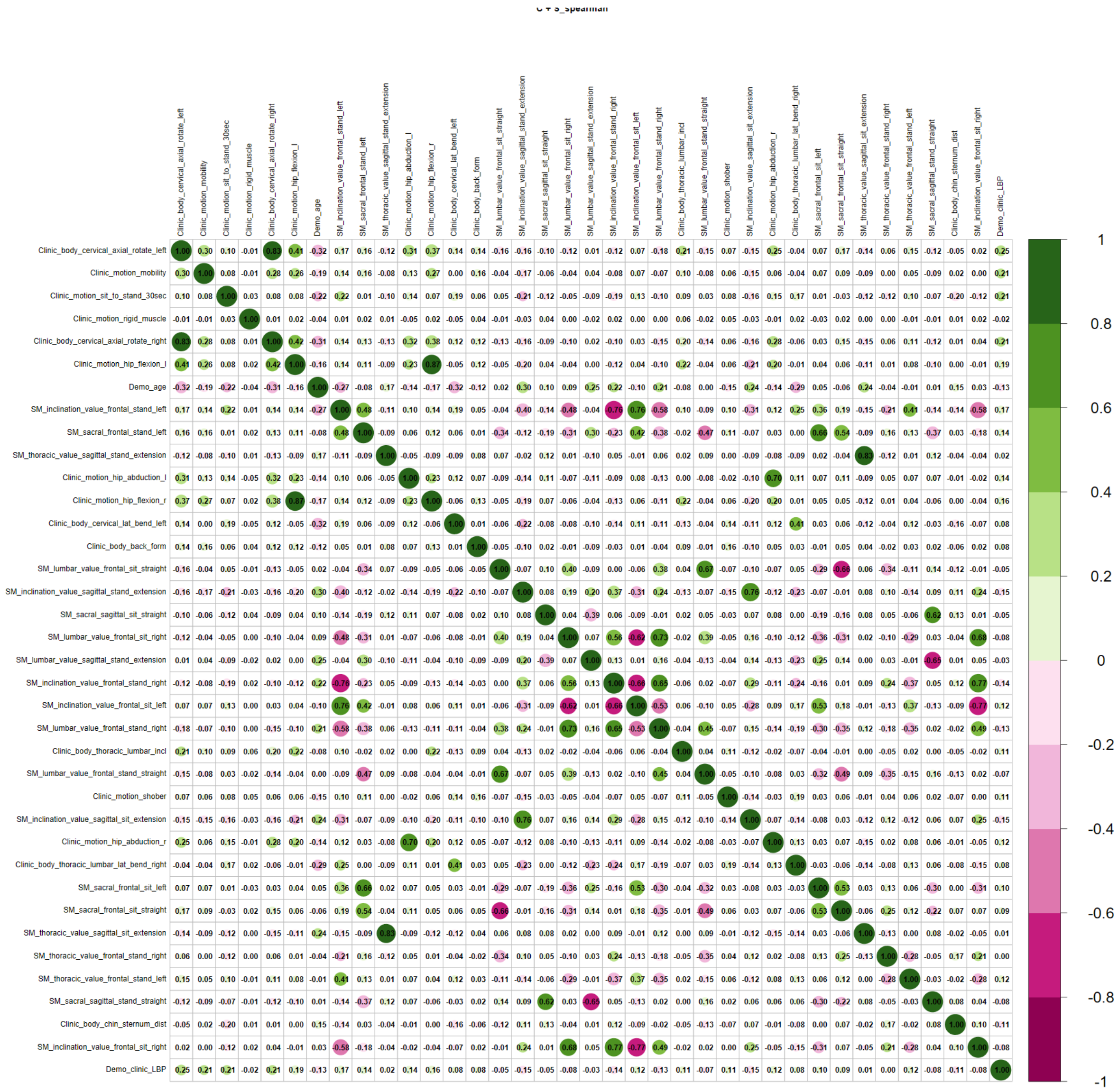

Supplementary Figure S12. Spearman correlation matrix of Boruta selected important features for clinic physical assessment and back shape and function dataset

Supplementary Table S30. Boruta selected important features for MRI and questionnaire dataset

| Modality | Variable | selected (%) | Importance (mean) |
|---|---|---|---|
| Questionnaire | SF-36 social function | 100 | 18.18 |
| Questionnaire | SF-36 psychological well being | 100 | 16.93 |
| MRI | intervertebral disc herniation L4 - L5 | 100 | 14.05 |
| MRI | spinal canal stenosis L2 | 100 | 11.4 |
| MRI | intervertebral disc degeneration L2 - L3 | 100 | 9.11 |
| Questionnaire | hip pain | 100 | 8.53 |

| | | | |
|---|---|---|---|
| MRI | spinal canal stenosis L1 | 100 | 8.05 |
| MRI | intervertebral disc degeneration L4 - L5 | 100 | 7.75 |
| MRI | intervertebral disc degeneration L5 - S1 | 100 | 6.86 |
| MRI | osteochondrosis intervertebralis L4 - L5 | 20 | 6.16 |
| MRI | intervertebral disc herniation L5 - S1 | 100 | 5.98 |
| MRI | intervertebral disc degeneration L3 - L4 | 90 | 5.76 |
| MRI | facet joint L3 - L4 right | 100 | 5.37 |
| MRI | facet joint L5/S1 - L5/L6 right | 90 | 4.8 |
| MRI | osteochondrosis intervertebralis L5 - S1 | 30 | 4.5 |
| MRI | spinal canal stenosis L4 | 60 | 4.32 |
| MRI | facet joint L4 - L5 right | 20 | 4.16 |
| MRI | intervertebral disc degeneration L1 - L2 | 30 | 4.08 |
| Questionnaire | physical activity 150min/week | 20 | 4.08 |
| MRI | facet joint L3 - L4 left | 50 | 4.06 |
| Questionnaire | living situation | 40 | 3.98 |
| MRI | spinal canal stenosis L3 | 20 | 3.97 |
| Questionnaire | SF-36 emotional role function | 10 | 3.68 |

SF-36 - Short-form 36 Health Status Questionnaire

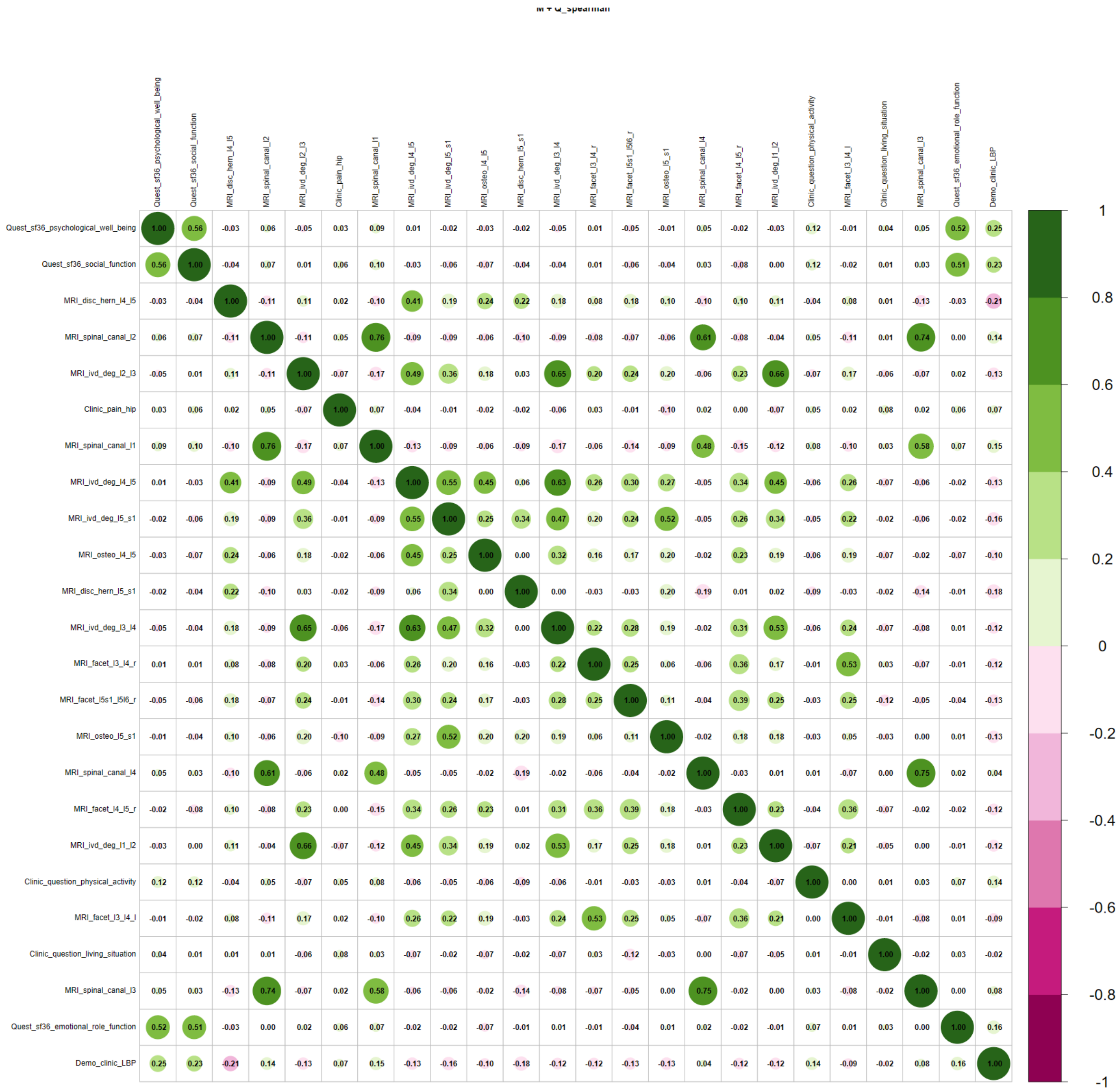

Supplementary Figure S13. Spearman correlation matrix of Boruta selected important features for questionnaire + MRI dataset

Supplementary Table S31. Boruta selected important features for MRI and clinical assessment modality dataset

| Modality | Assessment | Selected (%) | Importance (mean) |
|---|---|---|---|
| MRI | IVD herniation L4 - L5 | 100 | 13.72 |
| clinical examination | cervical axial rotate left | 100 | 13.68 |
| clinical examination | hip flexion left | 100 | 11.01 |
| clinical examination | general mobility | 100 | 9.92 |
| clinical examination | sit to stand 30sec | 100 | 9.69 |

| | | | |
|---|---|---|---|
| clinical examination | hip flexion right | 100 | 9 |
| MRI | spinal canal width L2 | 100 | 8.41 |
| MRI | IVD degeneration L2 - L3 | 100 | 7.01 |
| clinical examination | cervical axial rotate right | 100 | 6.41 |
| MRI | IVD degeneration L4 - L5 | 100 | 6.24 |
| MRI | IVD degeneration L3 - L4 | 100 | 6.03 |
| MRI | osteochondrosis L4 - L5 | 100 | 5.96 |
| clinical examination | rigid muscle | 100 | 5.66 |
| MRI | IVD herniation L5 - S1 | 100 | 5.49 |
| MRI | IVD degeneration L5 - S1 | 100 | 5.25 |
| MRI | facet joint L5/S1 - L5/L6 right | 90 | 5.05 |
| clinical examination | chin sternum distance | 90 | 4.99 |
| clinical examination | hip internal rotate right | 100 | 4.95 |
| MRI | osteochondrosis L5 - S1 | 90 | 4.85 |
| MRI | spinal canal width L1 | 100 | 4.72 |
| clinical examination | hip internal rotate left | 90 | 4.68 |
| MRI | IVD degeneration L1 - L2 | 90 | 4.67 |
| MRI | facet joint L5/S1 - L5/L6 left | 70 | 4.47 |
| clinical examination | hip abduction right | 20 | 4.47 |
| clinical examination | hip abduction left | 30 | 4.3 |
| MRI | facet joint L3 L4 right | 60 | 4.11 |
| Demographic | age | 40 | 4.09 |
| clinical examination | thoracic lumbar inclination | 30 | 4.08 |
| clinical examination | thoracic lumbar axial rotate left | 10 | 3.87 |
| clinical examination | asymmetrical waist triangle | 40 | 3.72 |
| MRI | IVD herniation L3 - L4 | 10 | 3.51 |
| MRI | facet joint L3 - L4 left | 20 | 3.48 |
| clinical examination | back form | 10 | 3.42 |

IVD – intervertebral disc

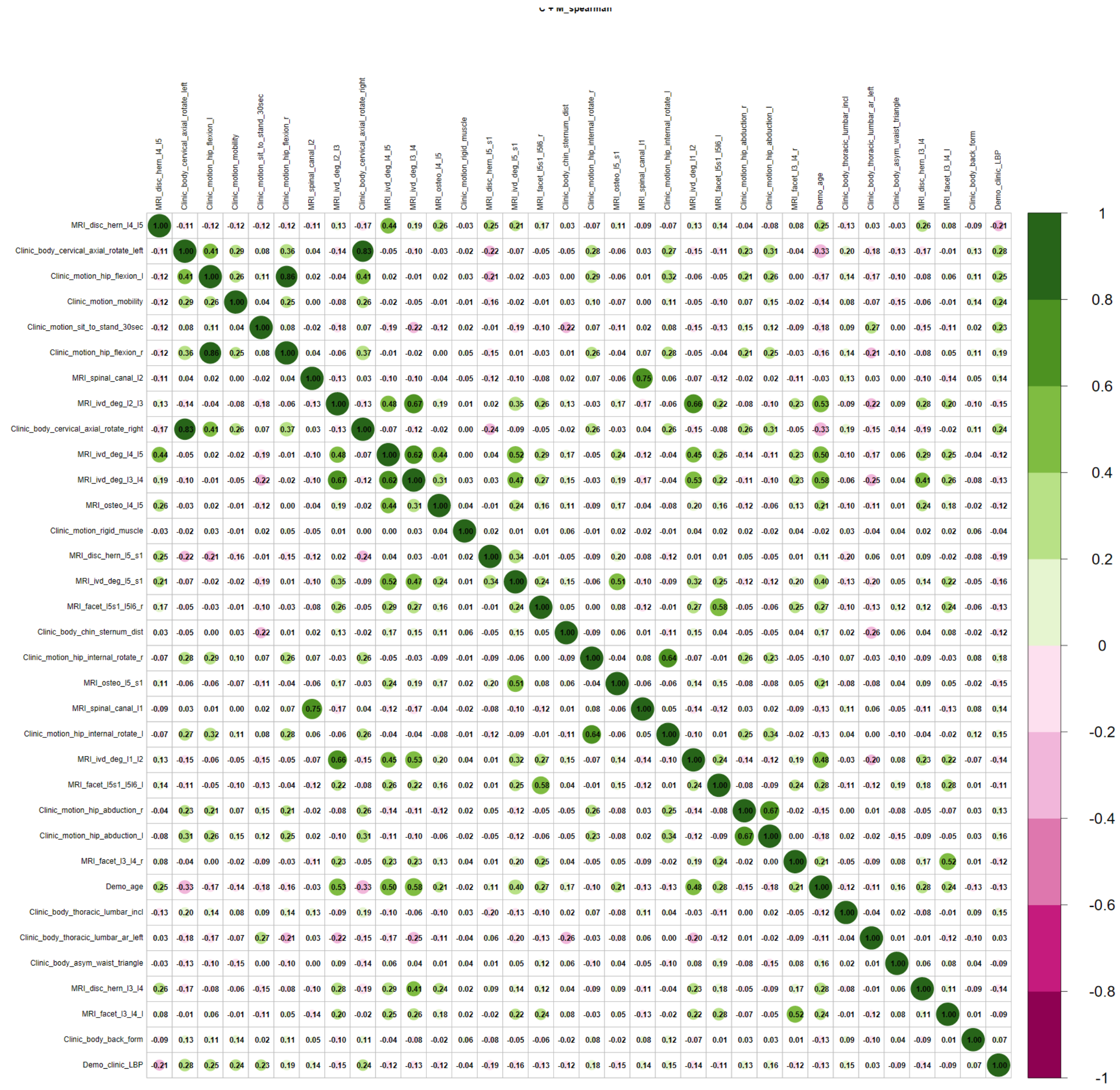

Supplementary Figure S14. Spearman correlation matrix of Boruta selected important features for clinical assessment + MRI dataset

Supplementary Table S32. Boruta selected important features for MRI and superficial spine morphology dataset

| Modality | Variable | selected (%) | Importance (mean) |
|---|---|---|---|
| MRI | intervertebral disc herniation L4 - L5 | 100 | 16.13 |
| MRI | spinal canal stenosis L2 | 100 | 9.18 |
| MRI | intervertebral disc degeneration L4 - L5 | 100 | 8.14 |
| MRI | intervertebral disc degeneration L3 - L4 | 100 | 7.25 |
| MRI | osteochondrosis intervertebralis L4 - L5 | 100 | 7.09 |

| | | | |
|---|---|---|---|
| MRI | spinal canal stenosis L1 | 100 | 6.45 |
| MRI | intervertebral disc degeneration L2 - L3 | 100 | 6.08 |
| Back shape & function | spine length sagittal stand extension | 100 | 5.42 |
| MRI | intervertebral disc herniation L5 - S1 | 100 | 5.25 |
| Back shape & function | inclination value sagittal sit extension | 90 | 5.03 |
| Back shape & function | inclination value sagittal stand extension | 80 | 4.78 |
| MRI | intervertebral disc degeneration L5 - S1 | 80 | 4.76 |
| Back shape & function | inclination value frontal stand right | 90 | 4.65 |
| MRI | facet joint L3 - L4 right | 80 | 4.54 |
| Back shape & function | inclination value sagittal sit flexion | 10 | 4.47 |
| Back shape & function | lumbar value frontal stand left | 50 | 4.43 |
| Demographic | age | 70 | 4.39 |
| Back shape & function | sacral frontal stand left | 40 | 4.39 |
| Back shape & function | inclination value frontal stand left | 50 | 4.37 |
| Back shape & function | thoracic value sagittal stand extension | 40 | 4.3 |
| Back shape & function | lumbar value sagittal sit extension | 60 | 4.29 |
| Back shape & function | lumbar value frontal stand right | 60 | 4.23 |
| MRI | osteochondrosis intervertebralis L5 - S1 | 70 | 4.18 |
| Back shape & function | lumbar value frontal stand straight | 40 | 4.13 |
| MRI | intervertebral disc degeneration L1 - L2 | 20 | 4.06 |
| Back shape & function | thoracic value frontal sit left | 40 | 3.95 |
| MRI | intervertebral disc herniation L2 - L3 | 60 | 3.85 |
| MRI | facet joint L5/S1 - L5/L6 right | 30 | 3.81 |
| Back shape & function | lumbar value sagittal sit flexion | 30 | 3.62 |
| Back shape & function | sacral frontal sit left | 10 | 3.36 |
| MRI | facet joint L3 - L4 left | 10 | 3.32 |

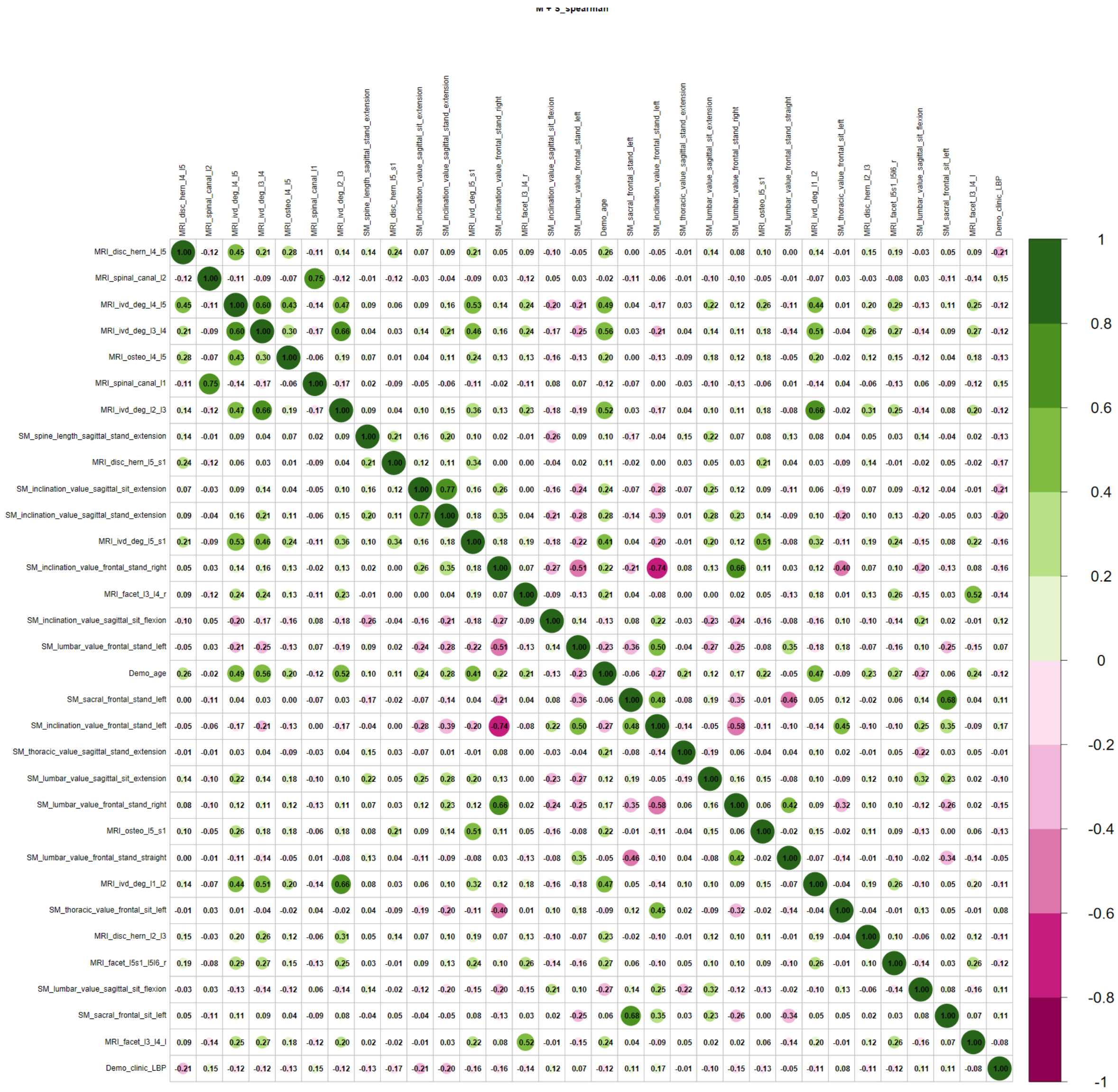

Supplementary Figure S15. Spearman correlation matrix of Boruta selected important features for back shape and function + MRI dataset

Supplementary Table S33. Boruta selected important features for questionnaire and superficial spine morphology dataset

| Modality | Variable | selected (%) | Importance (mean) |
|---|---|---|---|
| Questionnaire | SF36 social function | 100 | 19.85 |
| Questionnaire | SF36 psychological well being | 100 | 19.21 |
| Questionnaire | hip pain | 100 | 10.33 |
| Questionnaire | smoking pack years | 100 | 8.62 |
| Back shape & function | inclination value frontal stand left | 100 | 6.52 |

| | | | |
|---|---|---|---|
| Demographic | age | 80 | 5.34 |
| Back shape & function | inclination value frontal sit left | 90 | 5.2 |
| Questionnaire | family back pain | 100 | 5.17 |
| Back shape & function | inclination value sagittal stand flexion | 10 | 5.09 |
| Questionnaire | SF36 emotional role function | 20 | 4.97 |
| Questionnaire | physical activity 150min/week | 90 | 4.95 |
| Back shape & function | thoracic value frontal stand left | 50 | 4.93 |
| Back shape & function | inclination value frontal sit right | 10 | 4.83 |
| Back shape & function | sacral frontal stand left | 60 | 4.63 |
| Back shape & function | thoracic value frontal sit left | 80 | 4.61 |
| Back shape & function | lumbar value sagittal sit flexion | 60 | 4.58 |
| Back shape & function | inclination value frontal stand right | 20 | 4.45 |
| Back shape & function | inclination value sagittal stand extension | 60 | 4.24 |
| Questionnaire | family conflicts | 20 | 4.24 |
| Back shape & function | lumbar value sagittal stand extension | 10 | 4.01 |
| Back shape & function | lumbar value frontal stand right | 30 | 3.94 |
| Back shape & function | spine length sagittal stand extension | 30 | 3.91 |
| Back shape & function | inclination value frontal sit straight | 20 | 3.88 |
| Back shape & function | lumbar value frontal sit right | 30 | 3.75 |
| Back shape & function | sacral sagittal stand extension | 10 | 3.75 |
| Back shape & function | sacral frontal sit straight | 40 | 3.73 |
| Back shape & function | inclination value sagittal sit extension | 40 | 3.63 |
| Back shape & function | sacral frontal sit left | 10 | 3.5 |

SF36 - Short-form 36 Health Status Questionnaire

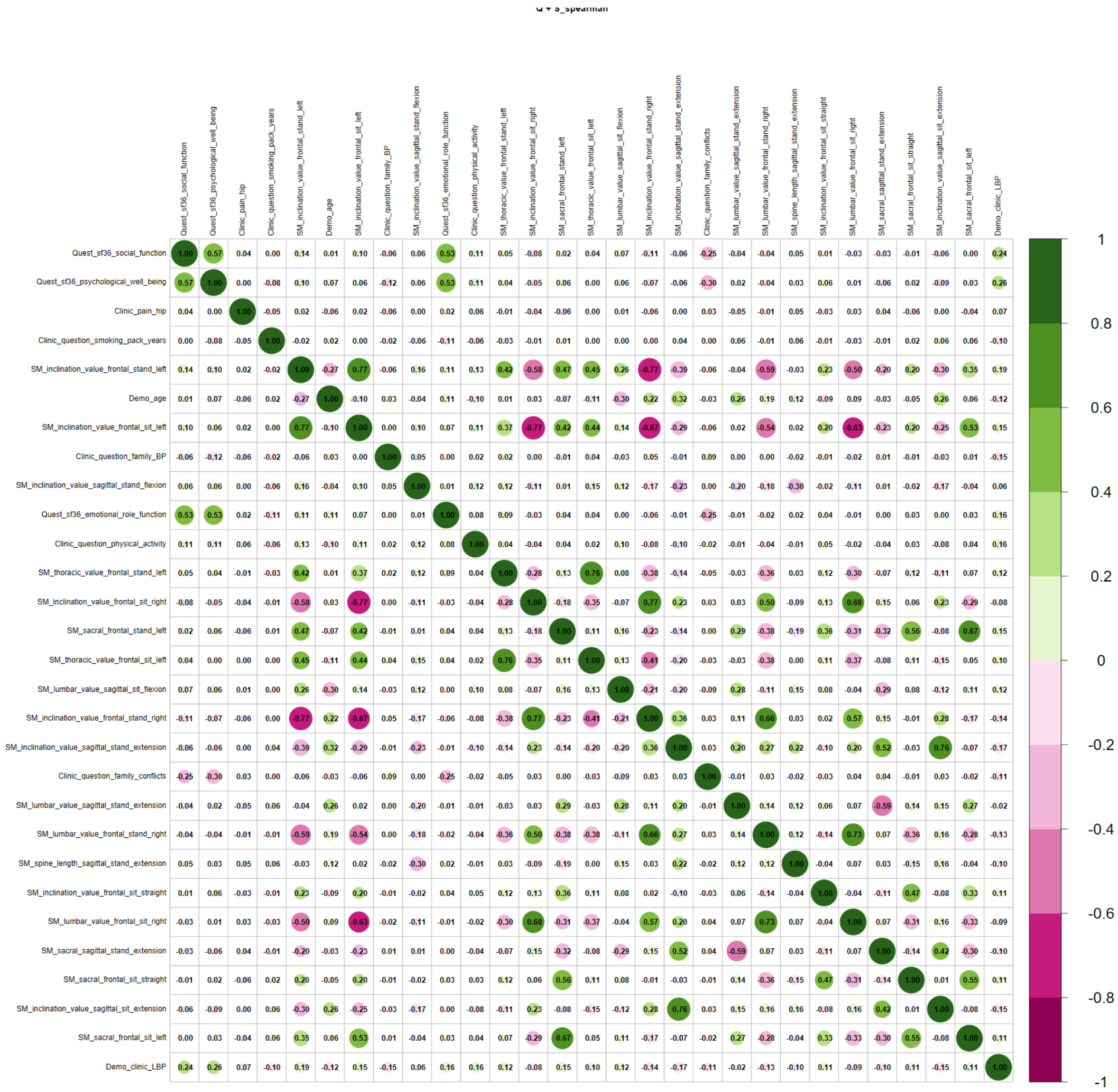

Supplementary Figure S16. Spearman correlation matrix of Boruta selected important features for questionnaire + back shape and function dataset

Supplementary Table S34. Boruta selected important features for questionnaire clinical assessment, and MRI dataset

| Modality | Assessment | Selected (%) | Importance (mean) |
|---|---|---|---|
| questionnaire | SF-36 psychological well being | 100 | 13.36 |
| questionnaire | SF-36 social function | 100 | 13.07 |
| questionnaire | hip pain | 100 | 9.71 |
| clinical examination | cervical axial rotate left | 100 | 9.47 |
| MRI | IVD herniation L4 - L5 | 100 | 9.12 |

| | | | |
|---|---|---|---|
| clinical examination | general mobility | 100 | 7.96 |
| MRI | IVD degeneration L2 - L3 | 100 | 7.6 |
| clinical examination | hip flexion left | 100 | 7.19 |
| MRI | spinal canal width L2 | 100 | 6.72 |
| clinical examination | sit to stand 30sec | 100 | 6.4 |
| MRI | IVD degeneration L4 - L5 | 100 | 6.32 |
| clinical examination | hip internal rotate right | 100 | 6.22 |
| clinical examination | hip flexion right | 100 | 6.09 |
| MRI | IVD degeneration L5 - S1 | 100 | 6.04 |
| clinical examination | hip abduction left | 100 | 5.59 |
| MRI | osteochondrosis L5 - S1 | 90 | 5.29 |
| clinical examination | hip abduction right | 50 | 5.05 |
| MRI | IVD herniation L5 - S1 | 90 | 5 |
| clinical examination | cervical axial rotate right | 100 | 4.83 |
| MRI | IVD degeneration L3 - L4 | 90 | 4.75 |
| MRI | facet joint L5/S1 - L5/L6 right | 40 | 4.7 |
| clinical examination | rigid muscle | 70 | 4.63 |
| MRI | osteochondrosis L4 - L5 | 60 | 4.47 |
| clinical examination | hip internal rotate left | 70 | 4.18 |
| questionnaire | physical activity 150min/week | 20 | 4.15 |
| MRI | facet joint L3 - L4 right | 40 | 4.07 |
| MRI | spinal canal width L1 | 40 | 3.98 |
| questionnaire | living situation | 10 | 3.94 |
| Demographic | age | 40 | 3.9 |
| questionnaire | SF-36 emotional role function | 10 | 3.83 |
| clinical examination | square shoulders | 20 | 3.75 |
| MRI | IVD degeneration L1 - L2 | 20 | 3.65 |
| questionnaire | family back pain | 10 | 3.41 |
| clinical examination | thoracic lumbar inclination | 10 | 3.35 |

IVD – intervertebral disc, SF36 - Short-form 36 Health Status Questionnaire

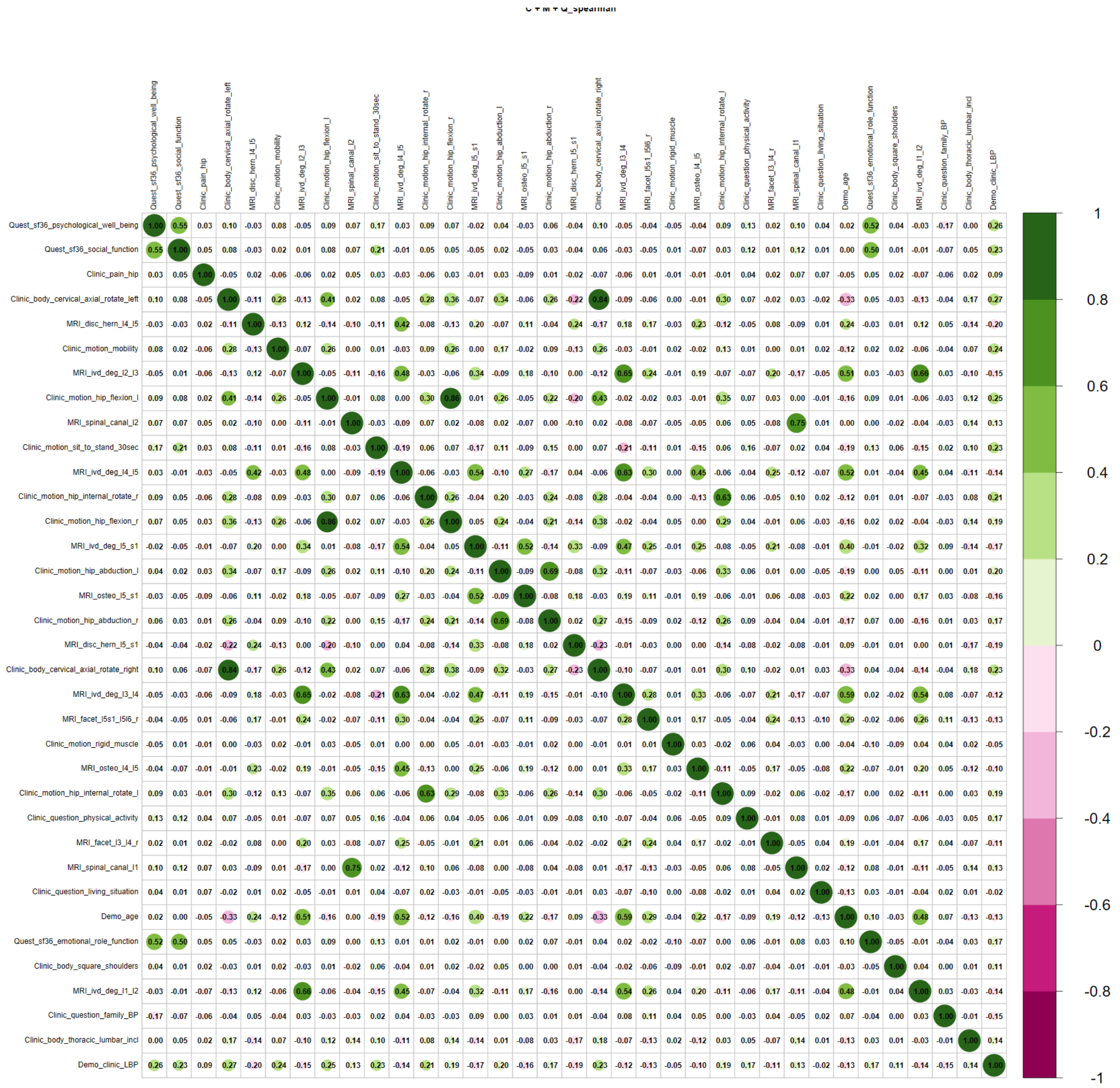

Supplementary Figure S17. Spearman correlation matrix of Boruta selected important features for questionnaire + clinical assessment + MRI dataset

Supplementary Table S35. Boruta selected important features for clinical assessment, MRI, and superficial spine morphology dataset

| Modality | Variable | selected (%) | Importance (mean) |
|---|---|---|---|
| Clinical | cervical axial rotate left | 100 | 13.13 |
| MRI | intervertebral disc herniation L4 - L5 | 100 | 12.85 |
| Clinical | hip flexion left | 100 | 10.11 |
| Clinical | sit to stand 30sec | 100 | 8.61 |
| Clinical | general mobility | 100 | 8.54 |

| | | | |
|---|---|---|---|
| Clinical | hip flexion right | 100 | 8.24 |
| MRI | spinal canal stenosis L2 | 100 | 7.24 |
| MRI | intervertebral disc degeneration L2 - L3 | 100 | 6.58 |
| MRI | intervertebral disc degeneration L4 - L5 | 100 | 6.39 |
| MRI | osteochondrosis intervertebralis L4 - L5 | 100 | 6.38 |
| MRI | intervertebral disc degeneration L3 - L4 | 100 | 6.07 |
| Clinical | cervical axial rotate right | 100 | 6.06 |
| Clinical | rigid muscle | 100 | 5.6 |
| MRI | intervertebral disc degeneration L5 - S1 | 100 | 5.46 |
| Clinical | thoracic lumbar inclination | 10 | 5.4 |
| MRI | osteochondrosis intervertebralis L5 - S1 | 90 | 5.3 |
| MRI | facet joint L3 - L4 right | 20 | 5.19 |
| Clinical | hip internal rotate right | 70 | 4.88 |
| MRI | spinal canal stenosis L1 | 70 | 4.82 |
| MRI | intervertebral disc herniation L5 - S1 | 90 | 4.66 |
| Clinical | hip internal rotate left | 40 | 4.52 |
| Clinical | chin sternum distance | 70 | 4.41 |
| MRI | intervertebral disc degeneration L1 - L2 | 40 | 4.41 |
| Back shape & function | inclination value sagittal sit flexion | 20 | 4.34 |
| Back shape & function | inclination value sagittal stand extension | 40 | 4.27 |
| Back shape & function | lumbar value sagittal sit extension | 40 | 4.18 |
| Back shape & function | lumbar value frontal stand right | 20 | 4.18 |
| Back shape & function | inclination value frontal stand right | 20 | 4.05 |
| | inclination value sagittal sit extension | 60 | 4 |
| MRI | facet joint L5/S1 - L5/L6 right | 30 | 3.96 |
| MRI | intervertebral disc herniation L3 - L4 | 30 | 3.95 |
| Clinical | hip abduction left | 30 | 3.91 |
| Demographic | age | 50 | 3.84 |
| Clinical | asymmetrical waist triangle | 10 | 3.78 |
| Back shape & function | thoracic value frontal stand right | 10 | 3.67 |
| Back shape & function | inclination value frontal stand left | 10 | 3.64 |
| Clinical | hip abduction right | 40 | 3.6 |
| Back shape & function | spine length sagittal stand extension | 20 | 3.51 |
| Back shape & function | thoracic value sagittal stand extension | 10 | 3.51 |
| MRI | facet joint L5/S1 - L5/L6 left | 40 | 3.5 |

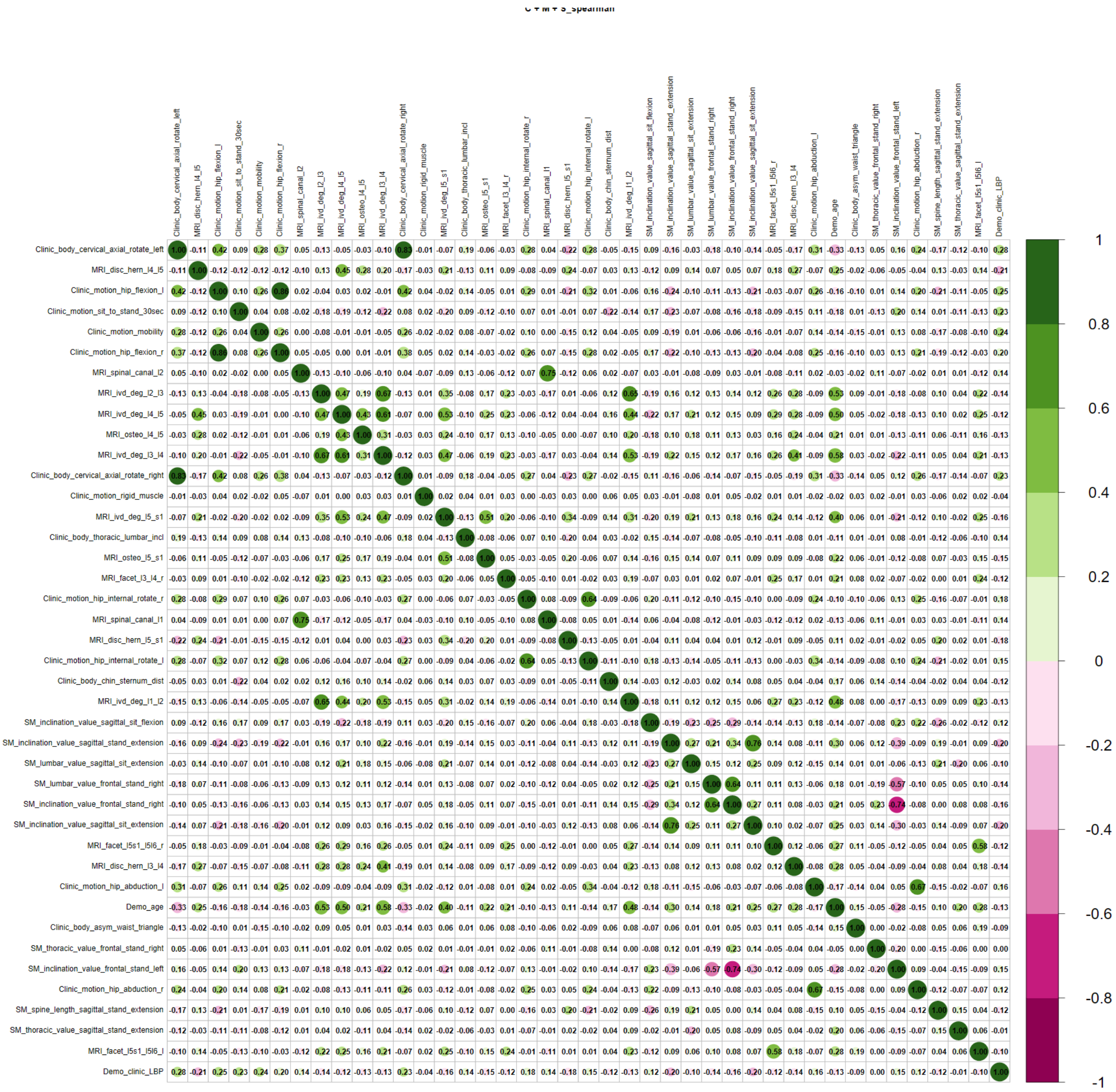

332

333 Supplementary Figure S18. Spearman correlation matrix of Boruta selected
334 important features for MRI + clinical assessment + back shape and function
335 dataset

336

337 Supplementary Table S36. Boruta selected important features for questionnaire
338 clinical assessment, and superficial spine morphology dataset

| Modality | Assessment | Selected (%) | Importance (mean) |
|---|---|---|---|
| Questionnaire | SF36 psychological well being | 100 | 16.99 |
| Questionnaire | SF36 social function | 100 | 16.03 |
| Questionnaire | hip pain | 100 | 11.21 |
| Clinical | mobility | 100 | 10.6 |

| | | | |
|---|---|---|---|
| Clinical | cervical axial rotate left | 100 | 10.18 |
| Clinical | rigid muscle | 100 | 8.56 |
| Clinical | sit to stand 30sec | 100 | 8.11 |
| Clinical | smoking pack years | 100 | 8.06 |
| Clinical | physical activity 150min/week | 90 | 6.14 |
| Back shape & function | inclination value frontal sit left | 20 | 5.39 |
| Clinical | hip abduction left | 100 | 5.27 |
| Clinical | family back pain | 100 | 5.25 |
| Demographic | age | 100 | 5.09 |
| Clinical | cervical axial rotate right | 100 | 4.95 |
| Back shape & function | inclination value frontal stand left | 100 | 4.88 |
| Back shape & function | sacral frontal sit straight | 50 | 4.72 |
| Back shape & function | sacral frontal stand left | 80 | 4.57 |
| Questionnaire | SF36 emotional role function | 40 | 4.31 |
| Back shape & function | inclination value frontal sit straight | 10 | 4.31 |
| Clinical | hip flexion right | 90 | 4.25 |
| Clinical | back form | 80 | 4.24 |
| Back shape & function | lumbar value frontal sit right | 10 | 4.11 |
| Clinical | hip abduction right | 70 | 4.08 |
| Back shape & function | lumbar value frontal stand right | 10 | 4.08 |
| Back shape & function | thoracic value frontal stand left | 40 | 4.06 |
| Clinical | hip flexion left | 80 | 3.94 |
| Clinical | family conflicts | 30 | 3.89 |
| Back shape & function | sacral frontal sit left | 10 | 3.87 |
| Back shape & function | thoracic value frontal sit left | 30 | 3.84 |
| Clinical | hip internal rotate right | 20 | 3.83 |
| Clinical | thoracic lumbar lat bend right | 80 | 3.81 |
| Clinical | shober | 10 | 3.73 |
| Back shape & function | inclination value sagittal stand extension | 50 | 3.65 |
| Back shape & function | spine length sagittal stand extension | 10 | 3.62 |
| Back shape & function | lumbar value sagittal stand extension | 20 | 3.59 |
| Back shape & function | lumbar value sagittal sit flexion | 10 | 3.48 |
| Back shape & function | thoracic value sagittal stand extension | 10 | 3.37 |
| Back shape & function | sacral sagittal stand extension | 10 | 3.36 |
| Questionnaire | job load 1 | 10 | 3.32 |
| Back shape & function | inclination value sagittal sit extension | 10 | 3.31 |

SF36 - Short-form 36 Health Status Questionnaire

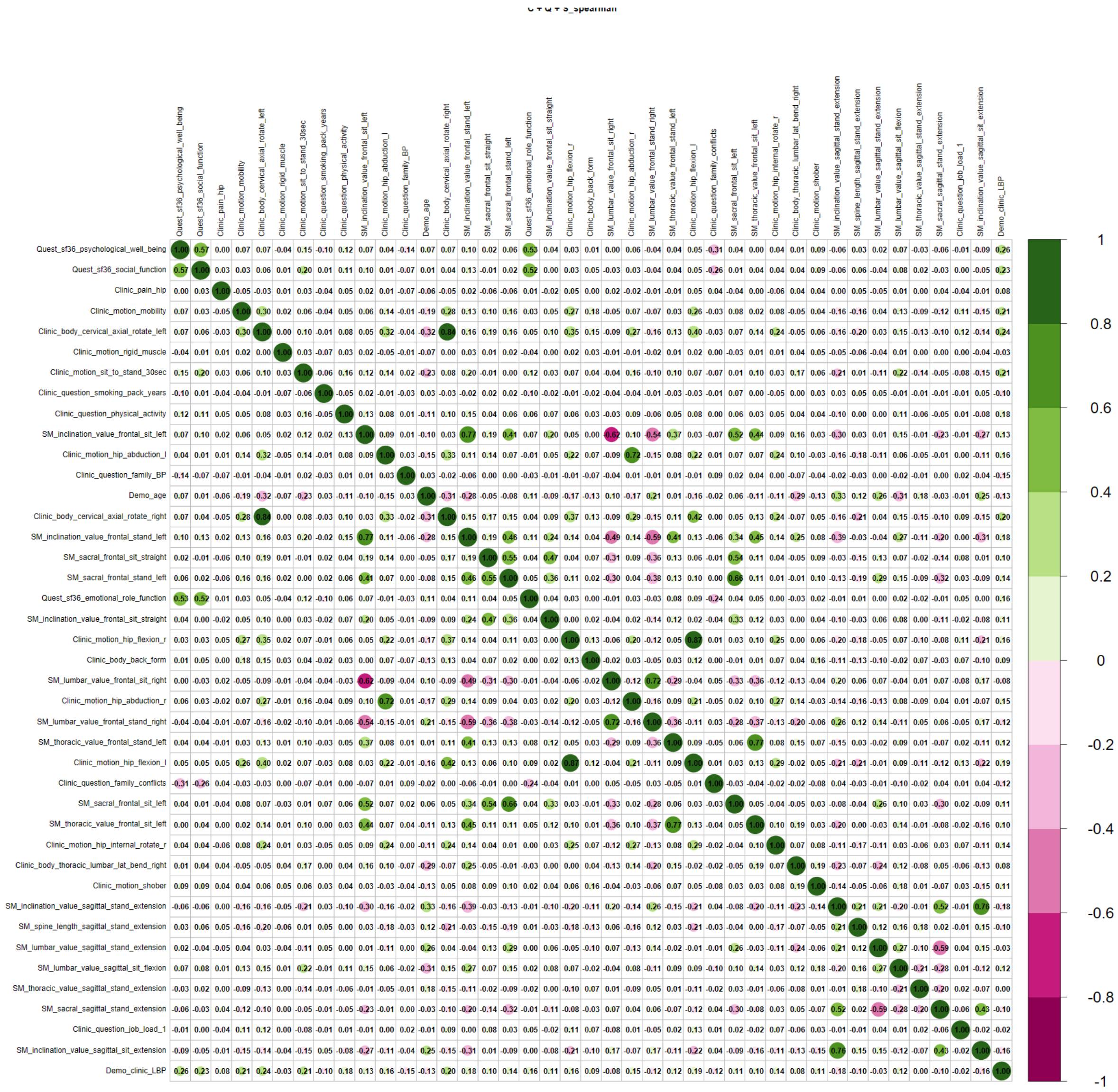

Supplementary Figure S19. Spearman correlation matrix of Boruta selected important features for questionnaire + clinical assessment + back shape and function dataset

Supplementary Table S37. Boruta selected important features for questionnaire, superficial spine morphology, and MRI dataset

| Modality | Variable | Selected (%) | Importance (mean) |
|---|---|---|---|
| Questionnaire | SF36 social function | 100 | 16.92 |
| Questionnaire | SF36 psychological well being | 100 | 15.13 |
| MRI | intervertebral disc herniation L4 - L5 | 100 | 12.69 |
| MRI | spinal canal stenosis L2 | 100 | 9.91 |

| | | | |
|---|---|---|---|
| Questionnaire | hip pain | 100 | 8 |
| MRI | intervertebral disc degeneration L4 - L5 | 100 | 7.79 |
| MRI | intervertebral disc degeneration L2 - L3 | 100 | 7.64 |
| Back shape & function | spine length sagittal stand extension | 100 | 6.97 |
| MRI | intervertebral disc degeneration L5 - S1 | 100 | 6.46 |
| MRI | spinal canal stenosis L1 | 100 | 5.92 |
| Back shape & function | inclination value sagittal stand extension | 90 | 5.37 |
| MRI | facet joint L3 - L4 right | 50 | 5.27 |
| MRI | osteochondrosis intervertebralis L5 - S1 | 20 | 5.15 |
| Back shape & function | lumbar value sagittal sit extension | 30 | 5.13 |
| MRI | intervertebral disc degeneration L3 - L4 | 70 | 5.03 |
| Back shape & function | inclination value frontal stand left | 20 | 4.97 |
| Back shape & function | inclination value sagittal sit extension | 50 | 4.95 |
| MRI | intervertebral disc herniation L5 - S1 | 70 | 4.79 |
| Questionnaire | physical activity 150min/week | 20 | 4.61 |
| Back shape & function | sacral sagittal stand extension | 60 | 4.59 |
| MRI | osteochondrosis intervertebralis L4 - L5 | 70 | 4.55 |
| Back shape & function | lumbar value frontal stand right | 10 | 4.45 |
| MRI | spinal canal stenosis L3 | 10 | 4.44 |
| Questionnaire | IPAQ met sum | 30 | 4.41 |
| Back shape & function | sacral frontal sit straight | 10 | 4.35 |
| Demographic | age | 10 | 4.19 |
| Back shape & function | sacral frontal sit left | 10 | 4.13 |
| Back shape & function | inclination value sagittal sit flexion | 20 | 4.01 |
| Back shape & function | thoracic value frontal sit left | 10 | 3.97 |
| Back shape & function | lumbar value frontal stand left | 10 | 3.92 |
| MRI | facet joint L5/S1 - L5/L6 right | 10 | 3.66 |
| MRI | facet joint L4 - L5 right | 10 | 3.65 |
| MRI | intervertebral disc degeneration L1 - L2 | 10 | 3.62 |
| Questionnaire | SF36 emotional role function | 10 | 3.29 |

SF36 - Short-form 36 Health Status Questionnaire

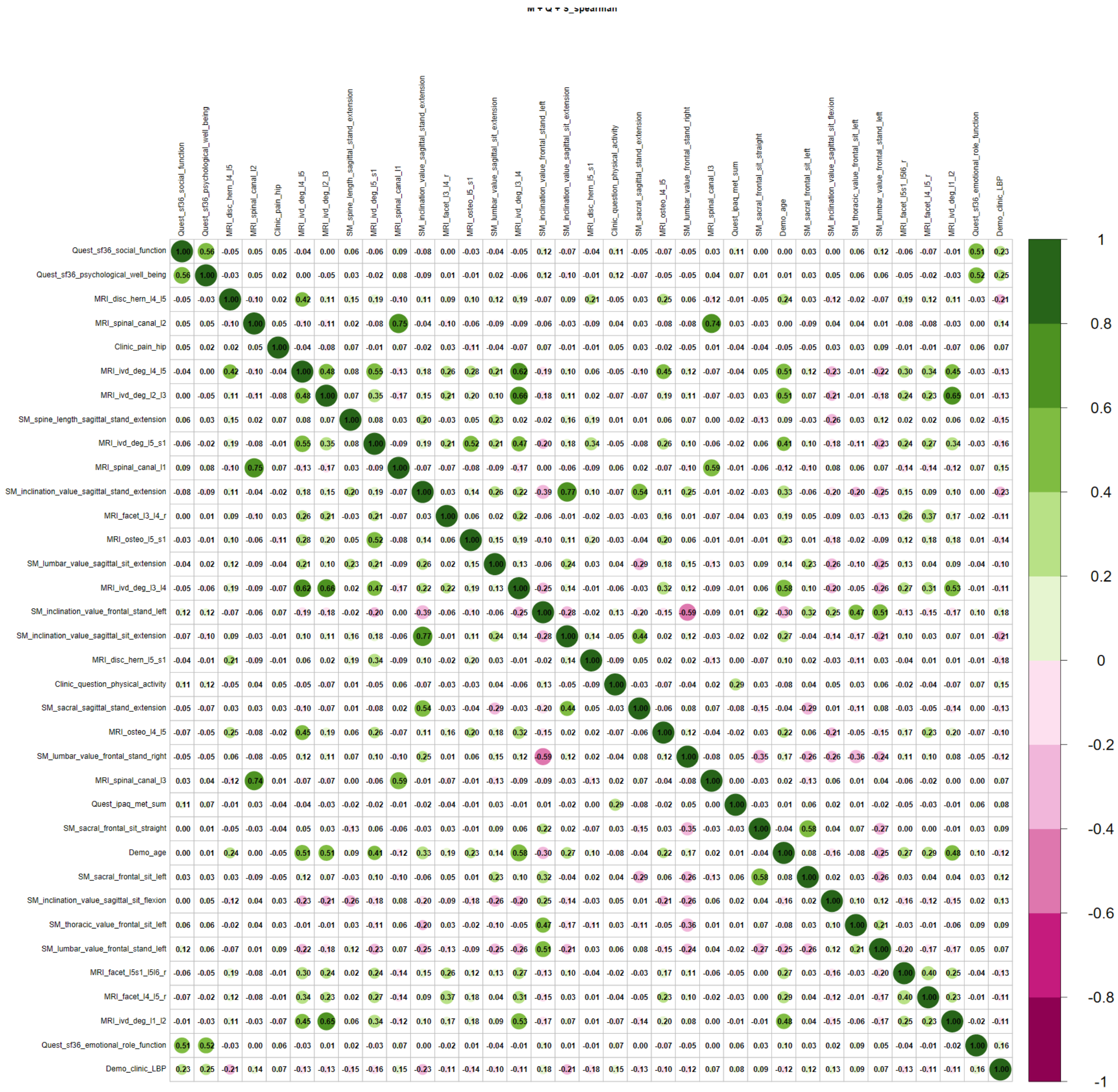

Supplementary Figure S20. Spearman correlation matrix of Boruta selected important features for MRI + questionnaire + back shape and function dataset

Supplementary Table S38. Boruta selected important features for clinical assessment, MRI, questionnaire, and superficial spine morphology dataset

| Modality | Assessment | Selected (%) | Importance (mean) |
|---|---|---|---|
| Questionnaire | SF36 social function | 100 | 13.04 |
| Questionnaire | SF36 psychological well being | 100 | 11.58 |
| Questionnaire | Clinic pain hip | 100 | 9.43 |
| MRI | intervertebral disc herniation L4 - L5 | 100 | 8.61 |
| Clinical | cervical axial rotate left | 100 | 8.6 |

| | | | |
|---|---|---|---|
| MRI | intervertebral disc degeneration L2 - L3 | 100 | 6.8 |
| Clinical | hip flexion left | 100 | 6.79 |
| Clinical | general mobility | 100 | 6.62 |
| MRI | intervertebral disc degeneration L4 - L5 | 100 | 6.45 |
| MRI | spinal canal stenosis L2 | 90 | 6.28 |
| MRI | intervertebral disc degeneration L5 - S1 | 100 | 5.81 |
| Clinical | hip flexion right | 100 | 5.68 |
| Clinical | sit to stand 30sec | 100 | 5.65 |
| Clinical | hip internal rotate right | 90 | 5.62 |
| MRI | osteochondrosis intervertebralis L5 - S1 | 90 | 5.09 |
| Back shape & function | lumbar value sagittal sit extension | 60 | 5.01 |
| MRI | spinal canal stenosis L1 | 10 | 4.96 |
| Clinical | hip abduction left | 80 | 4.8 |
| Back shape & function | spine length sagittal stand extension | 70 | 4.75 |
| Clinical | rigid muscle | 60 | 4.75 |
| Back shape & function | inclination value sagittal stand extension | 100 | 4.73 |
| Questionnaire | SF36 emotional role function | 20 | 4.65 |
| MRI | osteochondrosis intervertebralis L4 - L5 | 80 | 4.63 |
| Clinical | cervical axial rotate right | 90 | 4.62 |
| Back shape & function | sacral sagittal stand extension | 10 | 4.5 |
| Clinical | Clinic question physical activity | 60 | 4.47 |
| MRI | intervertebral disc degeneration L3 - L4 | 80 | 4.43 |
| MRI | facet joint L3 - L4 right | 10 | 4.41 |
| Back shape & function | inclination value sagittal sit extension | 60 | 4.34 |
| Clinical | hip abduction right | 30 | 4.27 |
| Clinical | hip internal rotate left | 40 | 4.02 |
| MRI | intervertebral disc herniation L5 - S1 | 70 | 3.97 |
| Demographic | age | 10 | 3.77 |
| Back shape & function | inclination value sagittal sit flexion | 10 | 3.68 |
| Clinical | thomas handle right | 10 | 3.41 |

SF36 - Short-form 36 Health Status Questionnaire, IPAQ – International Physical Activity Questionnaire

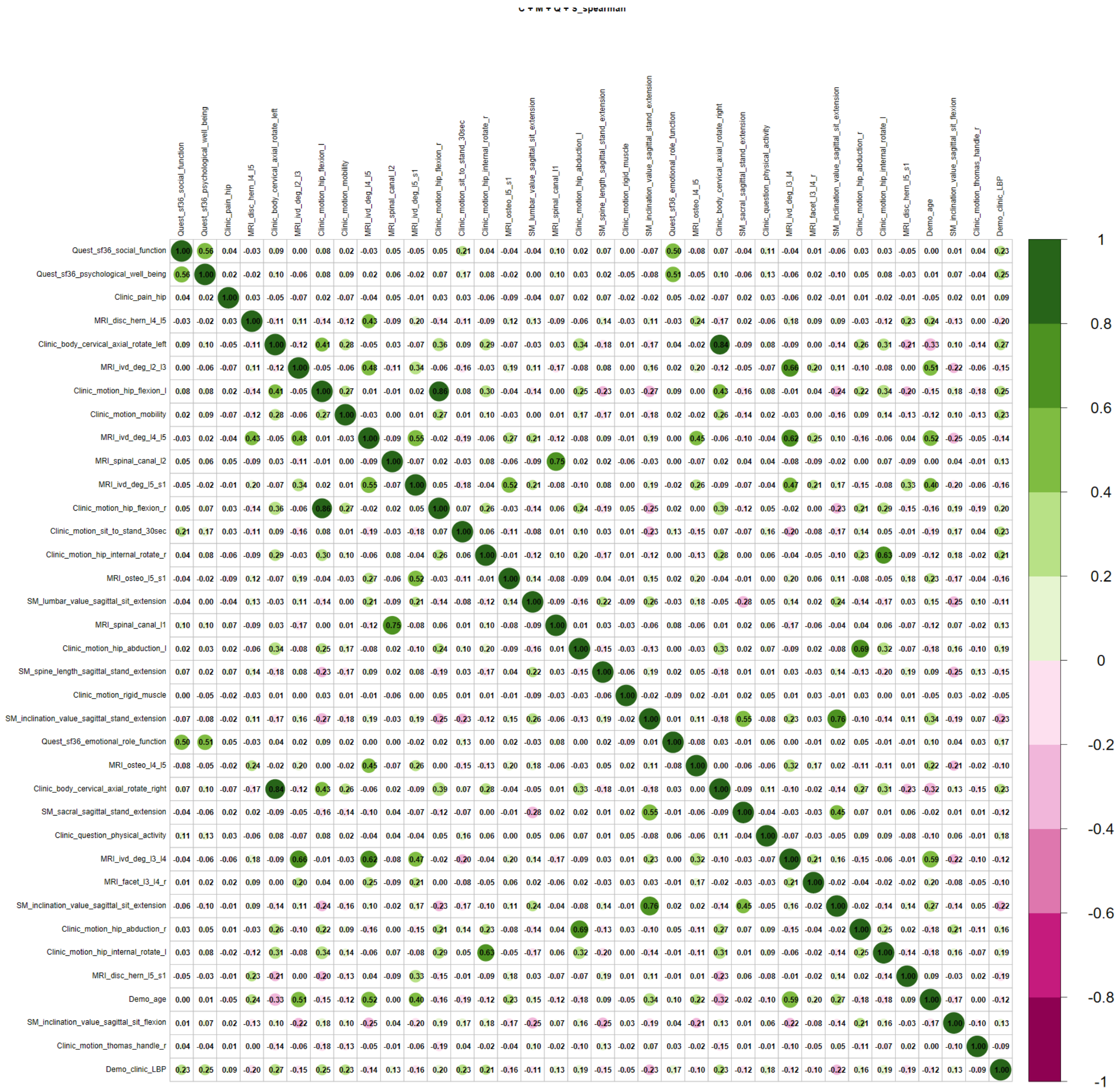

Supplementary Figure S21. Spearman correlation matrix of Boruta selected important features for questionnaire + clinical assessment + back shape and function + MRI dataset

Supplementary Table S39. Overall feature importance and robustness across all modality datasets.

| Modality | Variable | selected (%) | Importance (mean) |
|---|---|---|---|
| Questionnaire | SF36 social function | 100 | 16.54 |
| Questionnaire | SF36 psychological well being | 100 | 16.44 |
| MRI | intervertebral disc herniation L4 - L5 | 100 | 13.33 |
| Clinic | cervical axial rotate left | 100 | 11.76 |
| Questionnaire | Clinic pain hip | 100 | 10.57 |
| Clinic | mobility | 100 | 10.03 |

| | | | |
|---|---|---|---|
| Clinic | sit to stand 30sec | 100 | 9.17 |
| MRI | spinal canal stenosis L2 | 98.75 | 8.63 |
| Clinic | rigid muscle | 91.25 | 7.36 |
| MRI | intervertebral disc degeneration L4 - L5 | 100 | 7.28 |
| MRI | intervertebral disc degeneration L2 - L3 | 100 | 7.21 |
| Clinic | hip flexion left | 93.75 | 6.88 |
| MRI | intervertebral disc degeneration L3 - L4 | 91.25 | 6.1 |
| Clinic | hip flexion right | 97.5 | 5.97 |
| MRI | osteochondrosis intervertebralis L4 - L5 | 78.75 | 5.92 |
| Clinic | cervical axial rotate right | 98.75 | 5.89 |
| MRI | spinal canal stenosis L1 | 77.5 | 5.88 |
| MRI | intervertebral disc degeneration L5 - S1 | 95 | 5.7 |
| MRI | intervertebral disc herniation L5 - S1 | 90 | 5.32 |
| Questionnaire | physical activity 150min/week | 58.75 | 5.04 |
| Clinic | hip abduction left | 77.5 | 5.01 |
| MRI | facet joint L3 - L4 right | 57.5 | 4.97 |
| MRI | osteochondrosis intervertebralis L5 - S1 | 67.5 | 4.88 |
| Questionnaire | smoking pack years | 50 | 4.76 |
| Back shape & function | inclination value frontal stand left | 58.75 | 4.49 |
| Clinic | hip internal rotate right | 65 | 4.46 |
| Back shape & function | inclination value sagittal stand extension | 57.5 | 4.46 |
| Clinic | hip abduction right | 50 | 4.3 |
| Back shape & function | inclination value sagittal sit extension | 52.5 | 4.28 |
| MRI | facet joint L5/S1 - L5/L6 right | 48.75 | 4.07 |
| Back shape & function | spine length sagittal stand extension | 50 | 4.04 |
| Back shape & function | lumbar value frontal stand right | 37.5 | 3.69 |
| MRI | intervertebral disc degeneration L1 - L2 | 38.75 | 3.64 |
| Clinic | family back pain | 51.25 | 3.45 |
| Back shape & function | sacral frontal stand left | 46.25 | 3.15 |
| Questionnaire | SF36 emotional role function | 13.75 | 3.09 |
| Back shape & function | sacral frontal sit left | 20 | 2.92 |
| Back shape & function | lumbar value sagittal sit extension | 28.75 | 2.76 |
| Clinic | hip internal rotate left | 35 | 2.71 |
| Back shape & function | thoracic value sagittal stand extension | 30 | 2.65 |
| Back shape & function | inclination value frontal stand right | 32.5 | 2.63 |
| Clinic | back form | 43.75 | 2.59 |
| Clinic | thoracic lumbar inclination | 15 | 2.59 |
| Back shape & function | thoracic value frontal sit left | 28.75 | 2.54 |
| Back shape & function | inclination value frontal sit left | 26.25 | 2.3 |
| Clinic | chin sternum distance | 31.25 | 2.16 |
| Back shape & function | thoracic value frontal stand left | 25 | 2.16 |
| Clinic | thoracic lumbar lat bend right | 33.75 | 2.08 |
| Back shape & function | inclination value sagittal sit flexion | 7.5 | 2.06 |

| | | | |
|---|---|---|---|
| MRI | facet joint L3 - L4 left | 21.25 | 2.05 |
| Back shape & function | sacral frontal sit straight | 18.75 | 2.05 |
| Clinic | shober | 11.25 | 2.05 |
| Back shape & function | sacral sagittal stand extension | 11.25 | 2.02 |
| Back shape & function | lumbar value sagittal sit flexion | 22.5 | 1.98 |
| Back shape & function | lumbar value sagittal stand extension | 6.25 | 1.88 |
| Clinic | cervical lateral bend left | 13.75 | 1.62 |
| Questionnaire | living situation | 7.5 | 1.56 |
| MRI | facet joint L5/S1 - L5/L6 left | 23.75 | 1.54 |
| Questionnaire | job load 1 | 11.25 | 1.51 |
| Back shape & function | lumbar value frontal sit right | 7.5 | 1.49 |
| MRI | intervertebral disc herniation L3 - L4 | 8.75 | 1.44 |
| Clinic | asymmetrical waist triangle | 7.5 | 1.36 |
| MRI | spinal canal stenosis L3 | 3.75 | 1.05 |
| Back shape & function | lumbar value frontal stand left | 7.5 | 1.04 |
| Questionnaire | BRSQ intrinsic motivation | 6.25 | 1.02 |
| Questionnaire | family conflicts | 6.25 | 1.02 |
| Back shape & function | inclination value frontal sit straight | 3.75 | 1.02 |
| MRI | spinal canal stenosis L4 | 8.75 | 1.01 |
| MRI | intervertebral disc herniation L2 - L3 | 15 | 1 |
| Back shape & function | inclination value frontal sit right | 2.5 | 1 |
| Back shape & function | lumbar value frontal stand straight | 8.75 | 0.99 |
| MRI | facet joint L4 - L5 right | 3.75 | 0.98 |
| Clinic | square shoulders | 3.75 | 0.98 |
| Clinic | thomas handle right | 2.5 | 0.93 |
| Back shape & function | thoracic value frontal stand right | 2.5 | 0.89 |
| MRI | facet joint L1 - L2 right | 3.75 | 0.64 |
| Back shape & function | inclination value sagittal stand flexion | 1.25 | 0.64 |
| MRI | facet joint L1 - L2 left | 2.5 | 0.57 |
| Questionnaire | IPAQ met sum | 3.75 | 0.55 |
| Clinic | cervical inclination | 1.25 | 0.54 |
| Back shape & function | lumbar value frontal sit straight | 1.25 | 0.53 |
| Clinic | hip extension right | 1.25 | 0.52 |
| Back shape & function | sacral sagittal sit straight | 1.25 | 0.51 |
| Questionnaire | BRSQ integrated regulation | 2.5 | 0.48 |
| Clinic | lumbar bulge | 1.25 | 0.48 |
| Clinic | thoracic lumbar axial rotation left | 1.25 | 0.48 |
| Clinic | roussoly type | 1.25 | 0.46 |
| Back shape & function | sacral frontal stand straight | 1.25 | 0.46 |
| Clinic | cervical reclination | 5 | 0.45 |
| Back shape & function | thoracic value sagittal sit extension | 2.5 | 0.45 |
| Clinic | finger floor distance | 1.25 | 0.45 |
| Clinic | hip adduction right | 1.25 | 0.42 |
| Back shape & function | sacral sagittal stand straight | 2.5 | 0.41 |

### Supplementary Data 4: Univariate statistics

#### Supplementary Table S40: Demographic Wilcoxon-Mann-Whitney test (continuous and ordinal)

Median values and corresponding IQR for patients and controls with statistical u-values, z-values, r-values and p-values of the Wilcoxon-Mann-Whitney test after FWE correction.

| Feature | Patients | | Controls | | Statistic | | | |
|---|---|---|---|---|---|---|---|---|
| | Median | IQR | Median | IQR | u-value | z-value | r-value | p-value |
| Age | 44.0 | 20.25 | 39.0 | 22.00 | 320583 | 4.08 | 0.120 | < .001 |
| BMI | 23.3 | 4.03 | 23.5 | 3.86 | 295745 | -0.30 | -0.009 | 1 |

BMI – Body mass index

#### Supplementary Table S41: Demographic Chi2 Test (nominal)

Statistical Chi$^2$-values, ω-values and p-values of the Chi-Square test after FWE correction.

| Feature | Statistic | | |
|---|---|---|---|
| | x2-value | Cohen's ω | p-value |
| Sex | 3.37 | 0.05 | .205 |

#### Supplementary Table S42: Questionnaire Wilcoxon-Mann-Whitney test (continuous and ordinal)

Median values and corresponding IQR for patients and controls with statistical u-values, z-values, r-values and p-values of the Wilcoxon-Mann-Whitney test after FWE correction. As some ordinal variables (e.g. "Alcohol frequency") were expressed as text, only statistical results are shown in the table.

| Variable | Patients | | Controls | | Statistic | | | |
|---|---|---|---|---|---|---|---|---|
| | Median | IQR | Median | IQR | u-value | z-value | r-value | p-value |
| Alcohol frequency | | | | | 216527 | 0.91 | 0.029 | 1 |
| Alcohol glasses | | | | | 211454 | -0.33 | -0.011 | 1 |
| BRSQ external regulation | 3 | 3 | 3 | 2 | 215646 | 0.69 | 0.022 | 1 |
| BRSQ integrated regulation | 9 | 4 | 9 | 4 | 205967 | -1.53 | -0.049 | 1 |
| BRSQ intrinsic motivation | 10 | 4 | 10 | 3 | 198539 | -3.27 | -0.104 | .019 |
| COVID19 activity | | | | | 217410 | 1.44 | 0.046 | 1 |
| IPAQ met sum | 834 | 868 | 910 | 772 | 206393 | -1.42 | -0.045 | 1 |
| Job load 1 | | | | | 225160 | 3.67 | 0.117 | .005 |
| Job load 1 duration | 8 | 3 | 7 | 3 | 214772 | 0.48 | 0.015 | 1 |
| Physical activity | | | | | 194505 | -4.77 | -0.152 | < .001 |
| SF36 emotional role function | 100 | 33 | 100 | 0 | 196952 | -4.39 | -0.140 | < .001 |
| SF36 psychological well-being | 72 | 20 | 80 | 16 | 178507 | -7.74 | -0.247 | < .001 |
| SF36 social function | 88 | 38 | 100 | 13 | 183751 | -7.08 | -0.225 | < .001 |
| Smoking pack years | 0 | 0 | 0 | 0 | 221047 | 3.33 | 0.106 | .016 |

| | | | | | | | | |
|---|---|---|---|---|---|---|---|---|
| SRBAI sum | 16 | 8 | 17 | 7 | 201693 | -2.49 | -0.079 | .206 |

BRSQ - Behavioural Regulation in Sport Questionnaire, IPAQ – International Physical Activity Questionnaire, SF36 - Short-form 36 Health Status Questionnaire, SRBAI - Self-Report Behavioural Automaticity Index

#### Supplementary Table S43: Questionnaire Chi2 Test (nominal)

Statistical Chi$^2$-values, ω-values and p-values of the Chi-Square test after FWE correction.

| Variable | Statistic | | |
|---|---|---|---|
| | x2-value | Cohen's ω | p-value |
| Family back pain | 20.88 | 0.15 | .004 |
| Family conflicts | 9.95 | 0.10 | .007 |
| Hip pain | 40.17 | 0.20 | .004 |
| Job field | 28.63 | 0.17 | .112 |
| Job posture | 5.37 | 0.07 | .303 |
| Living situation | 13.86 | 0.12 | .006 |
| Prior surgeries | 2.25 | 0.05 | .303 |
| Psychological stress | 13.37 | 0.12 | .007 |

#### Supplementary Table S44: Clinical Physcial Assessment Wilcoxon-Mann-Whitney test (continuous and ordinal)

Median values and corresponding IQR for patients and controls with statistical u-values, z-values, r-values and p-values of the Wilcoxon-Mann-Whitney test after FWE correction.

| Variable | Patients | | Controls | | Statistic | | | |
|---|---|---|---|---|---|---|---|---|
| | Median | IQR | Median | IQR | u-value | z-value | r-value | p-value |
| Cervical axial rotate left | 70 | 15 | 80 | 20 | 221595 | -7.98 | -0.243 | < .001 |
| Cervical axial rotate right | 75 | 20 | 80 | 20 | 227209 | -6.86 | -0.209 | < .001 |
| Cervical inclination | 45 | 10 | 45 | 10 | 256777 | -0.97 | -0.029 | 1 |
| Cervical lateral bend left | 35 | 10 | 35 | 15 | 248306 | -2.65 | -0.081 | .225 |
| Cervical lateral bend right | 35 | 10 | 40 | 15 | 245196 | -3.27 | -0.099 | .033 |
| Cervical reclination | 45 | 25 | 50 | 20 | 253672 | -1.57 | -0.048 | 1 |
| Chin sternum distance | 2 | 2 | 1 | 2 | 280047 | 3.92 | 0.119 | .003 |
| Finger floor distance | 0 | 16 | 0 | 18 | 279513 | 3.55 | 0.108 | .013 |
| Hip abduction left | 50 | 15 | 50 | 15 | 237322 | -4.83 | -0.147 | < .001 |
| Hip abduction right | 50 | 15 | 50 | 15 | 240564 | -4.19 | -0.128 | < .001 |
| Hip adduction left | 25 | 10 | 25 | 10 | 260121 | -0.30 | -0.009 | 1 |
| Hip adduction right | 25 | 10 | 30 | 10 | 260129 | -0.30 | -0.009 | 1 |
| Hip extension left | 10 | 0 | 10 | 0 | 252058 | -2.32 | -0.071 | .529 |
| Hip extension right | 10 | 0 | 10 | 0 | 253208 | -2.02 | -0.062 | 1 |
| Hip external rotate left | 40 | 11 | 40 | 15 | 268678 | 1.42 | 0.043 | 1 |
| Hip external rotate right | 40 | 15 | 40 | 15 | 262804 | 0.24 | 0.007 | 1 |
| Hip flexion left | 120 | 15 | 125 | 20 | 231410 | -5.99 | -0.182 | < .001 |

| | | | | | | | | |
|---|---|---|---|---|---|---|---|---|
| Hip flexion right | 120 | 10 | 130 | 20 | 236225 | -5.03 | -0.153 | < .001 |
| Hip internal rotate left | 30 | 15 | 35 | 10 | 244180 | -3.49 | -0.106 | .016 |
| Hip internal rotate right | 30 | 15 | 35 | 10 | 240555 | -4.21 | -0.128 | < .001 |
| OTT | 3 | 1 | 3 | 1 | 253072 | -1.69 | -0.052 | 1 |
| Roussoly type | 2 | 0 | 2 | 1 | 248275 | -2.99 | -0.091 | .080 |
| Shober | 5 | 1 | 5 | 2 | 243079 | -3.68 | -0.112 | .008 |
| Sit to stand 30sec | 21 | 8 | 23 | 7 | 226819 | -6.83 | -0.208 | < .001 |
| Thoracic lumbar ar left | 40 | 20 | 40 | 20 | 253276 | -1.65 | -0.050 | 1 |
| Thoracic lumbar ar right | 40 | 20 | 40 | 20 | 253955 | -1.51 | -0.046 | 1 |
| Thoracic lumbar inclination | 70 | 21 | 70 | 30 | 244361 | -3.40 | -0.104 | .021 |
| Thoracic lumbar lateral bend left | 30 | 15 | 30 | 10 | 250461 | -2.22 | -0.068 | .655 |
| Thoracic lumbar lateral bend right | 30 | 15 | 35 | 10 | 249415 | -2.43 | -0.074 | .403 |
| Thoracic lumbar reclination | 30 | 15 | 30 | 15 | 258021 | -0.71 | -0.022 | 1 |

#### Supplementary Table S45: Clinical Physcial Assessment Chi2 Test (nominal)

Statistical Chi$^2$-values, ω-values and p-values of the Chi-Square test after FWE correction.

| Variable | Statistic | | |
|---|---|---|---|
| | x2-value | Cohen's ω | p-value |
| Asymmetrical waist triangle | 7.82 | 0.09 | .045 |
| Back form | 24.70 | 0.15 | .006 |
| Lumbar bulge | 13.51 | 0.11 | .018 |
| Mobility | 46.81 | 0.21 | .006 |
| Pelvic tilt frontal | 0.52 | 0.02 | .790 |
| Pelvic tilt sagital | 7.11 | 0.08 | .130 |
| Plump line | 4.73 | 0.07 | .130 |
| Rib hump | 12.17 | 0.11 | .024 |
| Rigid muscle | 50.32 | 0.22 | .006 |
| Sagittal balance | 2.15 | 0.04 | .317 |
| Square shoulders | 15.64 | 0.12 | .010 |
| Thomas handle left | 6.61 | 0.08 | .090 |
| Thomas handle right | 5.23 | 0.07 | .130 |

#### Supplementary Table S46: Superficial spine morphology Wilcoxon-Mann-Whitney test (continuous)

| Variable | Patients | | Controls | | Statistic | | | |
|---|---|---|---|---|---|---|---|---|
| | Median | IQR | Median | IQR | u-value | z-value | r-value | p-value |
| Inclination value frontal sit left | 18 | 11 | 20 | 11 | 248689 | -4.45 | -0.133 | < .001 |
| Inclination value frontal sit right | -24 | 10 | -26 | 11 | 287297 | 2.81 | 0.084 | 0.165 |
| Inclination value frontal sit straight | -4 | 3 | -3 | 2 | 255324 | -3.24 | -0.097 | 0.046 |

| | | | | | | | | |
|---|---|---|---|---|---|---|---|---|
| Inclination value frontal stand left | 20 | 11 | 24 | 11 | 240941 | -5.91 | -0.177 | < .001 |
| Inclination value frontal stand right | -27 | 10 | -29 | 10 | 298681 | 4.95 | 0.148 | < .001 |
| Inclination value frontal stand straight | -3 | 3 | -3 | 3 | 272113 | -0.05 | -0.001 | 1 |
| Inclination value sagittal sit extension | -14 | 11 | -17 | 12 | 299339 | 5.07 | 0.152 | < .001 |
| Inclination value sagittal sit flexion | 84 | 14 | 85 | 14 | 260158 | -2.3 | -0.069 | 0.628 |
| Inclination value sagittal sit straight | 5 | 5 | 5 | 4 | 279373 | 1.32 | 0.04 | 1 |
| Inclination value sagittal stand extension | -19 | 10 | -21 | 11 | 298587 | 4.93 | 0.148 | < .001 |
| Inclination value sagittal stand flexion | 116 | 21 | 118 | 21 | 266091 | -1.18 | -0.035 | 1 |
| Inclination value sagittal stand straight | 1 | 3 | 1 | 3 | 276904 | 0.86 | 0.026 | 1 |
| Lumbar value frontal sit left | 16 | 9 | 17 | 9 | 259322 | -2.45 | -0.074 | 0.423 |
| Lumbar value frontal sit right | -16 | 11 | -17 | 12 | 287942 | 2.93 | 0.088 | 0.12 |
| Lumbar value frontal sit straight | 1 | 6 | 1 | 6 | 282311 | 1.87 | 0.056 | 1 |
| Lumbar value frontal stand left | 20 | 11 | 21 | 10 | 263580 | -1.65 | -0.05 | 1 |
| Lumbar value frontal stand right | -17 | 12 | -20 | 12 | 296706 | 4.57 | 0.137 | < .001 |
| Lumbar value frontal stand straight | 3 | 8 | 2 | 9 | 284300 | 2.24 | 0.067 | 0.671 |
| Lumbar value sagittal sit extension | -33 | 14 | -34 | 14 | 283974 | 2.18 | 0.065 | 0.759 |
| Lumbar value sagittal sit flexion | 23 | 13 | 25 | 11 | 253884 | -3.48 | -0.104 | 0.02 |
| Lumbar value sagittal sit straight | -14 | 15 | -13 | 14 | 263916 | -1.59 | -0.048 | 1 |
| Lumbar value sagittal stand extension | -36 | 12 | -37 | 14 | 278643 | 1.18 | 0.035 | 1 |
| Lumbar value sagittal stand straight | -28 | 12 | -27 | 11 | 264400 | -1.5 | -0.045 | 1 |
| Sacral frontal sit left | -1 | 6 | 0 | 5 | 252795 | -3.69 | -0.111 | 0.009 |
| Sacral frontal sit right | -4 | 5 | -4 | 6 | 265294 | -1.33 | -0.04 | 1 |
| Sacral frontal sit straight | -4 | 5 | -3 | 4 | 256209 | -3.05 | -0.091 | 0.082 |
| Sacral frontal stand left | -3 | 8 | -1 | 7 | 247591 | -4.66 | -0.14 | < .001 |
| Sacral frontal stand right | -6 | 5 | -6 | 7 | 278563 | 1.17 | 0.035 | 1 |
| Sacral frontal stand straight | -5 | 5 | -4 | 5 | 259268 | -2.47 | -0.074 | 0.418 |
| Sacral sagittal sit extension | 8 | 13 | 6 | 14 | 282534 | 1.91 | 0.057 | 1 |
| Sacral sagittal sit flexion | 47 | 15 | 46 | 17 | 274473 | 0.39 | 0.012 | 1 |
| Sacral sagittal sit straight | 8 | 10 | 7 | 10 | 281418 | 1.7 | 0.051 | 1 |
| Sacral sagittal stand extension | 5 | 17 | 2 | 15 | 287444 | 2.83 | 0.085 | 0.157 |
| Sacral sagittal stand flexion | 78 | 22 | 79 | 23 | 273788 | 0.27 | 0.008 | 1 |

| | | | | | | | | |
|---|---|---|---|---|---|---|---|---|
| Sacral sagittal stand straight | 14 | 10 | 13 | 10 | 284501 | 2.28 | 0.068 | 0.632 |
| Spine length sagittal stand extension | 495 | 63 | 486 | 55 | 288916 | 3.11 | 0.093 | 0.07 |
| Thoracic value frontal sit left | 33 | 16 | 34 | 14 | 258113 | -2.68 | -0.08 | 0.235 |
| Thoracic value frontal sit right | -37 | 12 | -37 | 13 | 272398 | 0 | 0 | 1 |
| Thoracic value frontal sit straight | -3 | 10 | -2 | 9 | 262542 | -1.85 | -0.055 | 1 |
| Thoracic value frontal stand left | 32 | 16 | 35 | 14 | 252969 | -3.65 | -0.109 | 0.011 |
| Thoracic value frontal stand right | -36 | 14 | -36 | 13 | 270986 | -0.26 | -0.008 | 1 |
| Thoracic value frontal stand straight | -3 | 9 | -2 | 10 | 263520 | -1.67 | -0.05 | 1 |
| Thoracic value sagittal sit extension | 29 | 21 | 29 | 21 | 270485 | -0.35 | -0.011 | 1 |
| Thoracic value sagittal sit flexion | 73 | 12 | 73 | 13 | 275612 | 0.61 | 0.018 | 1 |
| Thoracic value sagittal sit straight | 40 | 14 | 40 | 13 | 277132 | 0.89 | 0.027 | 1 |
| Thoracic value sagittal stand extension | 34 | 18 | 33 | 20 | 268380 | -0.75 | -0.022 | 1 |
| Thoracic value sagittal stand flexion | 64 | 16 | 64 | 15 | 274505 | 0.4 | 0.012 | 1 |
| Thoracic value sagittal stand straight | 46 | 14 | 45 | 13 | 275202 | 0.53 | 0.016 | 1 |

#### Supplementary Table S47: Spino-pelvic MRI Wilcoxon-Mann-Whitney test (continuous and ordinal)

Median values and corresponding IQR for patients and controls with statistical u-values, z-values, r-values and p-values of the Wilcoxon-Mann-Whitney test after FWE correction.

| **Variable** | **Patients** | | **Controls** | | **Statistic** | | | |
|---|---|---|---|---|---|---|---|---|
| | Median | IQR | Median | IQR | u-value | z-value | r-value | p-value |
| Intervertebral disc herniation L2-L3 | 1 | 0 | 1 | 0 | 142051 | 3.17 | 0.112 | .018 |
| Intervertebral disc herniation L3-L4 | 1 | 0 | 1 | 0 | 144077 | 3.26 | 0.115 | .016 |
| Intervertebral disc herniation L4-L5 | 1 | 1 | 1 | 1 | 153471 | 6.02 | 0.213 | < .001 |
| Intervertebral disc herniation L5-S1 | 1 | 1 | 1 | 1 | 150122 | 4.75 | 0.168 | < .001 |
| Facet L1-L2 left | 1 | 1 | 1 | 1 | 143273 | 2.19 | 0.078 | .114 |
| Facet L1-L2 right | 1 | 1 | 1 | 1 | 143551 | 2.28 | 0.081 | .113 |
| Facet L2-L3 left | 1 | 1 | 1 | 1 | 140482 | 1.19 | 0.042 | .234 |
| Facet L2-L3 right | 1 | 1 | 1 | 1 | 142246 | 1.86 | 0.066 | .188 |
| Facet L3-L4 left | 1 | 1 | 1 | 1 | 144439 | 2.62 | 0.093 | .061 |
| Facet L3-L4 right | 2 | 1 | 1 | 1 | 148587 | 3.98 | 0.141 | .001 |
| Facet L4-L5 left | 2 | 1 | 1 | 1 | 145545 | 2.89 | 0.102 | .042 |
| Facet L4-L5 right | 2 | 1 | 1 | 1 | 145101 | 2.72 | 0.096 | .060 |
| Facet L5-S1 / L5-L6 left | 2 | 1 | 2 | 1 | 145096 | 2.69 | 0.095 | .060 |

| | | | | | | | | |
|---|---|---|---|---|---|---|---|---|
| Facet L5-S1 / L5-L6 right | 2 | 1 | 2 | 1 | 147498 | 3.49 | 0.124 | .009 |
| Intervertebral disc degeneration L1-L2 | 2 | 1 | 1 | 1 | 146699 | 3.26 | 0.115 | .016 |
| Intervertebral disc degeneration L2-L3 | 2 | 1 | 1 | 1 | 147168 | 3.38 | 0.120 | .011 |
| Intervertebral disc degeneration L3-L4 | 2 | 1 | 1 | 1 | 147661 | 3.51 | 0.124 | .009 |
| Intervertebral disc degeneration L4-L5 | 2 | 2 | 2 | 2 | 147691 | 3.43 | 0.121 | .010 |
| Intervertebral disc degeneration L5-S1 | 2 | 1 | 2 | 2 | 150652 | 4.34 | 0.154 | < .001 |
| Osteochondrosis L4-L5 | 1 | 0 | 1 | 0 | 143664 | 3.50 | 0.124 | .009 |
| Osteochondrosis L5-S1 | 1 | 0 | 1 | 0 | 144674 | 3.60 | 0.127 | .006 |
| Spinal cana stenosis l L1 | 17.5 | 1.90 | 18.0 | 1.90 | 123171 | -4.35 | -0.154 | < .001 |
| Spinal canal stenosis L2 | 16.9 | 2.25 | 17.3 | 1.90 | 123548 | -4.23 | -0.150 | < .001 |
| Spinal canal stenosis L3 | 16.5 | 2.10 | 16.8 | 2.20 | 128327 | -2.75 | -0.097 | .060 |
| Spinal canal stenosis L4 | 17.0 | 2.45 | 17.1 | 2.33 | 132006 | -1.61 | -0.057 | .215 |
| Spinal canal stenosis L5 | 17.4 | 2.95 | 17.9 | 2.90 | 128906 | -2.57 | -0.091 | .061 |
